# The State of Health Visiting in England: Workforce Composition, Caseloads and Service Delivery

**DOI:** 10.64898/2026.03.26.26349382

**Authors:** Gabriella Conti, Gabriel Weber Costa, Dylan Celestino D’Mello, Yichen Yu

## Abstract

Health visiting is a statutory, universal service at the core of the Healthy Child Programme. It delivers five mandated health and development reviews for all families with children, alongside non-mandated targeted support when needs are identified. These services are led by Health Visitors (HVs) and delivered by multidisciplinary health visiting teams composed of both HVs and Clinical Skill Mix Staff (CSMS). Since commissioning responsibilities were transferred to Local Authorities in 2015, the service has faced sustained workforce and financial pressures. Drawing on new data collected under the Freedom of Information Act from a large sample of Local Authorities across England, this report documents trends in team composition, caseloads, redeployment during the COVID-19 pandemic, and public spending between 2016 and 2021, and considers their implications for current policy.

The evidence points to a service under sustained strain: most areas have seen substantial reductions in the number of Health Visitors, rising reliance on lower-band staff, and widespread caseloads above recommended levels. Spending on mandated health visiting has fallen by nearly 20% in real terms. Unless workforce rebuilding and sustained investment are prioritised, the ambitions set out in the NHS 10-Year Health Plan and the Department for Education’s new early years strategy will remain structurally constrained. To inform future policy design, we also provide estimates of the staffing increases and associated costs required to restore caseloads to recommended levels under different workforce-mix scenarios.

**Key findings:**

- **Health Visitors fell by 21%** between 2016 and 2021, while **Clinical Skill Mix Staff rose by 33%**; overall workforce capacity contracted.
- Workforce composition shifted across England, with **nearly 80% of Local Authorities experiencing a decline in the HV share** within their teams.
- **Caseload pressures were widespread:** around **74% of Local Authorities exceed the recommended 250 children per practitioner**, with some exceeding 1,000.
- During the first wave of COVID-19, **68.7% of Local Authorities redeployed at least one member** of their health visiting workforce, often for sustained periods; redeployment was more limited in the second wave.
- **Spending on mandated services across all Local Authorities in England fell by almost 20% in real terms** between 2016-17 and 2024-25.
- **Spending cuts have been system-wide:** while more deprived Local Authorities typically spend more and employ more health visiting staff per child, spending reductions have followed similar trajectories across all deprivation groups, limiting the scope for early-years public health services to address regional inequalities.
- Based on 2020 pre-pandemic workforce data scaled to the mid-2024 under-5 population, restoring caseloads to the recommended level of 250 children per practitioner would require hiring **around 3,100 staff** and an estimated additional **£120 million per year in wages**.

## 1. Introduction

There is an established consensus that inequalities in parental investments in the first years of life are a major determinant of inequalities in child health and development (Conti, 2020). Home visiting programmes are a commonly used policy tool to enhance parenting skills and promote a healthy home environment as a strategy to address these disparities (Duffee et al., 2017). Existing evidence solidly shows the effectiveness of home visiting in providing crucial support to parents and children, leading to meaningful short- and long-term benefits for both (Conti et al., 2024b,a; Hjort et al., 2017; Bhalotra et al., 2017).

In recognition of this, both the Department for Education (DfE) and the Department of Health and Social Care (DHSC) have recently placed early years and community-based prevention at the heart of their policy agendas. The NHS 10-Year Health Plan outlines a strategic shift towards prevention and community-based care, with strengthened health visiting services identified as a key priority (Department of Health and Social Care, 2025). This renewed focus is also reflected in the recently announced Best Start Family Hubs, the next phase of the Family Hubs initiative, which aim to integrate early years and family support services locally (Department for Education, 2025). Delivering these ambitions will require rebuilding the public health workforce and a better understanding of local service capacity and variation.

#### What is health visiting?

Health visiting is the core statutory service delivering universal home visiting support for families with children under five in England. Health Visitors (HVs) are registered nurses or midwives who undertake specialist training in community public health nursing – which qualifies them as HVs under the Specialist Community Public Health Nursing (SCPHN) part of the Nursing and Midwifery Council (NMC) Register – and lead the 0-5 element of the Healthy Child Programme (HCP). Health visiting teams are composed of both HVs and Clinical Skill Mix Staff (CSMS), i.e. professionals such as community nursery nurses and staff nurses who are not registered Health Visitors.

Health visiting teams are responsible for delivering a combination of universal and targeted support structured around five mandated health reviews, as well as six high impact areas (Institute of Health Visiting, 2016; UK Government, 2018, 2021). Figure 1 presents a summarised timeline of the history of health visiting in England, with further details available in Appendix A.^1^

**Figure 1:**
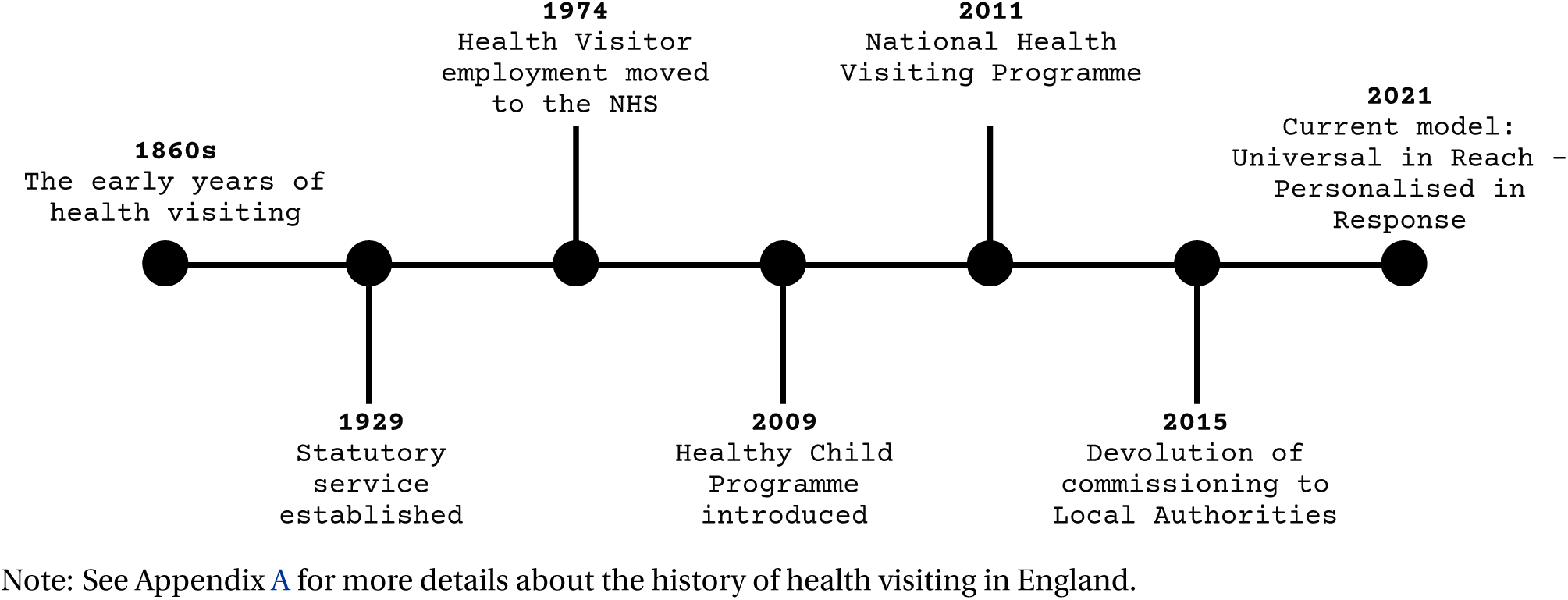
Timeline of Key Events in the History of Health Visiting in England. Note: See Appendix A for more details about the history of health visiting in England.

A major obstacle to studying health visiting in England has been the persistent lack of detailed, standardised data on its workforce. This challenge has become especially urgent since the 2015 devolution of 0-5 public health commissioning to Local Authorities, which enabled services to be outsourced not only to NHS Trusts but also to independent providers or local councils. While NHS Digital publishes monthly workforce statistics,^2^ these only report Health Visitors employed by NHS Trusts, either in aggregate at the national level or in combined figures with other nursing staff at the Trust or Integrated Care Board (ICB) level. They exclude CSMS, student HVs, and all staff employed by non-NHS providers. Experimental data for independent providers^3^ are less frequently published and also lack Local Authority granularity; additionally, reporting by non-NHS providers is entirely voluntary and inconsistent.^4^ Moreover, while both NHS and independent providers often serve multiple LAs, still staff numbers are not available disaggregated at LA level. A promising alternative could be the Community Services Dataset (CSDS), which offers patient-level contact data linked to geographical information^5^; however, the data suffers from patchy coverage, and as such it is unreliable for workforce planning (Clery et al., 2024). These limitations are reflective of a broader lack of standardisation in caseload management and recording across providers and localities (Reid and Tracey, 2023).

This fragmented data landscape is particularly problematic given the substantial decline in the NHS health visiting workforce since 2015 (Institute of Health Visiting, 2022). As shown in Figure 2, the number of Full-Time Equivalent (FTE) HVs employed in NHS Trusts fell from 10,213 in January 2016 to just 5,654 in August 2024, the lowest number recorded since the data series began in 2009 (NHS Digital, n.d.b). This decline coincided with repeated cuts to the public health grant that finances the delivery of public health services, and a shift in commissioning away from NHS Trusts toward independent and council-run provision.^6^ NHS figures thus capture only a partial picture: CSMS are excluded entirely, and the relative stability of HV numbers in independent providers (e.g., 924 FTE in March 2016 vs. 998 in September 2022) is not sufficient to counterbalance the overall workforce loss. From the 2011 National Health Visiting Program (NHVP), involving an influx of 4,200 new FTE Health Visitors, to the devolution of health visiting services from the NHS to Upper Tier Local Authorities (which we refer to as Local Authorities or LAs throughout this report) in 2015, as well as the subsequent cuts to the public health grant during the 2017-2020 period, health visiting has been particularly exposed to vast changes in political landscapes (Department of Health, 2009; Gulland, 2017; Department of Health, 2018; Institute of Health Visiting, 2019, 2022, 2025; NHS Digital, n.d.b). Despite the long-standing statutory role of health visiting, the absence of robust, comprehensive data continues to prevent any systematic evaluation of its effectiveness. This lack of evidence risks undermining the case for long-term support and investment, particularly in the context of renewed policy emphasis on early intervention and community-based prevention.

**Figure 2:**
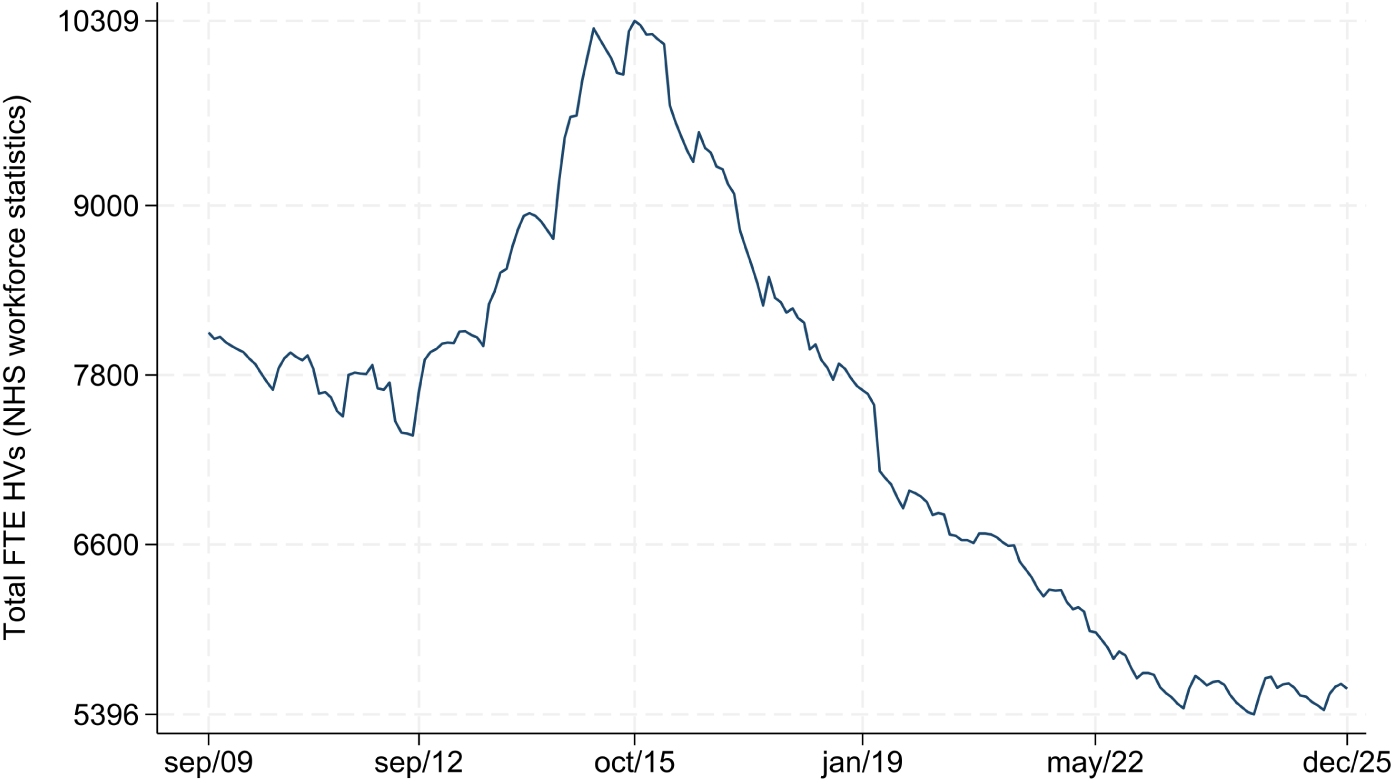
Full-Time Equivalent Health Visitors employed by NHS, 2009-2025. Note: Total number of Full-Time Equivalent Health Visitors employed by NHS, sourced from the NHS workforce statistics.

#### New Freedom of Information data

To overcome these data limitations, we have collected data under the Freedom of Information (FOI) Act, by sending FOI requests to Local Authorities and community providers to obtain detailed figures on health visiting teams from 2016 to 2021. Our FOI dataset captures FTE numbers for both HVs and CSMS, broken down by year, pay band, caseload status, and geographical coverage. Crucially, our data enables analysis at the Local Authority level, offering a more granular and comprehensive view of the health visiting workforce than has been available to date.

## 2. Data

We collected data on the health visiting workforce using the Freedom of Information Act, sending requests to LAs and service providers in England. To ensure consistency across responses, we based classifications on the NHS Occupation Code Manual. The FOI requests were sent in multiple waves beginning in October 2019, with the most recent round conducted between May and December 2022. Additional follow-ups were used to clarify anomalous figures (e.g., implausibly low pay bands^7^ or incomplete breakdowns by caseload status). For each year of data, we use the most recent and most complete response available. During the pandemic period, we also collected information on staff redeployment.^8^

#### What information was collected?

Through FOI requests, we collected annual figures on Full-Time Equivalent (FTE) numbers disaggregated by pay band, caseload-holding status, and staff type – distinguishing between registered Health Visitors and Clinical Skill Mix Staff (i.e. staff working in health visiting teams who are not registered Health Visitors). The data spans from 2016 to 2021, relative to the situation on 1^st^ February each year.

Figure 3 provides an overview of data completeness and provider composition by year and sample.^9^ Among the 147 LAs we contacted,^10^ the number of complete responses increased over time, from 81 LAs in 2016 to 109 in 2021. Among these, 75 provided complete data on the total FTE of both HVs and CSMS across all six years, 66 of which also reported FTE numbers by caseload-holding status for each year.^11^ We refer to these two groups as the *complete total FTE* and *complete caseload FTE* samples, respectively.

**Figure 3:**
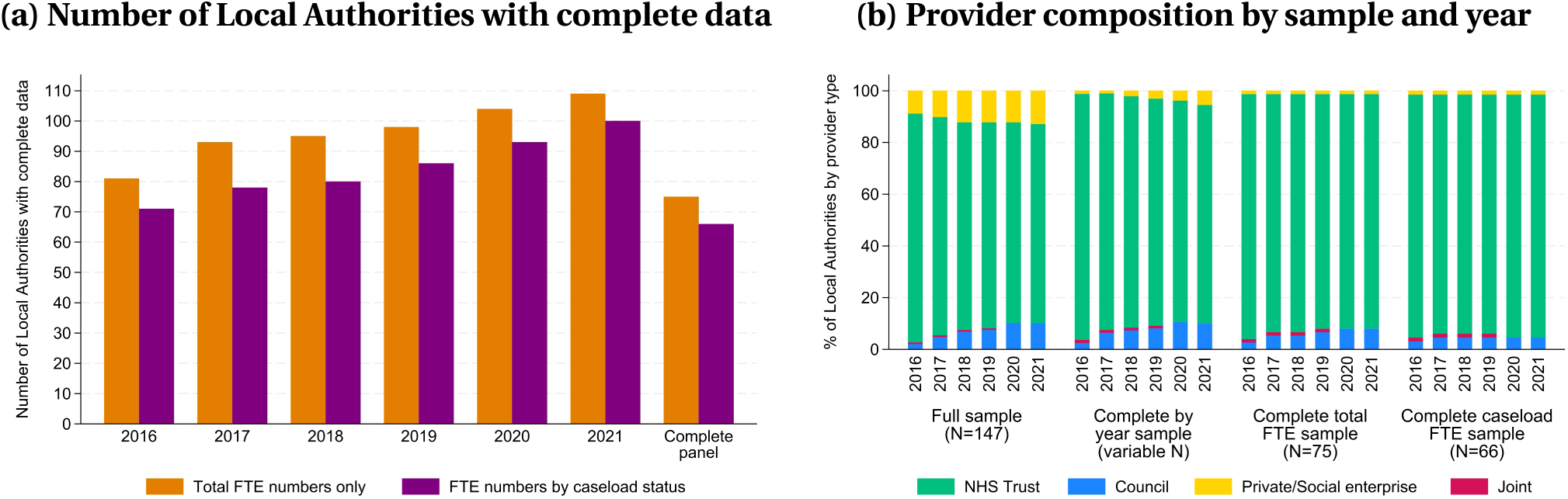
FOI response completeness and provider structure across Local Authorities. Note: In the left panel, “Total FTE numbers only” refers to LAs that provided complete annual data on the total number of FTE health visiting staff, regardless of caseload status. “FTE numbers by caseload status” refers to LAs that additionally reported FTE figures separately for caseload-holding and non-caseload staff. In the right panel, sample definitions are as follows: the “Full sample” includes all 147 LAs; the “Complete by year” sample includes all LAs with complete data in a given year (N = 81 in 2016, 93 in 2017, 95 in 2018, 98 in 2019, 104 in 2020, and 109 in 2021); the “Complete total FTE” sample includes LAs with non-missing total FTE data for all years (N = 75); and the “Complete caseload FTE” sample includes only those that also reported caseload-holding status for each year (N = 66). “Joint” corresponds to joint provision by the council and a private/social enterprise provider (Suffolk).

We use these complete samples when presenting trends over time to ensure comparability. However, for cross-sectional statistics in any given year, and when comparing the endpoints of our data (2016 and 2021), we use the largest available sample with non-missing data for that specific year or pair of years. We do not impute missing data for the remaining LAs, such that the figures provided in any analysis or statistic refer only to the LAs included in the respective sample, unless otherwise stated. Service providers for excluded Las either did not respond or submitted inconsistent or partial information despite repeated follow-ups.^12^

Figure 3b also illustrates how our analytic samples compare to the full sample in terms of provider type. While most LAs in England commissioned health visiting services from NHS Trusts in 2016 (88.4%), this share declined to 76.9% by 2021, as the use of private/social enterprise and council-run provision expanded.^13^ NHS Trusts remain overrepresented in our complete samples: in 2021, 90.7% of the *complete total FTE* sample and 93.9% of the *complete caseload FTE* sample were served by NHS Trusts, compared to 76.9% in the full sample. Conversely, private/social enterprise providers accounted for 12.9% of the full sample in 2021, but just 1.3% and 1.5% of the *complete total FTE* and *complete caseload FTE* samples, respectively. This likely reflects the fact that private/social enterprise providers are not subject to the FOI Act. For council providers, the difference between the full sample and the *complete total FTE* sample is relatively small (10.2% and 8% in 2021, respectively), but it is more pronounced when comparing the full sample to the *complete caseload FTE* sample (4.6% in 2021), suggesting that council providers were less able to provide information on health visiting staff with caseload.

Despite these differences in provider composition, Table B1 in the Appendix shows that the Local Authorities included in our samples – even in the most restrictive, *complete caseload FTE* sample, with 66 LAs – are not statistically distinguishable from excluded LAs across a wide range of observable characteristics, including deprivation indices, public health funding, demographics, and children’s outcomes. Most differences in means between included and excluded LAs are statistically indistinguishable from zero, and additional tests comparing the distributions of these characteristics (Table B2 in the Appendix) also show no significant differences. This suggests that any provider-driven selection into FOI response is unlikely to compromise the validity of our analysis.

## 3. The health visiting workforce over time

### Fewer Health Visitors, more Skill Mix

Our FOI data reveals not only a sustained decline in the total health visiting workforce – mirroring NHS statistics – but also a marked shift in team composition, with rising reliance on lower-paid staff. Between 2016 and 2021, the combined FTE workforce of Health Visitors and Clinical Skill Mix Staff fell by 6.5%, from 7,633 to 7,136 FTE across the 75 Local Authorities with complete data. However, as shown in Figure 4, this overall decline masks diverging trends between the two groups.^14^

**Figure 4:**
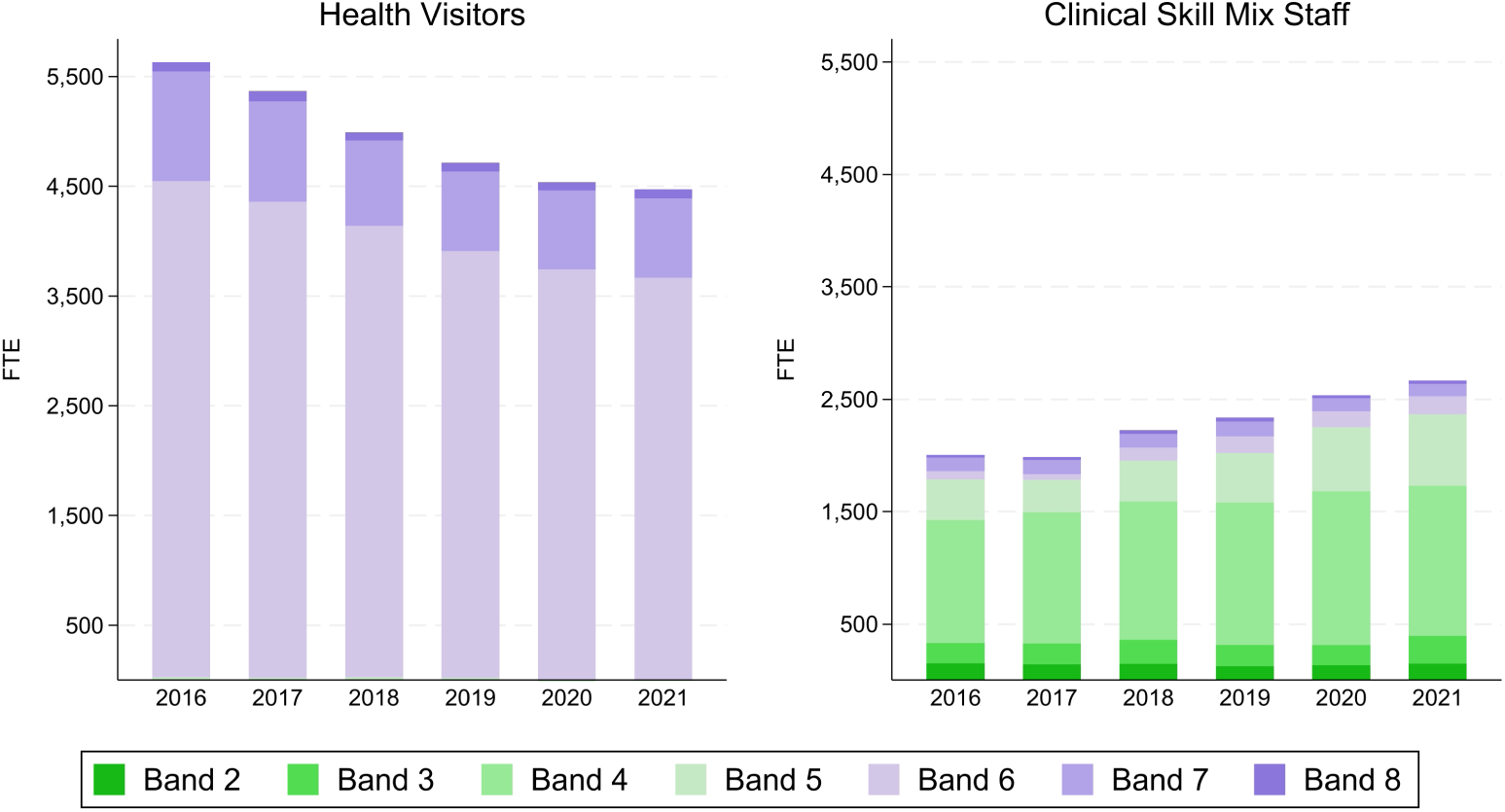
Health visiting workforce composition, 2016-2021. Note: Total Full-Time Equivalent staff in health visiting teams across 75 Local Authorities with complete data. Left panel shows FTE Health Visitors by pay band; right panel shows Clinical Skill Mix Staff by pay band.

The number of FTE HVs fell by 20.6%, from 5,631 in 2016 to 4,472 in 2021.^15^ This decline was driven primarily by reductions in Band 6 HVs – the core of the qualified workforce and the typical minimum grade for qualified HVs under the NHS definition – whose numbers dropped from 4,522 to 3,664 FTE. Band 7 HVs also declined, from 999 to 726 FTE. Band 5 HVs (i.e., student HVs) remained negligible throughout, while Band 8 HVs consistently represented a small but visible share of the workforce. By contrast, the number of CSMS increased by 33%, rising from 2,002 to 2,664 FTE over the same period. This growth was concentrated in Band 4 roles – which rose from 1,093 to 1,336 FTE – and Band 5 roles, which increased from 365 to 635 FTE. Unlike HVs, whose pay bands are concentrated in Bands 6 and 7, the CSMS workforce spans a wider range from Band 2 to Band 8. Nonetheless, Band 4 staff have consistently made up more than half of the CSMS workforce.

Another way to examine these trends is to look at the distribution of the workforce by pay band (Figure 5). Band 6 staff declined from around 60% of the workforce in 2016 to just over 53% in 2021. Over the same period, Bands 4 and 5 combined rose from 19% to 28%, pointing to a shift toward a lower-paid workforce. This reflects a structural transformation in service delivery: a shrinking workforce increasingly reliant on staff with lower pay bands to deliver elements of the Healthy Child Programme.

**Figure 5:**
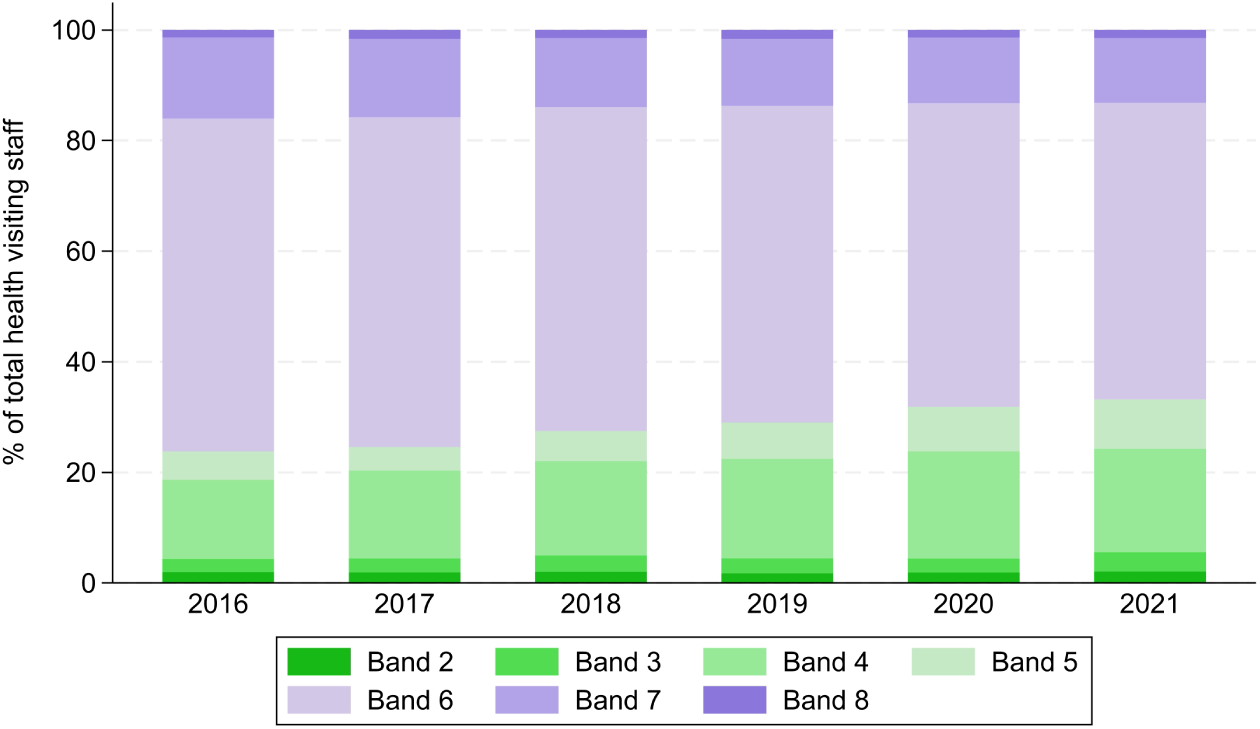
Distribution of health visiting workforce by pay band, 2016-2021. Note: Percentage of total health visiting workforce by Agenda for Change pay band. Based on 75 LAs with complete data.

One plausible contributor is budget pressure following reductions in the public health grant, which may have encouraged greater use of lower-cost staff and altered skill-mix within services. As the Institute of Health Visiting (iHV) stresses: “Community staff nurses, children’s centres workers and community nursery nurses are all highly valued team members, but they do not have this specialist preparation and should not be used as substitute Health Visitors, rather to supplement them by delivering Health Visitor determined interventions in the home” (Institute of Health Visiting, 2018, 2020). Evidence on the effectiveness of replacing HVs with lower-banded staff remains limited, making this a critical open question for workforce planning.^16^

### Declining Health Visitor shares in local health visiting teams

Figure 6 shows the geographical distribution of Health Visitors as a percentage of the total health visiting workforce across English Local Authorities, for each year from 2016 to 2021. These maps use the most complete sample of LAs available for each year. Although Health Visitors continue to make up the majority of the workforce in most areas, there is clear visual evidence of a downward shift in the HV share over time, with fewer LAs shaded in the darkest bands (indicating a share of HVs above 90%) and more in the lighter bands (indicating a lower HV percentage).

**Figure 6:**
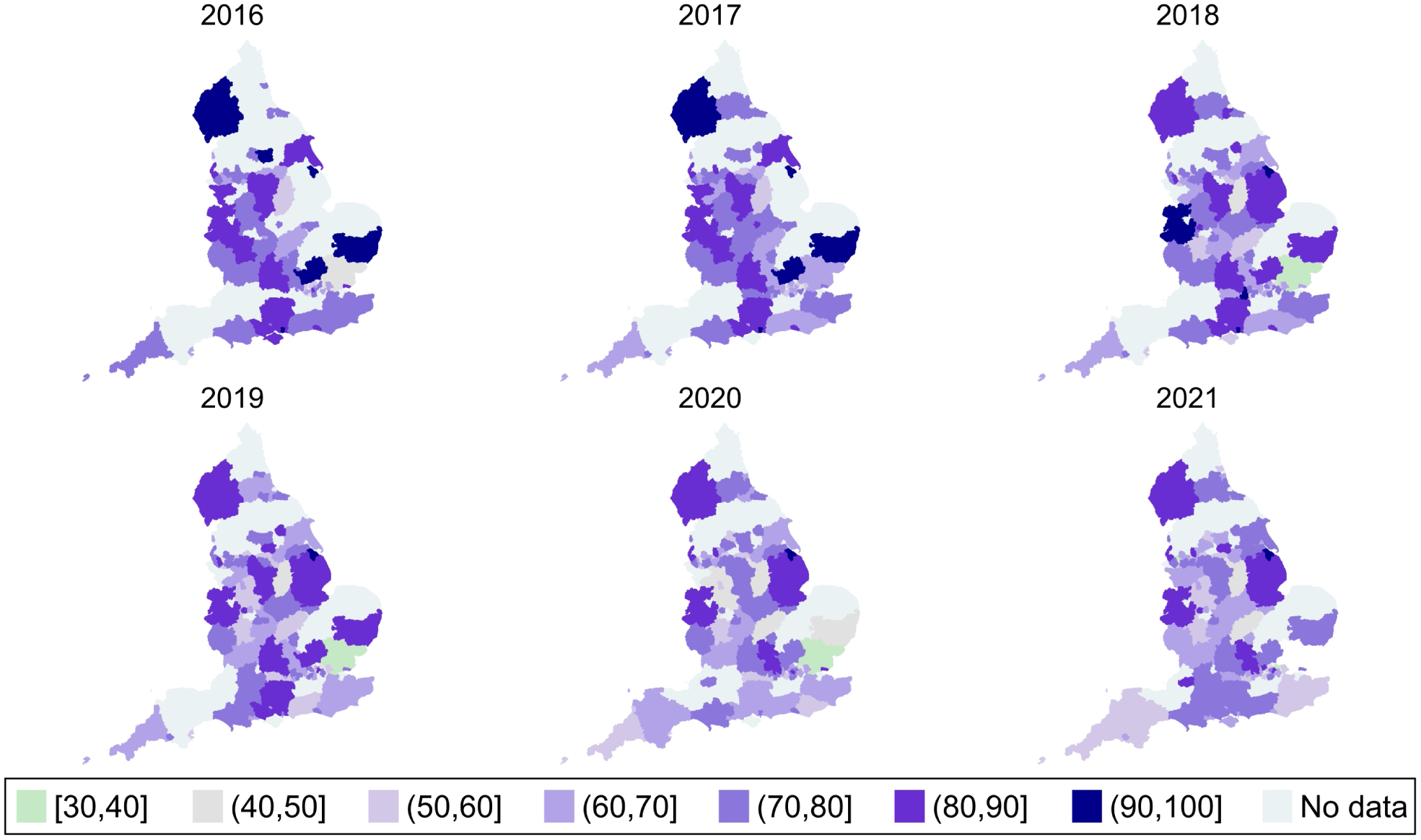
Percentage of Health Visitors in the total workforce, by Local Authority and year. Note: Percentage of Health Visitors in the total health visiting workforce, by Local Authority and year. Darker shading indicates a higher HV share. Based on the most complete sample available in each year (N = 81 in 2016, 93 in 2017, 95 in 2018, 98 in 2019, 104 in 2020, and 109 in 2021).

This compositional shift is also reflected in the data when holding the sample constant: among the 81 LAs with complete data in both 2016 and 2021, 77.8% experienced a decline in the share of Health Visitors within their total workforce. Comparing the cross-sectional distributions at the two endpoints further illustrates this trend. In 2016, nearly half of the LAs had a workforce composed of more than 74% Health Visitors, with only a small number falling below the 60% mark. By 2021, the distribution had shifted: the median share of HVs had declined, and almost a third of LAs had fewer than 60% HVs in their teams. Only 17% of LAs remained above the previous median of 74%. While some areas still maintained a high HV share, they had become the exception rather than the norm.

To further examine how local workforce composition evolved over time, Figure 7 classifies Local Authorities according to the direction of change in the number of Health Visitors and Clinical Skill Mix Staff between 2016 and 2021. This map highlights striking geographical variation in how LAs responded to workforce pressures. While some areas managed to expand both HV and CSMS numbers (8.6% of LAs), and a few increased HVs despite reductions in CSMS (2.5%), these were clear exceptions. The most common pattern, observed in 54.3% of LAs, was a decrease in Health Visitors alongside an increase in CSMS – reinforcing the overall shift toward a more skill-mixed model. A further 32.1% of LAs saw declines in both HV and CSMS staffing, pointing to broader workforce contraction rather than substitution. These local differences highlight how workforce restructuring has unfolded unevenly across the country.

**Figure 7:**
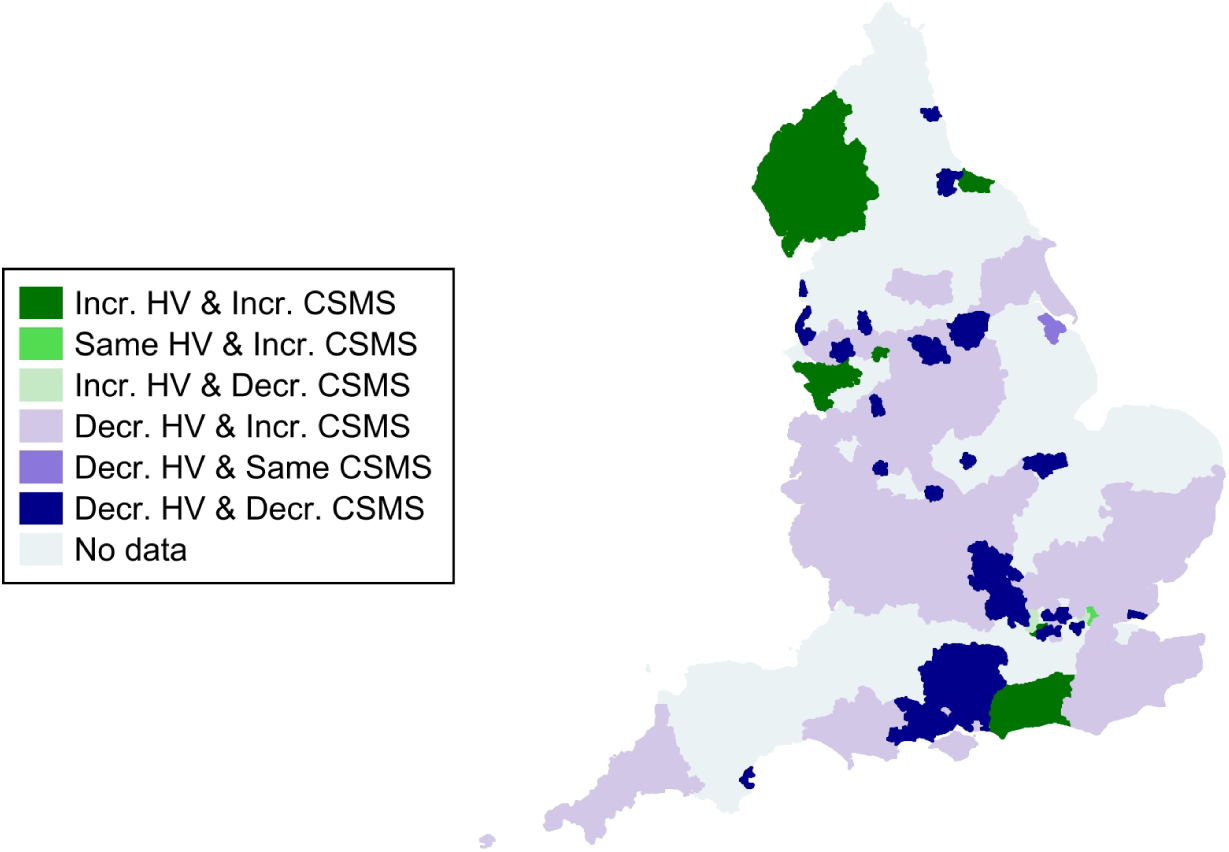
Change in workforce composition, 2016–2021. Note: Classification of Local Authorities by the direction of change in Health Visitor and Clinical Skill Mix Staff numbers between 2016 and 2021. Based on 81 LAs with complete data for both years.

Table B4 in the Appendix reports results from multinomial logit models examining associations between baseline LA characteristics and subsequent changes in the health visiting workforce between 2016 and 2021. The dependent variable is a three-category indicator distinguishing *Expansion* (an increase or no change in Health Visitor FTE alongside an increase in Clinical Skill Mix Staff FTE), *Substitution* (a decline in HV FTE combined with an increase in CSMS FTE), and *Contraction* (declines in both HV and CSMS FTE).

Overall, the results provide limited evidence of systematic relationships between baseline characteristics and subsequent workforce change patterns. The clearest associations are observed for deprivation measured using the Income Deprivation Affecting Children Index (IDACI, IMD 2015) and provider type. Higher IDACI scores are positively associated with the probability of workforce contraction, suggesting that more income-deprived LAs were somewhat more likely to experience reductions in both Health Visitors and CSMS between 2016 and 2021. LAs where health visiting services were provided by NHS Trusts appear more likely to experience workforce expansion and less likely to experience contraction. Beyond these associations, there is weak evidence that higher reception-year obesity prevalence is associated with a lower probability of workforce contraction, although the estimated effect is small in magnitude. Higher spending on mandated health visiting per child is similarly associated with a slightly higher probability of contraction, but this effect is also small. For the remaining covariates, estimated effects are small and not statistically significant.

### Rising caseloads, shrinking capacity

The preceding sections documented how health visiting teams have shifted in size and composition, with widespread reductions in qualified Health Visitors and growing reliance on Clinical Skill Mix Staff. These workforce changes raise important questions about how service delivery is sustained at the local level – particularly in relation to caseloads, a key measure of workforce pressure and capacity. The caseload is the number of children assigned to each practitioner, which the Institute of Health Visiting recommends should not exceed 250 per Health Visitor (Institute of Health Visiting, 2017, 2020).^17^

##### Who holds caseloads?

Despite some variation in how teams are structured, Health Visitors are consistently the majority of the caseload-holding workforce. Across the 66 LAs with complete data on caseload-holding status, around 85% of staff with assigned caseloads were registered HVs throughout the 2016-2021 period. This declined only modestly over time – from 86.7% in 2016 to 83.1% in 2021 –, indicating that HVs continue to carry the bulk of direct responsibility.

However, the composition of the caseload-holding workforce is shifting. As shown in Figure 8, most LAs experienced a drop in the number of caseload-holding Health Visitors between 2016 and 2021, while increases in caseload-holding CSMS were more mixed. Among the 71 LAs with complete data in both years, 85.9% saw a decline in caseload-holding HVs. By contrast, 43.7% of LAs increased their number of caseload-holding CSMS^18^, 29.6% reported no change, and 26.8% experienced a decline. These trends suggest that some teams have adapted to HV shortfalls by assigning more families to CSMS.^19^

**Figure 8:**
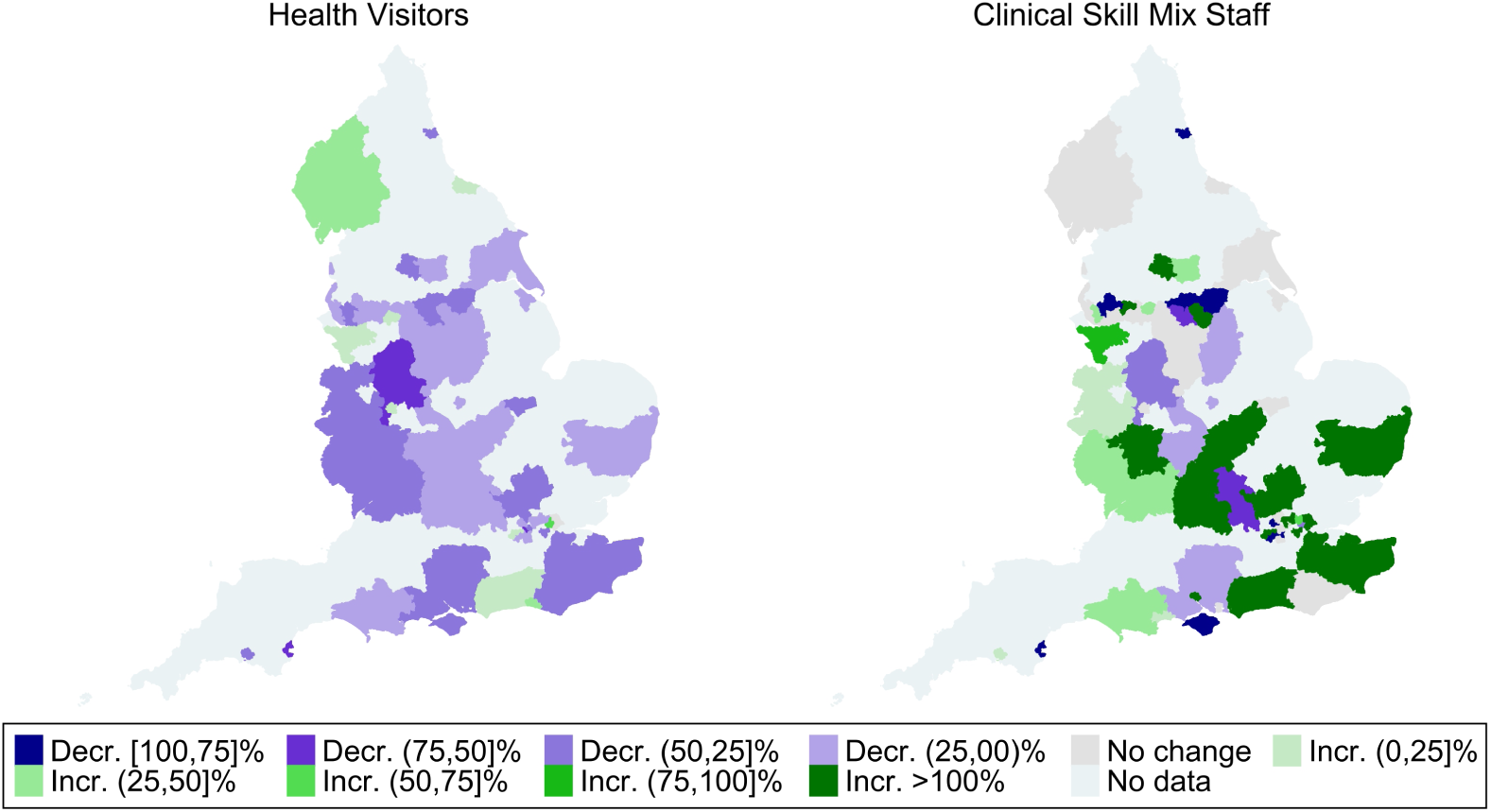
Change in number of caseload-holding HVs and CSMS, 2016–2021. Note: Left panel shows percentage change in the number of caseload-holding Health Visitors; right panel shows percentage change in caseload-holding Clinical Skill Mix Staff. Based on 71 LAs with complete caseload data in both 2016 and 2021. Five LAs (Greenwich, Haringey, Islington, Suffolk, and West Sussex) had an undefined percentage change for CSMS because they started with 0 FTE in 2016; these were included in the ‘Incr. >100%’ category.

The net effect of these workforce changes has been a rise in caseload sizes across much of the country. Figure 9 shows the full distribution of average caseloads by year: over time, the distribution shifts clearly to the right, with more LAs with higher average caseloads. Notably, the tail of the distribution has lengthened, with a growing number of LAs at extremely high average caseloads exceeding 1,000.^20^ While the percentage of LAs with average caseloads exceeding 250 remained relatively stable from 2016 until 2021 (approximately 74%), those above 300 rose steadily from 42.4% in 2016 to a peak of 62.1% in 2020, before falling slightly in 2021. Similarly, the share of LAs exceeding 350 rose from 30.3% in 2016 to 43.9% in 2020, then declined to 34.8% in 2021.^21^

**Figure 9:**
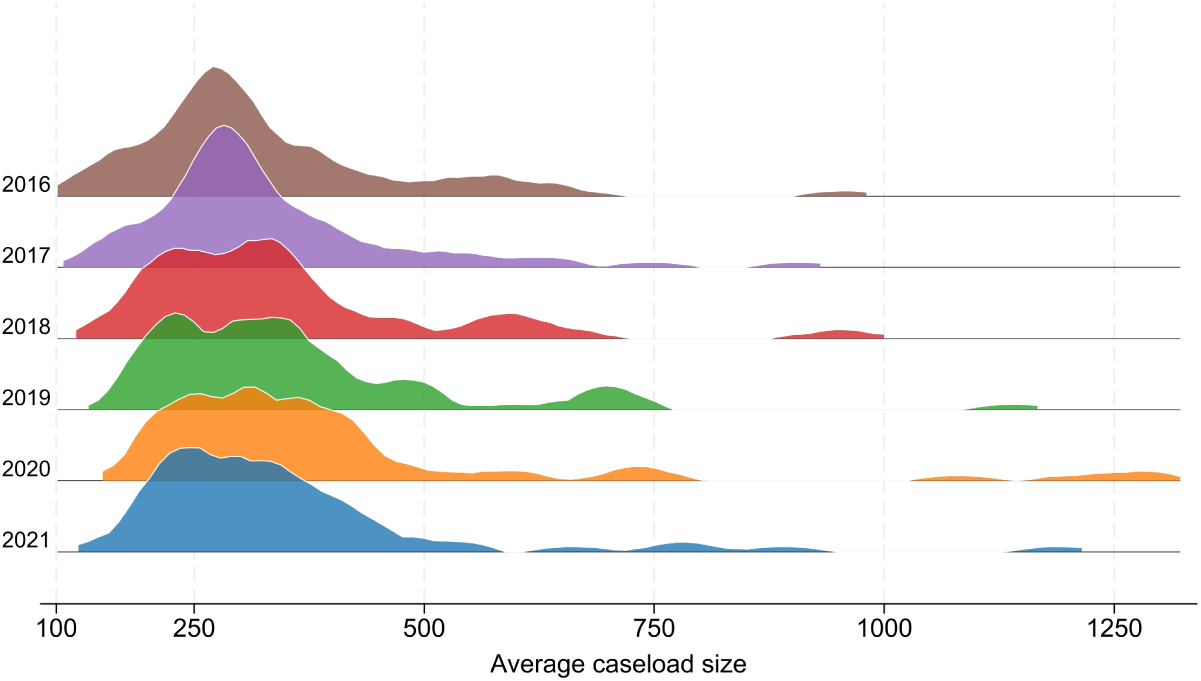
Distribution of average caseload size, 2016–2021. Note: Distribution of average caseload size among caseload-holding staff across LAs, by year. Caseload size is computed as the number of children under 5 divided by the total number of FTE caseload-holding staff (HV + CSMS) working within health visiting teams. Based on 66 LAs with complete data.

### More CSMS hold caseloads where teams use a corporate model

Another important aspect of health visiting is how caseloads are organised. Caseloads are either *individually managed*, when each Health Visitor is responsible for a single group of cases (with possible delegation of some tasks to CSMS members of the health visiting teams), or follow a *corporate model*, when the entire health visiting team (including CSMS) shares a single caseload and families receive services from any team member (Whittaker et al., 2021).^22^ During the period of our data collection, we identified the following Local Authorities as having a corporate or shared caseload model: Kent, Oxfordshire, Rochdale, Staffordshire, Stoke-on-Trent, and Swindon.^23^

Comparing LAs with and without a known corporate caseload model shows significant differences, as shown in Figure 10: in LAs with corporate caseloads, the percentage of CSMS who were caseload holders (63.3%) was larger than in other LAs (41.5%), while the respective percentages of HVs were similar.^24^ Higher percentages of caseload-holding CSMS seem consistent with a model of shared caseload organisation, in which families are provided services by any available team member. CSMS without caseloads may be involved in several other activities, such as participating in health education programmes, providing preceptorship and mentorship to students and other staff members, taking part in clinical supervision, assisting in research and audits, record keeping, attending meetings and conferences.^25^

**Figure 10:**
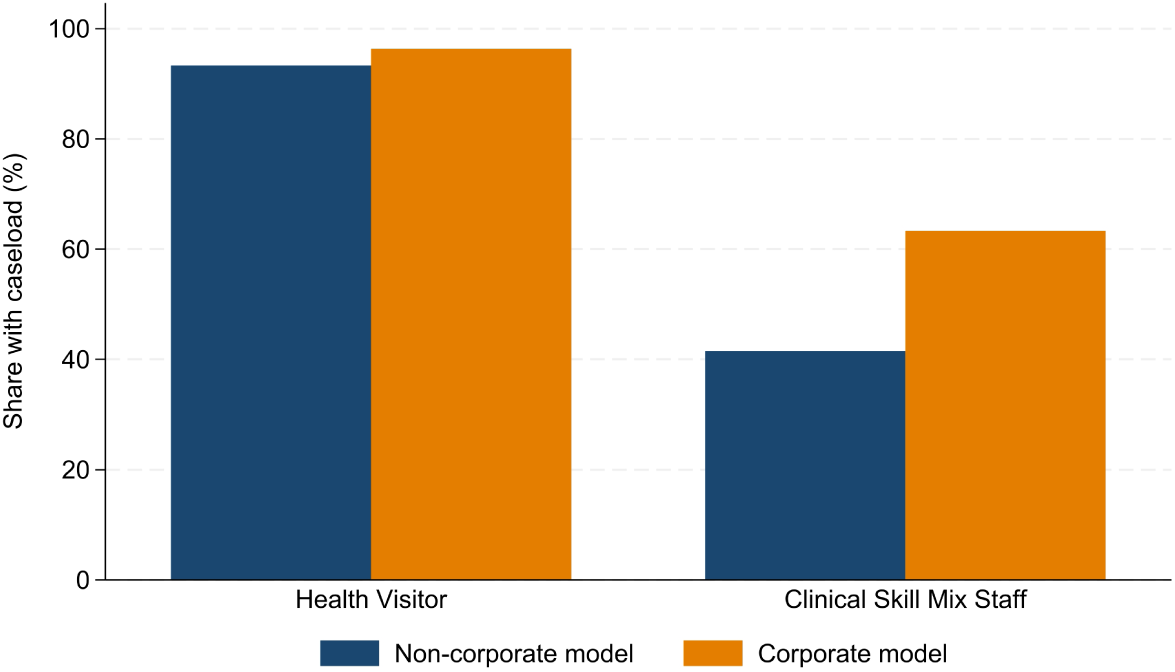
Percentage of staff with direct caseload responsibility, by caseload model. Note: Share of Health Visitors and Clinical Skill Mix Staff holding caseloads, by caseload organisation model (corporate vs. non-corporate). Based on averages across all years in the 66 LAs with complete data.

## 4. Redeployment of health visiting staff during COVID-19

The COVID-19 pandemic occurred within the 2016-2021 period covered by our analysis, introducing major disruptions to community and early years services. During this time, Health Visitors and members of health visiting teams were redeployed to support the wider NHS response, reducing capacity for routine contacts and statutory home visits. To document these effects, our FOI data collection was extended to include information on redeployments during the first and second pandemic waves, providing a local record of how services were affected across England.

### Mandated contacts were suspended under emergency guidance

In England, just prior to the first national lockdown, NHS England and NHS Improvement issued guidance redirecting staff to support the emergency COVID-19 response. Published on 17 March 2020, this letter called for registered nurses in non-patient-facing roles – including Health Visitors and nurses in health visiting teams – to be redeployed to front-line clinical care (NHS England, 2020c). Two days later, further guidance introduced a COVID-19 prioritisation plan for community health services (NHS England, 2020a), pausing routine pre-birth and 0-5 services except for antenatal (virtual) and new birth (virtual or face-to-face) contacts. Other mandated visits – at 6-8 weeks, 1 year, and 2-2.5 years – were to be delivered only for families identified as vulnerable or with clinical need.

This emergency reprioritisation effectively suspended three of the five mandated Healthy Child Programme contacts for most families. However, guidance was revised later in the year, on 3 June 2020, and health visiting services were restored (NHS England, 2020b). On 7 October 2020, a joint winter planning letter from Public Health England, NHS England, and the Local Government Association advised that “professionals supporting children and families, such as health visitors, […] should not be redeployed to other services” (Local Government Association, 2020). The letter called for continued front-line provision for pregnant women and families with young children, while allowing short redeployments only where staff had specialist skills needed locally (e.g., Intensive Therapy Unit training).^26^

To capture the scale and timing of redeployment, we extended our FOI data collection to cover the pandemic period. In England, data for the first wave was collected between 27 August 2020 and 26 January 2021; data for the second wave was collected between 1 April and 26 October 2021. We asked Local Authorities and providers to report the maximum number of FTE Health Visitors and Clinical Skill Mix Staff redeployed between 1 September 2020 and 31 March 2021.^27^

For more detailed analysis, we divided the second wave into three sub-periods: 1 September-7 October 2020 (prior to updated guidance), 8 October-31 December 2020, and 1 January-31 March 2021. Respondents were also asked to report the start and end dates of redeployment episodes. In cases where redeployment occurred in multiple phases, these refer to the earliest departure and latest return of any team member.

### Substantial redeployment in the first COVID-19 wave

Figure 11 presents redeployment levels across Local Authorities during the first wave of the COVID-19 pandemic, expressed as a percentage of the local health visiting workforce. The left panel shows redeployment of Health Visitors, while the right panel shows redeployment of Clinical Skill Mix Staff. Many areas reported no redeployment (shaded in green), but others experienced substantial losses, particularly among CSMS.

**Figure 11:**
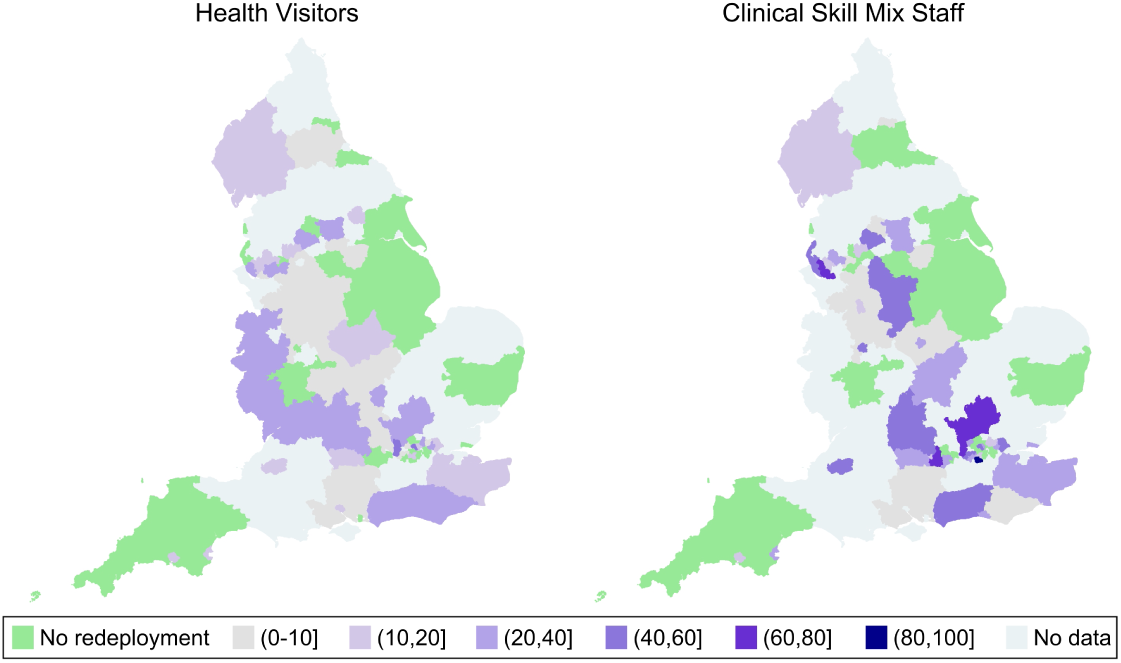
Percentage of health visiting staff redeployed during the first COVID-19 wave. Note: Percentage of local Health Visitors (left) and Clinical Skill Mix Staff (right) redeployed during the first wave of COVID-19. Sample restricted to 103 LAs with complete FTE data in both 2020 and 2021. Data on HV redeployment available for 99 LAs; data on CSMS redeployment for 90 LAs.

These patterns are based on 103 LAs with complete FTE data in both 2020 and 2021. Among them, 99 LAs provided data on HV redeployment, and 90 on CSMS. Overall, 68.7% of LAs redeployed at least one staff member from their health visiting team; 57.6% redeployed at least one HV, and 57.8% redeployed at least one CSMS.

On average, 9.1% of HVs and 17.8% of CSMS were redeployed during this period. However, the intensity of redeployment varied widely. In the most affected LAs, up to 45% of HVs and 83.3% of CSMS were reassigned. High-intensity redeployment was more common among CSMS: 12.2% of LAs redeployed over half of their CSMS workforce, while 10.1% redeployed more than a quarter of their HVs.

Health visiting staff began to be redeployed immediately following the issuance of NHS guidance on 19 March 2020. In 91.2% of Local Authorities that redeployed staff and provided start dates (62 out of 68), redeployment began before May. Among the 50 LAs that reported both start and end dates, the average duration was 82.4 days, highlighting the sustained disruption to service capacity during the first wave. Notably, redeployments often extended well beyond the official restoration of health visiting services on 3 June 2020. In 71.9% of LAs with end-date data (46 out of 64), staff remained redeployed after this date.

### Redeployment was more limited during the second wave

Figure 12 presents redeployment levels across three phases of the second COVID-19 wave, disaggregated by staff type. The top panel shows redeployment of Health Visitors; the bottom panel shows redeployment of Clinical Skill Mix Staff. Compared to the first wave, redeployments during this period were considerably less widespread and generally of lower intensity.

**Figure 12:**
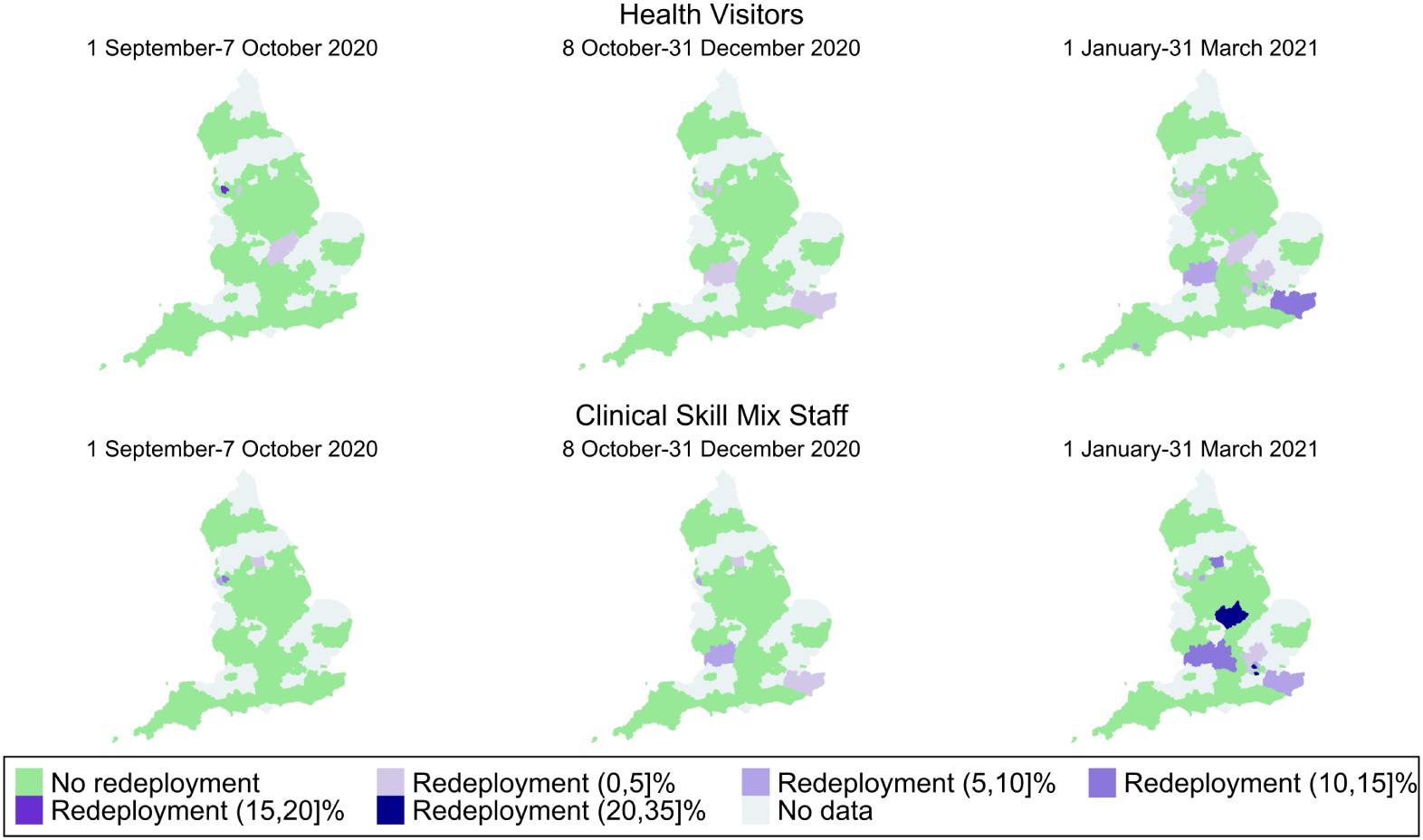
Percentage of health visiting staff redeployed during the second COVID-19 wave. Note: Percentage of local Health Visitors (top row) and Clinical Skill Mix Staff (bottom row) redeployed in each phase of the second wave: 1 September-7 October 2020; 8 October-31 December 2020; and 1 January-31 March 2021. Data on HV redeployment available for 93 LAs; data on CSMS redeployment available for 92 LAs.

Across the entire period from 1 September 2020 to 31 March 2021, only 25 LAs reported redeploying any health visiting staff. Among the 93 LAs with data on HV redeployment, just 17.2% (16 LAs) reported redeploying at least one HV. Similarly, among the 92 LAs with data on CSMS, 18.5% (17 LAs) reported at least one CSMS redeployed. Few LAs reported redeployments during the first two periods analysed: five had redeployments in place between 1 September and 7 October 2020, and eight between 8 October and 31 December 2020. In the final period (January-March 2021), the number of LAs redeploying at least one staff member went up to 23, with redeploying LAs having reassigned an average of 3.8% of HVs and 11.3% of CSMS, and some areas redeploying up to 10.7% of HVs and over 30% of CSMS.

These patterns suggest that although redeployment continued into the second wave, it was more contained, both geographically and in terms of workforce disruption, when compared to the first wave.

## 5. Public spending on health visiting

### Spending on mandated 0-5 services faced sustained reductions

Since October 2015, the responsibility for commissioning health visiting services has been transferred to Local Authorities, predominantly funded by the public health grant – a ring-fenced allocation from central government intended to support a range of local public health functions. Within early years provision, Local Authorities are required to deliver a core set of prescribed functions (mandated services), while other elements of the Healthy Child Programme are considered non-prescribed and therefore non-mandated.

Spending on these two categories is reported separately in the Revenue Outturn (RO3) dataset.^28^ Line 84 covers 0-5 children’s services (prescribed functions), which include the five mandated universal health reviews: the antenatal, new baby, 6-8 week, 1 year, and 2-2.5 year reviews. Line 85 captures other 0-5 children’s services (non-prescribed functions), which encompass the broader Healthy Child Programme. These include services beyond the five mandated reviews – targeted support, community-based activities, and the Family Nurse Partnership.

Figure 13 shows national trends in Local Authority spending per child under five, in real terms (2023-24 prices), for both budget lines between 2016-17 and 2024-25. Population figures are based on the Office for National Statistics mid-year estimates.

**Figure 13:**
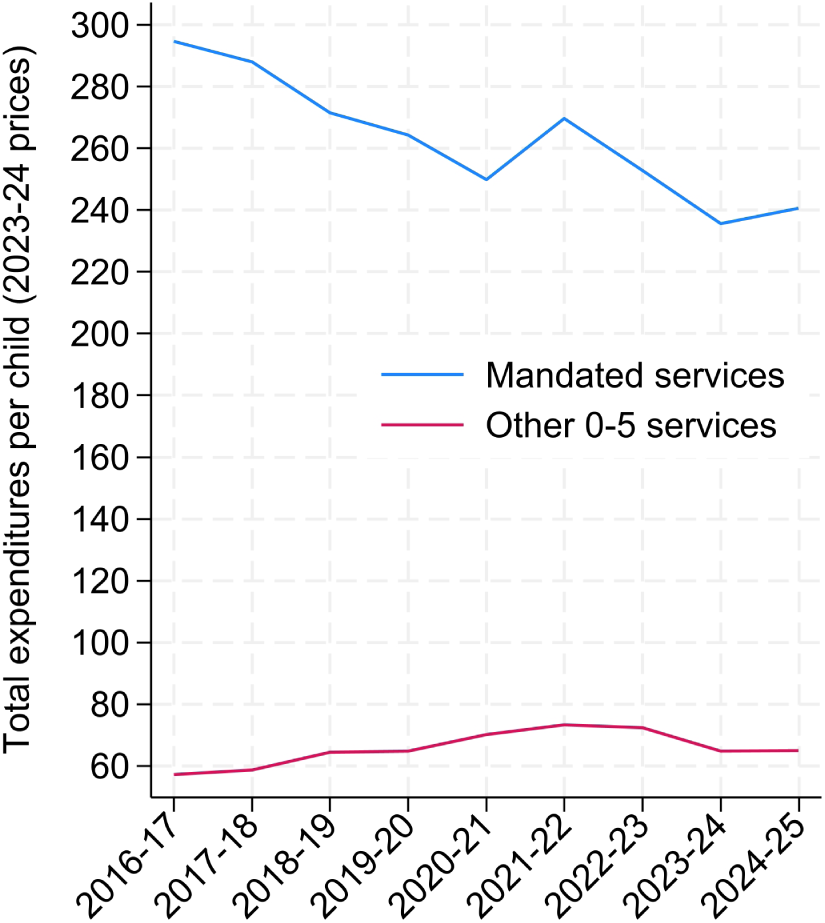
Spending per child on mandated and non-mandated 0-5 services, 2016-17 to 2024-25. Note: Real-terms spending per child under five (in 2023-24 GBP) on mandated 0-5 children’s services (prescribed functions; Line 84) and non-mandated 0-5 children’s services (non-prescribed functions; Line 85). Figures are based on Local Authority revenue out-turn (RO3) total expenditures and ONS mid-year population estimates, including all LAs with non-missing RO3 data (N = 146 in 2016-17, 147 in 2017-18 and 2018-19, 149 in 2019-20, 150 in 2020-21, 152 in 2021-22 and 2022-23, 148 in 2023-24, and 147 in 2024-25). Values adjusted using the GDP deflator.

Spending on mandated services has fallen consistently over time, from £294.7 per child in 2016-17 to £240.7 in 2024-25, a reduction of nearly 20% in real terms. There was a temporary rebound in 2021-22 (to £269.7), but this was short-lived, and spending resumed its down-ward trajectory thereafter. By contrast, spending on non-mandated services rose from £57.1 per child in 2016-17 to £64.9 in 2024-25, an overall increase of 13.6% in real terms, though with a peak of £73.2 in 2021-22. Importantly, however, non-mandated spending is significantly smaller in scale: total expenditure on non-prescribed 0-5 services has consistently amounted to less than one third of that on mandated health visiting across all years. Overall, these trends show that Local Authorities have faced sustained real-terms cuts to statutory health visiting provision, alongside more volatile investment in discretionary early years services.

Beyond these national averages, the dispersion of spending across Local Authorities reveals distinct patterns (Figure B3 in the Appendix). Variation in mandated service spending remained stable at around 31-35% between 2016-17 and 2020-21, before rising sharply to over 40% in 2021-22 and remaining elevated thereafter, suggesting increasing divergence in how LAs funded statutory provision during the post-pandemic recovery period (although the most recent data point shows a modest decline back to 36.6%). In contrast, non-mandated spending was initially far more uneven: its coefficient of variation exceeded 120% in 2016-17 but declined steadily to around 94% by 2024-25. This indicates that discretionary early-years investment, while still highly unequal, has become more uniform over time.

### Investment needed to hire health visiting staff

So far, we have documented a sustained decline in the health visiting workforce alongside reductions in Local Authority spending on mandated services. We now estimate the investment required to hire new Health Visitors (HVs) and Clinical Skill Mix Staff (CSMS).

We estimate annual and one-off costs of workforce expansion using the 2024/25 NHS Agenda for Change pay scales.^29^ For HVs, we include one year of training costs covering tuition fees, a year of Band 5 salary for student HVs, and 20% of a Band 7 practice teachers time supervising three students during their clinical placement. This training year is followed by ongoing annual wages at Band 6. For CSMS, costs are based solely on annual wages at Band 4, as no formal training costs are assumed. All wages are uplifted to reflect non-wage costs. Table 1 summarises the data sources and parameters used in these calculations.^30^

**Table 1:** Data sources and parameters used for cost calculations.

| Item | Source | Parameter used |
| --- | --- | --- |
| CSMS annual salary (band 4, top) | NHS Pay Scales, AfC 2024/25 | £29,114 |
| Student HV salary (band 5, mid-point) | NHS Pay Scales, AfC 2024/25 | £32,324 |
| HV annual salary (band 6, mid-point) | NHS Pay Scales, AfC 2024/25 | £39,405 |
| Practice teacher salary (band 7, mid-point) | NHS Pay Scales, AfC 2024/25 | £48,526 |
| Non-wage costs per hour | ONS Index of Labour Costs, 2020 Q3 | £2.40 |
| Average labour compensation per hour worked | ONS Labour Costs per Hour, 2020 Q3 | £21.65 |
| Wage uplift for non-wage costs | Calculated (21.65 / (21.65 – 2.4)) | 1.12 |
| Number of students per teacher | Confidential HV personnel | 3 |
| Fraction of student time in clinical practice | SCPHN degree course descriptions | 0.5 |
| Fraction of teacher time assessing students | Confidential HV personnel | 0.2 |
| Tuition cost per student | Institute For Apprenticeships And Technical Education | £14,000 |

Note: AfC = Agenda for change. SCPHN = Specialist Community Public Health Nursing. NMC = Nursing and Midwifery Council. The maximum funding for the SCPHN (NMC 2022) apprenticeship is reported on the Institute For Apprenticeships And Technical Education website: https://skillsengland.education.gov.uk/apprenticeship-standards/st1418-v1-1. We used the latest available version of index of labour costs (Health and Social Work), published in 2020 by the ONS, and the average labour costs (Human health and social work activities) for the same period for comparability, to calculate the wage uplift for non-wage costs. According to the new HV training systems, the tasks previously performed by Community Practice Teachers have been split between Practice Assessors (PAs, at pay band 7) and Practice Supervisors (PSs, at pay band 6). In our calculations, teachers are assumed to be only “Practice Assessors”.

Table 2 presents the estimated costs of hiring 1,000, 3,000, and 5,000 additional HVs and CSMS. Each new HV is estimated to cost approximately £44,319 per year in wages (including non-wage costs) and £52,174 in one-off training expenses, while each CSMS costs around £32,745 per year. A pledge to recruit 3,000 new Health Visitors would therefore require around £133 million annually once fully trained, plus approximately £157 million in one-off training costs.

**Table 2:**
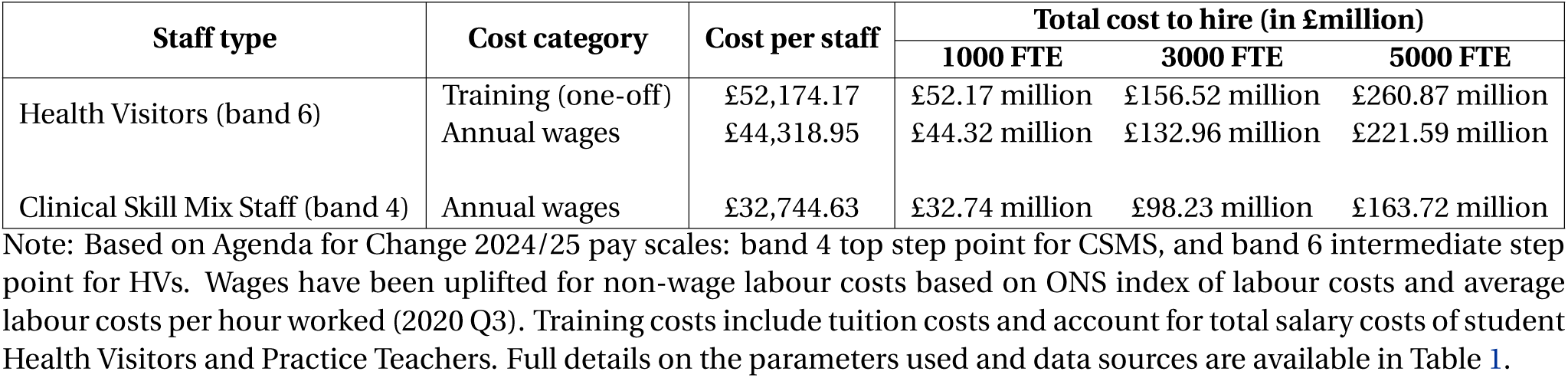
Estimated annual cost of hiring additional Health Visitors and Clinical Skill Mix Staff.

| Staff type | Cost category | Cost per staff | Total cost to hire (in £million) |  |  |
| --- | --- | --- | --- | --- | --- |
|  |  |  | 1000 FTE | 3000 FTE | 5000 FTE |
| Health Visitors (band 6) | Training (one-off) | £52,174.17 | £52.17 million | £156.52 million | £260.87 million |
|  | Annual wages | £44,318.95 | £44.32 million | £132.96 million | £221.59 million |
| Clinical Skill Mix Staff (band 4) | Annual wages | £32,744.63 | £32.74 million | £98.23 million | £163.72 million |

Note: Based on Agenda for Change 2024/25 pay scales: band 4 top step point for CSMS, and band 6 intermediate step point for HVs. Wages have been uplifted for non-wage labour costs based on ONS index of labour costs and average labour costs per hour worked (2020 Q3). Training costs include tuition costs and account for total salary costs of student Health Visitors and Practice Teachers. Full details on the parameters used and data sources are available in Table 1.

### Investment needed to reduce caseloads

To assess the scale of investment required to restore service capacity, we estimate the workforce and financial resources needed to reduce average caseload sizes using FOI data from 2020, the last pre-pandemic year for which we have workforce information.

Our approach proceeds in two steps. First, we calculated the additional FTE staff required for each Local Authority to reach target caseload thresholds of 200, 250, 300, and 350 children per practitioner. Caseloads were derived as the ratio of children under five (ONS mid-year population estimates) to the number of caseload-holding HVs and CSMS reported in FOI data. We then compared observed caseloads with each threshold and scaled the results to the national under-5 population in mid-2024. Second, we translated the estimated workforce shortfall into cost projections under three staffing scenarios: (i) all new staff as HVs (band 6), (ii) a 50:50 mix of HVs (band 6) and CSMS (band 4), and (iii) all new staff as CSMS (band 4). The resulting estimates are shown in Table 3.

**Table 3:**
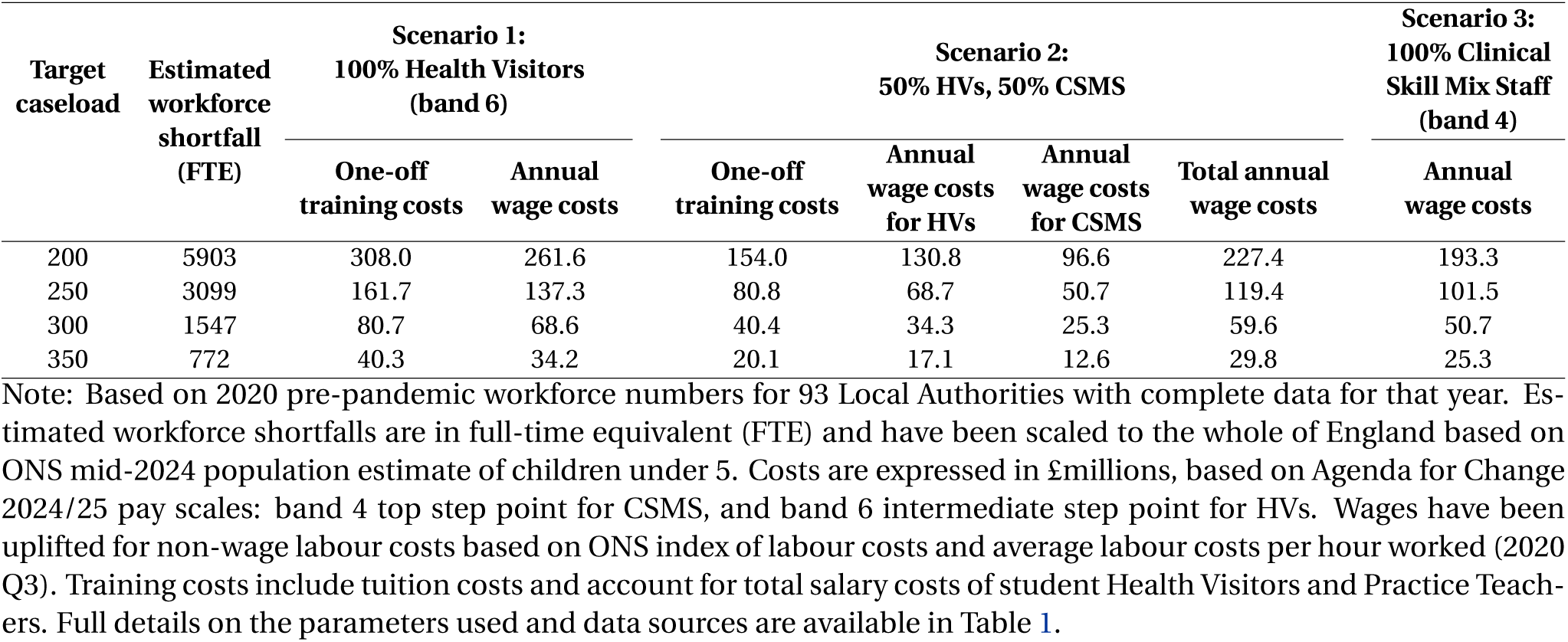
Estimated workforce shortfall and costs of meeting alternative caseload targets (in £million), by workforce mix scenario.

Meeting an average caseload of 250 children per practitioner – the Institute of Health Visiting’s recommended maximum – would require around 3,100 additional FTE staff. Depending on the staffing mix, this equates to one-off training costs between £81 million (50:50 mix) and £162 million (all HVs), and annual wage costs between £102 million (all CSMS) and £137 million (all HVs). Relaxing the target to 350 children per practitioner would still imply a shortfall of around 770 FTE, with wage costs between £25 million (all CSMS) and £34 million (all HVs).

Taken together, these estimates indicate that restoring safe caseloads should be a priority for early years investment and is financially achievable within the scale of recent early years commitments. The Family Hubs and Start for Life programme allocated about £330.45 million in 2022-25, plus £126 million in 2025-26, and the newly announced Best Start Family Hubs expansion commits over £500 million across three years (roughly £170 million per year to 2028). By comparison, returning to an average caseload of 250 children per practitioner would require around £120 million per year in additional wages under a 50:50 HV-CSMS mix, plus about £80 million in one-off training costs. Prioritising health visiting workforce rebuilding would align with these funding allocations and provide the capacity needed to advance the government’s ambitions on prevention and services delivered in the community, helping new investments such as the Best Start Family Hubs expansion to achieve their intended impact.

## 6. Inequalities by baseline deprivation

The preceding section document large differences across Local Authorities in the size and composition of health visiting teams, caseload pressures, and spending on 0-5 public health services. A central question is then whether these patterns map onto underlying disadvantage – i.e., whether the areas facing the greatest early-years need also experienced systematically different levels and trajectories of investment and service delivery over time.

To examine this, we present a set of analyses stratified by baseline deprivation, using the 2015 Income Deprivation Affecting Children Index (IDACI). We rank Local Authorities by their average IDACI score and assign them to tertiles: (i) least deprived, (ii) middle, and (iii) most deprived. We then compare both levels and trends in a set of outcomes spanning (1) local public spending on 0-5 services, (2) workforce numbers, and (3) health visiting service delivery metrics.

Specifically, for each Local Authority-year, we analyse: expenditure on mandated and non-mandated 0-5 services, measured both in total and per child under five (in real £); workforce inputs measured as FTE Health Visitors and Clinical Skill Mix Staff, again both in levels and per 1,000 children under five; and service delivery metrics capturing completion of mandated reviews (New Birth Visits, 6-8 week reviews, 12-month reviews, 2-2.5 year reviews, and 2-2.5 year reviews completed with ASQ-3).

We proceed in three steps. First, we plot mean outcomes for each IDACI tertile over 2016-2021. Second, we test for cross-sectional differences in levels within each year (using both non-parametric and parametric approaches, supplemented by pairwise comparisons). Third, we estimate Local Authority fixed effects regressions with Year × IDACI interactions, to assess whether trends differ by baseline deprivation. We report results under two specifications: a flexible model with categorical year interactions and a more parsimonious model with linear trend interactions.

### Spending on 0-5 services: persistent per-child gradients, similar trends over time

Figure 14 shows clear and contrasting deprivation gradients in spending on 0-5 services. In levels (left panel), mean expenditure on mandated health visiting is consistently highest in the least deprived Local Authorities and lowest in the most deprived, consistent with the fact that areas with low deprivation have the biggest population of children under 5 on average. Mean expenditure declines steadily over time across all tertiles, while spending on other 0-5 services is much smaller but shows a modest upward trend, particularly in the middle and most deprived groups. Once spending is expressed per child (right panel), the pattern on mandated services spending reverses: the most deprived tertile consistently spends more per 1,000 children on both mandated and non-mandated services, while the least deprived spends the least. Per-child spending on mandated services is also comparatively stable over time in high deprivation LAs, while it declines in the middle and least deprived areas. This pattern is consistent with the workforce trends presented below, where we show that the number of Health Visitors in the most deprived areas has declined more slowly than elsewhere.

**Figure 14:**
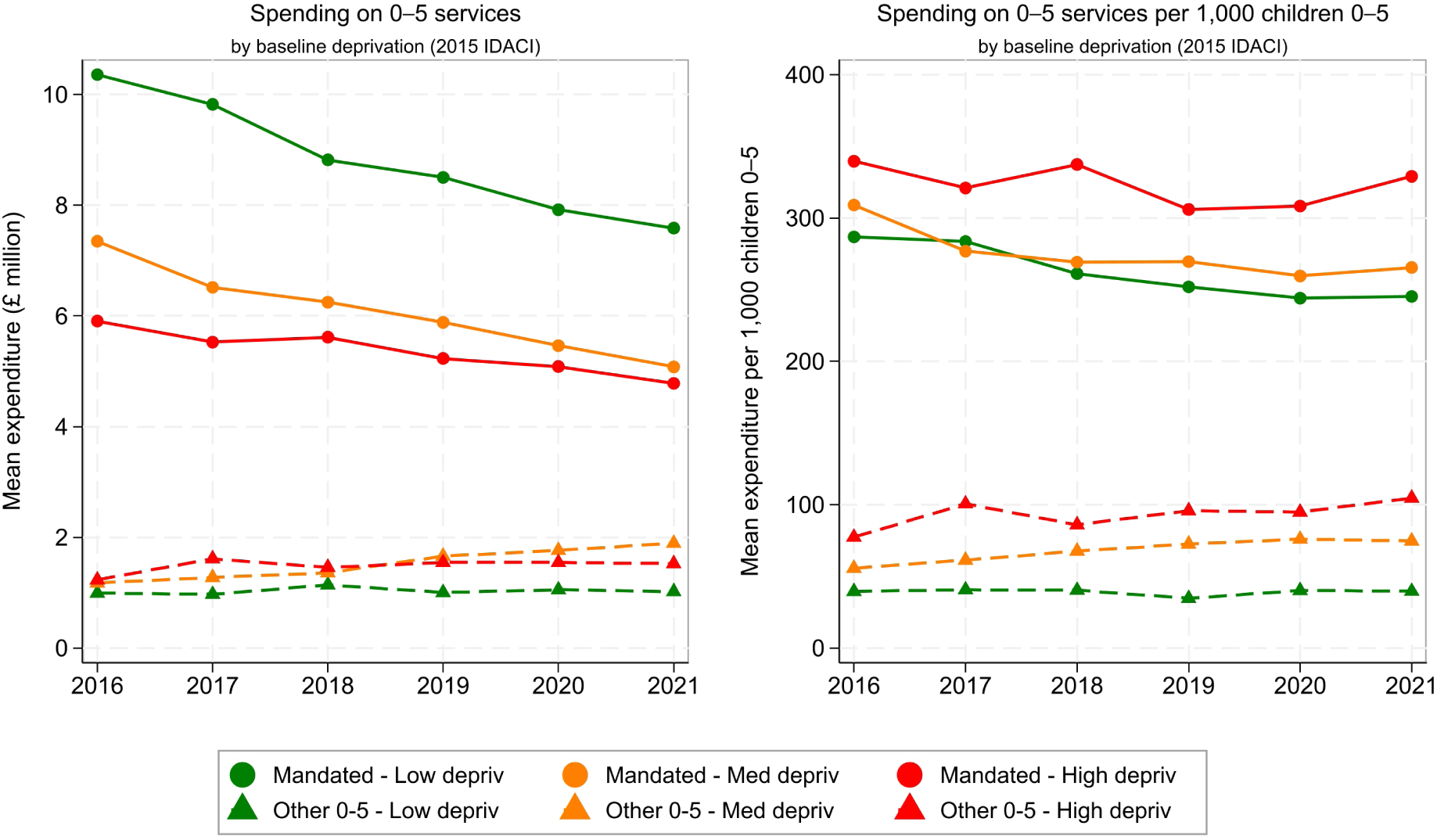
Spending on 0–5 services by baseline deprivation, 2016–2021. Note: Mean Local Authority expenditure on mandated health visiting (solid lines with circles) and other 0–5 services (dashed lines with triangles), shown in total spending (£million; left panel) and per 1,000 children aged 0–5 (right panel), in 2023–24 prices. Lines are stratified by baseline deprivation tertile defined using the 2015 IDACI (Low, Middle, High deprivation). The full sample includes 74 Local Authorities. The number of observations varies across outcomes and years due to missing data.

Tables B5-B7 confirm that the strongest deprivation gradients are in per-child spending. Mandated health visiting spending per child differs significantly across tertiles from 2018 onwards, with pairwise comparisons indicating higher per-child spending in the most deprived relative to the least deprived Local Authorities. A similar pattern holds for non-mandated spending per child, with significant differences from 2017 onwards. By contrast, results for spending in levels are weaker: mandated spending in levels is often significant under ANOVA but not Kruskal-Wallis, and non-mandated spending in levels does not differ significantly across tertiles in any year.

Table B8 shows little evidence that these spending gradients evolved differentially over time. Neither total nor per-child spending on mandated or non-mandated 0-5 services displays systematically different trends across deprivation groups under either specification. The main pattern is therefore persistent cross-sectional differences – especially in per-child terms – combined with broadly similar trajectories over time.

### Health visiting workforce: slower HV decline but weaker CSMS growth in deprived areas

Figure 15 shows similar contrasts for workforce numbers. In levels (left panel), the least deprived Local Authorities employ more Health Visitors, while the most deprived employ fewer, with all groups experiencing a steady decline in HV numbers over time. By contrast, Clinical Skill Mix Staff increase across all tertiles, with the strongest growth in the least and middle deprived groups. Expressed per child (right panel), the pattern reverses for Health Visitors: more deprived areas have more HVs per 1,000 children than less deprived areas, despite declines across all groups between 2016 and 2020 and a partial rebound by 2021. CSMS per child rise over time in all tertiles, with particularly marked increases in the middle and most deprived areas.

**Figure 15:**
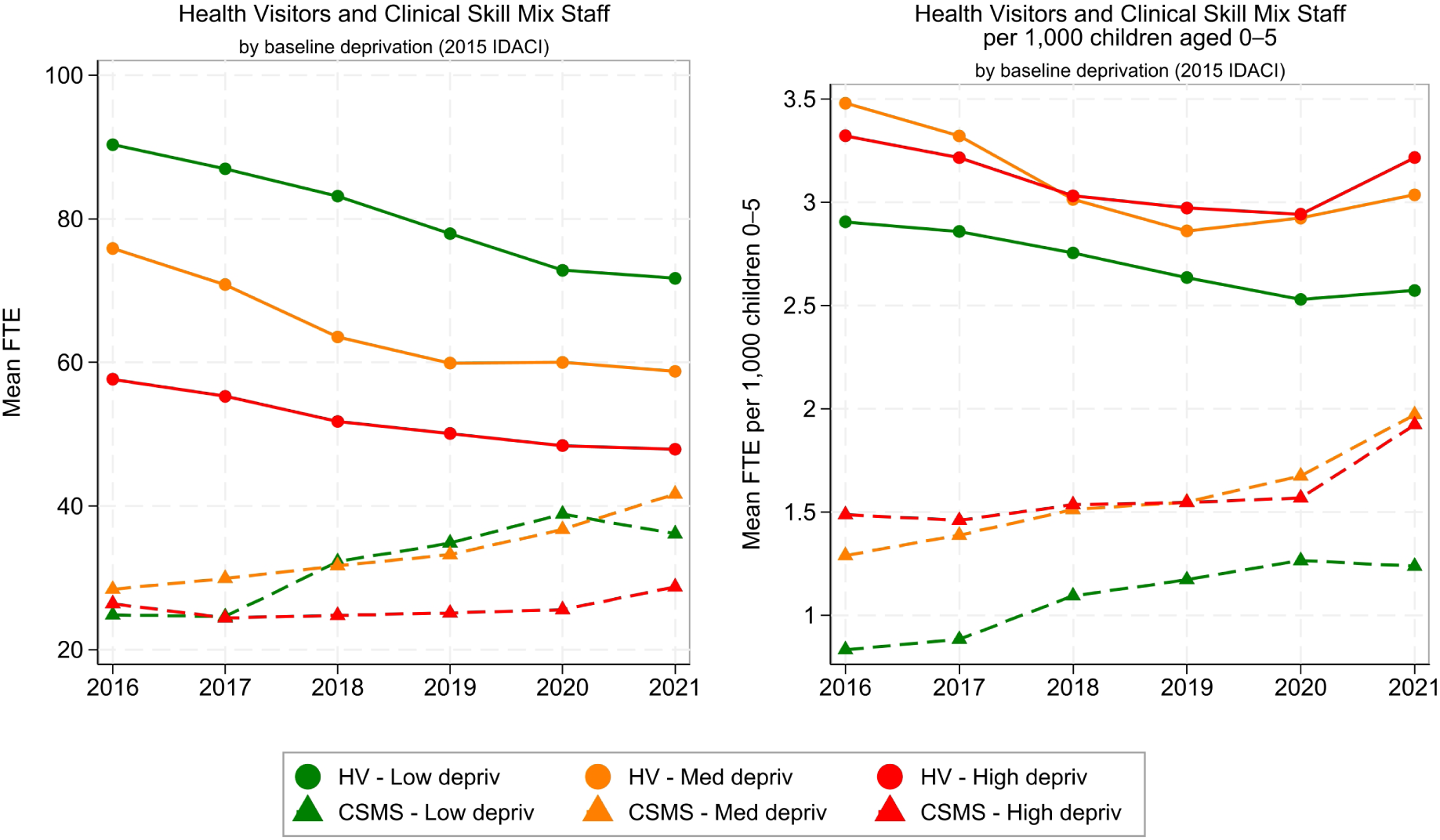
Health visiting workforce by baseline deprivation, 2016–2021. Note: Mean Local Authority Full-Time Equivalent (FTE) workforce levels for Health Visitors (solid lines with circles) and Clinical Skill Mix Staff (dashed lines with triangles), shown in total FTE (left panel) and per 1,000 children aged 0–5 (right panel). Lines are stratified by baseline deprivation tertile defined using the 2015 IDACI (Low, Middle, High deprivation). The full sample includes 74 Local Authorities. The number of observations varies across outcomes and years due to missing data.

Cross-sectional tests reinforce these patterns (Tables B5-B7). Health Visitor numbers differ across tertiles in most year (with higher HV totals in least deprived LAs), while HVs per 1,000 children show little evidence of systematic differences except in 2021. Clinical Skill Mix Staff display the opposite pattern: total CSMS FTE does not differ significantly across tertiles, but CSMS per 1,000 children shows clear differences in 2016-2017 and again in 2021, with higher CSMS intensity in more deprived areas.

Table B8 provides clearer evidence of differential trends for workforce than for spending. HV FTE declines more slowly in more deprived Local Authorities in the linear-trend specification, while HVs per 1,000 children show no evidence of differential trajectories. For CSMS, the year-by-year specification suggests limited divergence in CSMS per-child patterns, but the linear-trend model indicates significantly slower growth in CSMS FTE in more deprived Local Authorities.

### Delivery of mandated health visiting contacts: relative stability across the deprivation gradient

Figure 16 plots completion of mandated health visiting contacts by baseline deprivation. Completion of New Birth Visits is high and stable throughout. For the 6-8 week and 12-month reviews, completion is initially lower in more deprived Local Authorities, with some convergence up to 2020. For the 2-2.5 year review, the most deprived tertile starts from substantially lower completion in 2016 but more than catches up by 2020. Completion of the 2-2.5 year review with ASQ-3 rises steadily across all tertiles until 2020 before falling sharply in 2021, with the largest drop in the most deprived group.

**Figure 16:**
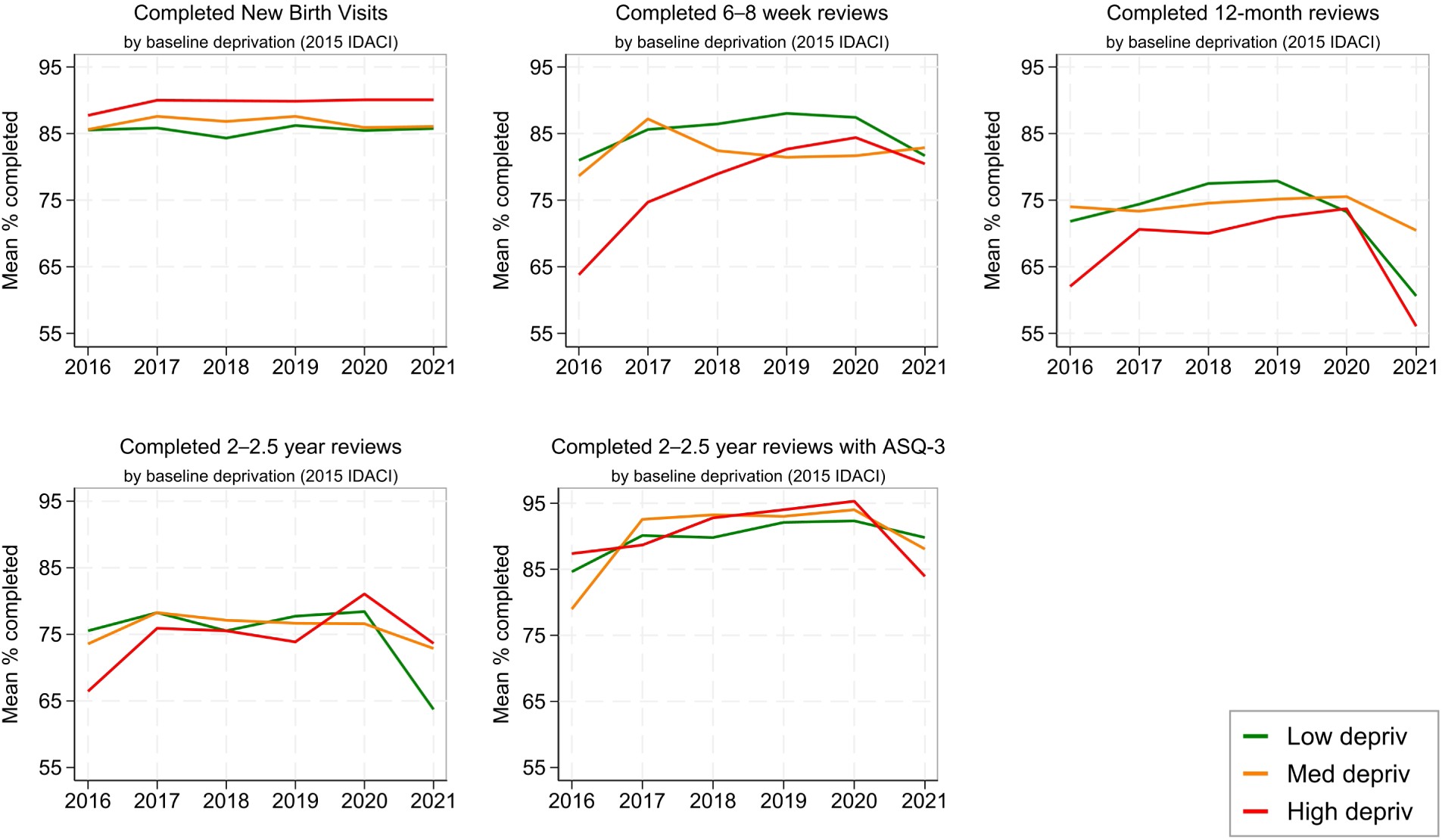
Completion of mandated health visiting contacts by baseline deprivation, 2016–2021. Note: Mean completion rates (%) for mandated health visiting contacts – New Birth Visit, 6-8 week review, 12-month review, 2–2.5 year review, and 2–2.5 year review with ASQ-3 – by baseline deprivation tertile defined using the 2015 IDACI (Low, Middle, High deprivation). Each panel plots annual mean completion rates for 2016–2021. The full sample includes 74 Local Authorities. The number of observations varies across outcomes and years due to missing data.

Once again, these patterns are confirmed by formal cross-sectional tests (Tables B5-B7). New Birth Visits show no meaningful differences in any year. For the 6-8 week review, disparities appear in earlier years but disappear by 2021. Evidence of differences in the 12-month review is weak and confined to 2021, and we find no persistent differences for the 2-2.5 year review or ASQ-3 completion. Overall, delivery outcomes are less consistently stratified by deprivation than spending or workforce measures.

Fixed effects regressions nonetheless indicate differential trends for a subset of contacts (Table B8). In the year-by-year specification, trajectories differ by deprivation for the 6-8 week and 12-month reviews. In the linear-trend specification, the only consistent pattern is faster improvement over time in more deprived areas for the 6-8 week review, whereas trends in other mandated contacts do not differ systematically across groups.

### Summary: linking resources and service delivery

Spending on 0-5 services declined across Local Authorities at all deprivation levels during a period of sustained reductions in the public health grant. While more deprived Local Authorities experienced sharper funding cuts,^31^ the evidence above shows that the distribution of observed resources remains broadly progressive: more deprived areas spend more per child and employ more HVs and CSMS per child, even though they spend less in total and employ smaller workforces in levels.^32^ At the same time, spending trajectories have evolved similarly across deprivation tertiles over time, indicating that cuts have been system-wide, while workforce evolved more unevenly: declines in Health Visitor FTE have been slower in more deprived areas, but growth in Clinical Skill Mix Staff has also been weaker.

Despite these differences in inputs, completion of mandated health visiting contacts shows relatively limited variation by deprivation. The main exception is the 6-8 week review, where more deprived areas show faster improvement over time. However, these delivery patterns coincide with rising caseload pressure, which increased most sharply in the most deprived Local Authorities in the pre-pandemic years (see Figure B5). Relatedly, this data does not speak to the quality of contacts. For example, Liu et al. (2024) document substantial variation in both the duration of contacts and the share of face-to-face visits across Local Authorities. As a result, the similarities in completion of contacts may mask important differences in the intensity or content of support received by families, particularly in higher-need settings.

## 7. Conclusions

Health Visitors are the foundation of the Healthy Child Programme, leading the delivery of universal and targeted support for families with children under five. Yet since the devolution of commissioning to Local Authorities in 2015, the service has faced declining numbers of qualified Health Visitors, growing reliance on lower-banded Clinical Skill Mix Staff, rising caseloads, and sustained cuts to statutory funding. Using new FOI data, this report has provided a comprehensive picture of the health visiting workforce between 2016 and 2021, presenting evidence on spending trends in mandated and non-mandated services and linking workforce pressures to service delivery.

Our findings reveal that the number of qualified Health Visitors fell by more than a fifth between 2016 and 2021, while the number of CSMS rose by a third. Despite this partial substitution, overall workforce capacity contracted, and in nearly three-quarters of Local Authorities, average caseloads exceeded the recommended threshold of 250 children per practitioner. These challenges were magnified during the COVID-19 pandemic, when widespread redeployment of staff further reduced capacity, even as safeguarding risks and family vulnerabilities increased. At the same time, Local Authority spending on mandated services declined by almost 20% in real terms between 2016-17 and 2024-25.

The patterns in our inequality analysis add important perspective. Resources remain broadly progressive, with more deprived Local Authorities typically spending more on health visiting per child and employing more health visiting staff than less deprived areas. However, the downward trajectory of spending is similar across LAs of all deprivation levels, making it evident that the cuts are system-wide.

The policy implications are clear. An under-resourced workforce risks undermining the Government’s renewed emphasis on prevention and early intervention, as set out in the NHS 10-Year Health Plan (Department of Health and Social Care, 2025) and the Department for Education’s new early years strategy (Department for Education, 2025). Both agendas place health visiting at the centre of ambitions to shift resources from hospitals to communities and to improve integration and quality in the early years system. Yet without urgent action to restore workforce capacity, these ambitions will remain structurally constrained. Our analysis shows that the scale of investment required to reduce caseloads to safe levels is substantial but feasible in light of recent early years funding commitments, such as the expansion of Best Start Family Hubs. Rebuilding the health visiting workforce is crucial to enable wider initiatives in prevention and community-based support to achieve their intended impact.

Looking forward, two priorities stand out. First, to secure long-term funding that allows Local Authorities to rebuild the health visiting workforce and restore recommended caseload levels, with a particular focus on addressing regional disparities. Second, to establish a unified workforce data system that captures all providers and staff groups, enabling consistent monitoring and planning. These steps are essential to ensure that health visiting can fulfil its statutory role, reduce inequalities in child health and development, and act as the foundation on which the Government’s broader prevention and early years strategies can succeed.

Based on our findings, we set out the following recommendations:

- Establish a unified national data system to capture the full health visiting workforce across all providers, including CSMS, with standardised reporting on caseloads and service delivery.
- Secure sustainable funding for Local Authorities to restore safe caseload levels, with priority given to reducing regional inequalities and explicitly supporting and strengthening provision in the most deprived areas.
- Align health visiting workforce restoration with the rollout of Best Start Family Hubs, Healthy Babies, and the wider NHS prevention agenda, ensuring that new investments can deliver maximum impact.

## Data Availability

The analyses in this manuscript use a dataset compiled by the authors from Freedom of Information responses provided by Local Authorities and service providers in England, together with publicly available secondary data cited in the manuscript. The compiled FOI-derived analytic dataset is available from the authors upon reasonable request. Publicly available secondary data can be accessed from the original sources referenced in the manuscript.

## Appendix A. A brief history of health visiting in England

### The early years of health visiting

Health visiting in England originated in the early 1860s, with services designed to support and inform new mothers about practices that fostered a healthy home environment (Institute of Health Visiting, n.d.). Since then, the profession has evolved substantially and is now among England’s most important preventative community health services.

By 1905, around 50 towns had appointed paid health visitors, whose work focused on the prevention of illnesses and the promotion of public health. Professionalisation followed in the early twentieth century: the Royal Sanitary Institute began overseeing qualifying courses in 1916, and in 1919 the Ministry of Health introduced the first statutory qualification for health visiting (Institute of Health Visiting, 2012).

### Health visiting as a statutory service in the 20^th^ century

Health visiting became a universal statutory service under the Local Government Act 1929. As described in Institute of Health Visiting (2012), from 1925 the Ministry of Health assumed responsibility for training, initially requiring a midwifery qualification for entry, and after 1945, nursing registration became mandatory to practise. From 1929 until 1974, through and beyond the creation of the National Health Service (NHS) in 1948, the profession remained based within local government, before responsibility was transferred to the NHS. The four principles of health visiting, introduced in 1977, continue to underpin practice today: to search for health needs, stimulate awareness of health needs, influence policies affecting health, and facilitate health-enhancing activities (Institute of Health Visiting, 2012).

### The Healthy Child Programme (2009-present)

Health visiting staff are responsible for delivering the 0-5 element of the Healthy Child Programme (HCP), introduced in 2009 as England’s main early intervention and prevention public health framework. The programme aims to improve family health, reduce inequalities, and support children’s development (Department of Health, 2009).

#### The National Health Visiting Programme (2011-2015)

Between 2011 and 2015, the government implemented the National Health Visiting Programme (NHVP), which funded the recruitment of an additional 4,200 full-time equivalent (FTE) health visitors (Department of Health, 2011). This was the largest workforce expansion since the post-war period, designed to strengthen the early years public health offer and rebuild universal provision.

#### The 2015 devolution and public health grant cuts

In 2015, commissioning responsibility for 0-5 children public health services was devolved from the NHS to Upper Tier Local Authorities (LAs) (Department of Health, 2018), allowing them to outsource health visiting services, not only to NHS Trusts but also to independent providers of community health services (such as social enterprises) or local councils. This reform marked a major structural shift in early years service delivery.

The transition coincided with reductions to the ring-fenced public health grant from the central government, which funds the delivery of public health services. The public health grant totalled £2.66 billion in 2013/14 and rose to £3.46 billion in 2015/16 following the devolution. However, between 2016/17 and 2019/20, it was reduced by 2.2-2.7% annually. (Gulland, 2017; Institute of Health Visiting, 2019, 2022, 2025).

#### The 4-5-6 approach (2016-2021)

From 2016, the HCP adopted the ‘4-5-6 approach’, structured around four levels of service, five mandated health reviews, and six high-impact areas, all led by Health Visitors (Institute of Health Visiting, 2016). This model remained in place when commissioning guides were updated in 2018 (UK Government, 2018). The four levels of service comprised: Community, linking families with local resources such as Children’s Centres and community-led services; Universal, ensuring all families received five key health reviews; Universal Plus, providing rapid expert help for families with specific needs, such as support with parental mental health or domestic violence; and Universal Partnership Plus, offering ongoing support for families with complex needs.

The five mandated reviews take place at 28 weeks of pregnancy, 10-14 days after birth, 6-8 weeks, 9-12 months, and 2-2.5 years. Through these contacts, Health Visitors and other team members build relationships with families to identify needs, monitor children’s health and development, and provide advice on breastfeeding, nutrition, and healthy behaviours. In addition, HVs lead six early years high impact areas: (i) parenthood and early weeks, (ii) maternal mental health, (iii) breastfeeding, (iv) healthy weight, (v) minor illness and accidents, and (vi) healthy 2 year olds and getting ready for school.

#### The current model: Universal in Reach - Personalised in Response (from 2021)

In 2021, the HCP framework was revised and rebranded as the ‘Universal in Reach - Personalised in Response’ model (UK Government, 2021). This replaced the previous 4-5-6 approach while retaining a structure based on four levels of service depending on the individual and family needs: community, universal, targeted, and specialist levels of support (UK Government, 2021).

The updated model reinforced the principle that all families should receive a universal offer, while additional and specialist input should be personalised and proportionate to risk and individual circumstances. Two additional suggested (non-mandated) contacts were introduced at 3 and 6 months, and the terminology of the high-impact areas was updated to place greater emphasis on mental health and early learning (UK Government, 2021).

## Appendix B. Additional information on data collection, sample composition and analysis

### FOI template and definitions used to classify and designate staff under either Health Visitors or Clinical Skill Mix Staff

During our data collection, we specified in the FOI requests the definitions used to classify staff as Health Visitors of Clinical Skill Mix Staff. We followed the definitions from notes 28 and 29 from the nursing, midwifery, and health visiting staff matrix of the Occupation Code Manual (NHS Digital, n.d.a). The general definition for a Health Visitor is: “an employee who holds a qualification as a Registered Health Visitor under the Specialist Community Public Health Nursing part of the NMC Register and who occupies a post where such a qualification is a requirement”. Notes 28 and 29 in the Manual specify the following:

Code as Health Visitors:

- Qualified nurses/midwives who also hold a qualification as a Registered Health Visitor under the Specialist Community Public Health Nursing part of the NMC Register working directly with children and families;
- Qualified and registered Health Visitors who perform specific activities such as providing breastfeeding advice to parents;
- Family nurses working within the Family Nurse Partnership Programme who are qualified and registered as Health Visitors;
- Sure Start Children’s Centre qualified and registered named Health Visitors;
- Managers within a Health visiting team who hold a health visiting qualification and registration and are involved in clinical work or safeguarding.

Code as Clinical Skill Mix Staff (Not Health Visitors):

- Any person working in a health visiting team who does not hold a qualification and registration as a Health Visitor;
- Any person who holds a qualification and registration as a Health Visitor but is not employed in a role where this is a requirement;
- Managers within a health visiting team who hold health visiting qualification and registration but are not involved in clinical work or safeguarding.

Figure B1 shows the most recent iteration of the FOI template we used during data collection.

**Figure B1:**
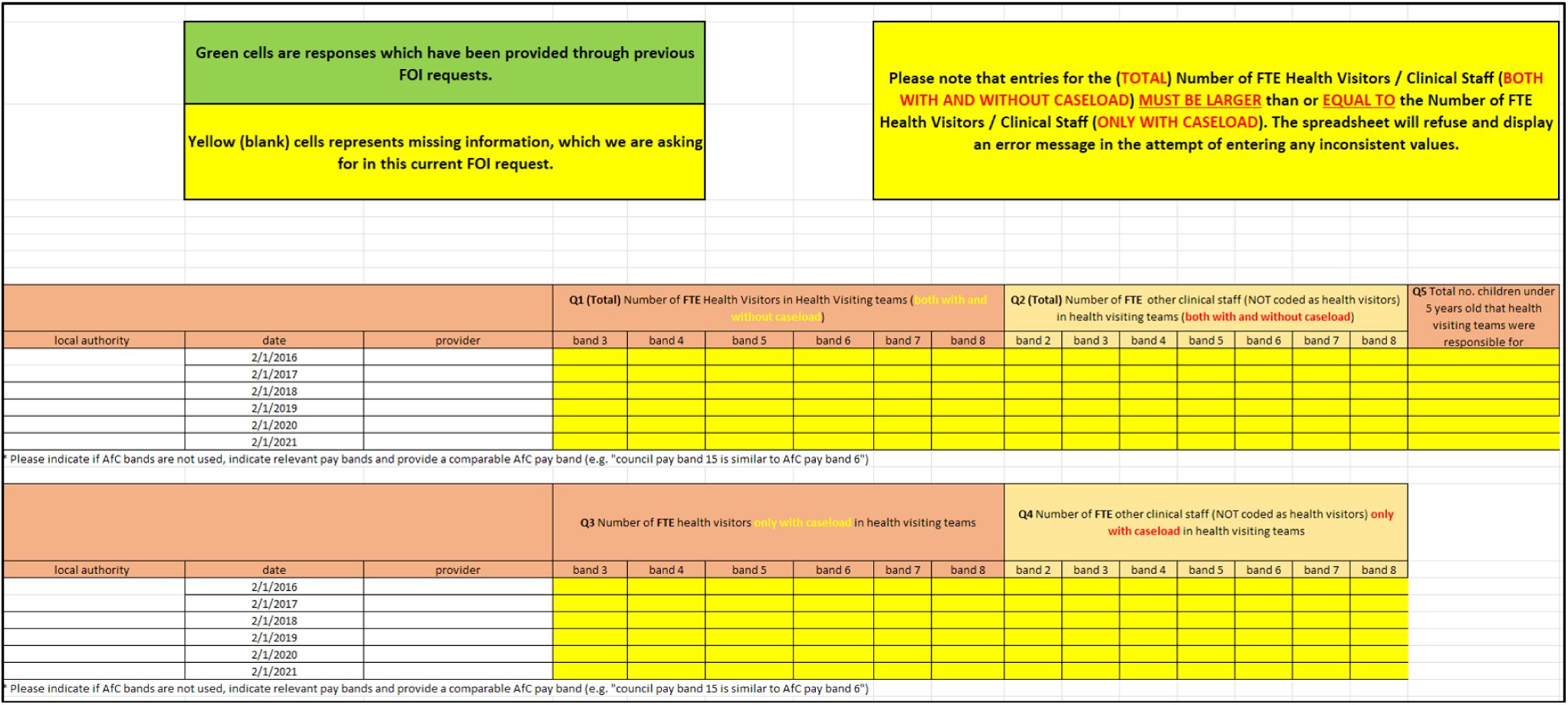
FOI template used during the latest round of data collection. Note: Data from Q5 has not been reported or used throughout the report. This template was the latest iteration drafted after a series of FOI data collections, refined over time to improve accuracy while maintaining comparability.

Figure B2 provides a visual representation of the decision-making process during our FOI data collection, using a flowchart.

**Figure B2:**
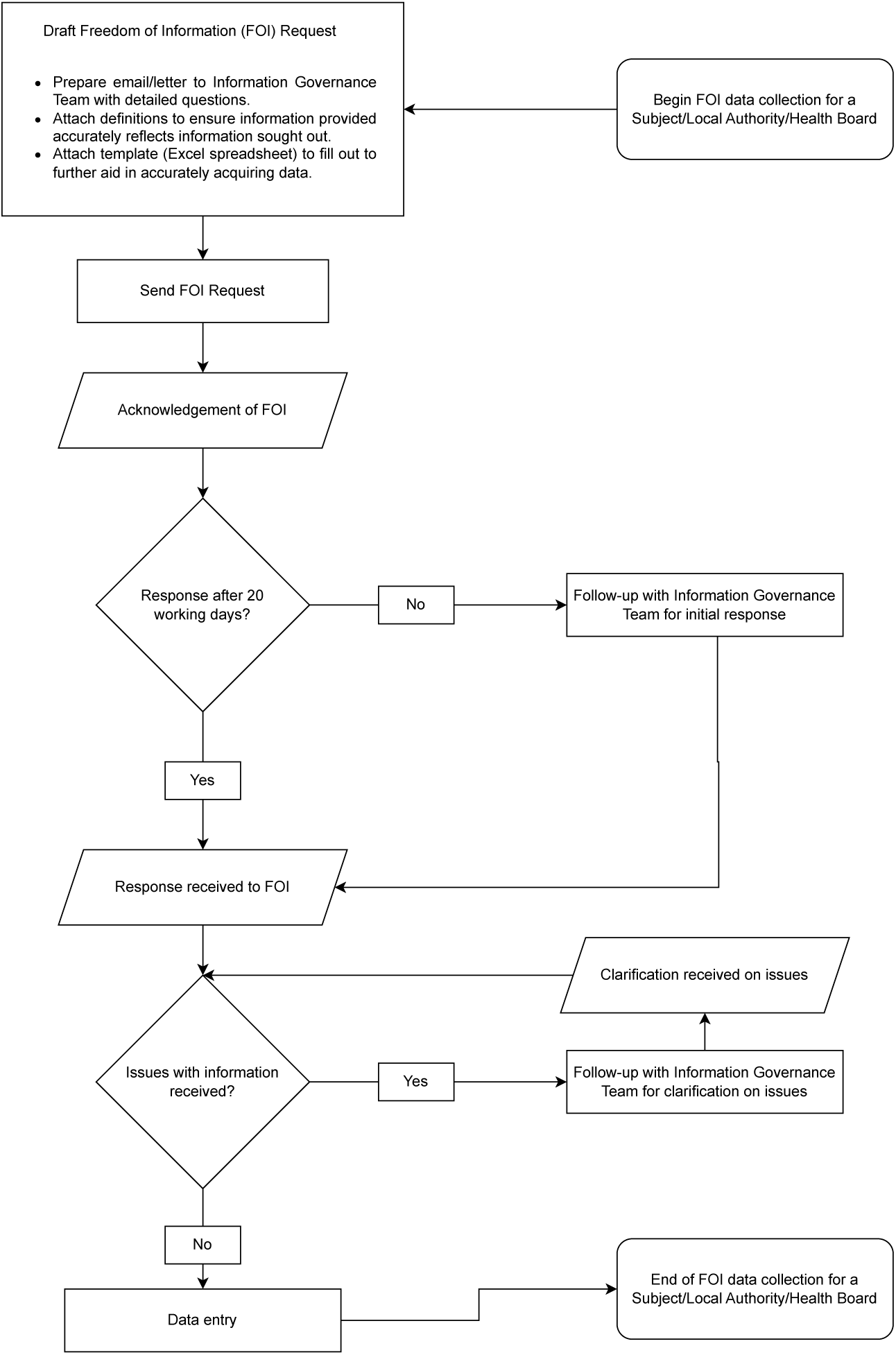
FOI data collection process map.

### Additional information on composition and characteristics of the samples

**Table B1:** Comparison of mean LA characteristics by sample.

|  | Complete total FTE sample |  |  | Complete caseload FTE sample |  |  |
| --- | --- | --- | --- | --- | --- | --- |
|  | Mean<br>excluded LAs | Mean<br>included LAs | Mean diff. | Mean<br>excluded LAs | Mean<br>included LAs | Mean diff. |
| IMD 2015 - Crime | 0.053 | 0.159 | -0.107 | 0.061 | 0.163 | -0.102 |
| IMD 2015 - Education, Skills and Training | 22.026 | 22.067 | -0.041 | 21.965 | 22.149 | -0.185 |
| IMD 2015 - Employment | 0.125 | 0.127 | -0.002 | 0.125 | 0.127 | -0.002 |
| IMD 2015 - Health Deprivation and Disability | 0.043 | 0.14 | -0.097 | 0.054 | 0.14 | -0.086 |
| IMD 2015 - Income Deprivation Affecting Children Index | 0.209 | 0.216 | -0.007 | 0.21 | 0.215 | -0.005 |
| IMD 2015 - Income | 0.155 | 0.159 | -0.004 | 0.156 | 0.159 | -0.003 |
| IMD 2019 - Crime | 0.071 | 0.126 | -0.055 | 0.063 | 0.144 | -0.081 |
| IMD 2019 - Education, Skills and Training | 22.221 | 21.91 | 0.312 | 22.109 | 22.005 | 0.104 |
| IMD 2019 - Employment | 0.104 | 0.106 | -0.002 | 0.104 | 0.106 | -0.002 |
| IMD 2019 - Health Deprivation and Disability | 0.054 | 0.089 | -0.034 | 0.054 | 0.094 | -0.039 |
| IMD 2019 - Income Deprivation Affecting Children Index | 0.179 | 0.182 | -0.003 | 0.18 | 0.181 | -0.001 |
| IMD 2019 - Income | 0.137 | 0.139 | -0.002 | 0.137 | 0.139 | -0.002 |
| Total Population (in thousands) | 341.56 | 409.01 | -67.45 | 351.32 | 406.24 | -54.92 |
| Population under 5 (in thousands) | 21.33 | 25.24 | -3.91 | 21.94 | 25.03 | -3.09 |
| % of children achieving a good level of development | 68.45 | 65.9 | 2.55 | 66.74 | 67.68 | -0.95 |
| % of children achieving at least expected level<br>in all prime areas of learning | 76.99 | 74.44 | 2.55 | 75.1 | 76.43 | -1.33 |
| % of children obese (4-5 years old) | 9.23 | 9.56 | -0.33 | 9.14 | 9.71 | -0.58* |
| % of children overweight (4-5 years old) | 21.69 | 22.44 | -0.74 | 21.56 | 22.71 | -1.15* |
| % of children underweight (4-5 years old) | 1.02 | 0.99 | 0.03 | 1.01 | 1 | 0 |
| CLA Rate (per 10,000 children 1 year old) | 64 | 60.37 | 3.62 | 63.25 | 60.75 | 2.51 |
| CLA Rate (per 10,000 children 1-4 years old) | 37.77 | 36.82 | 0.96 | 36.59 | 38.13 | -1.54 |
| CIN Total Episodes rate (per 10,000 children under 18) | 673.14 | 704.71 | -31.57 | 670.05 | 712.94 | -42.89 |
| CPP Total rate (per 10,000 children under 18) | 45.96 | 47.94 | -1.98 | 45.99 | 48.18 | -2.19 |
| Nominal Public Health Ring Fenced Grant Allocation<br>(per children under 5) | 1055.82 | 1138.88 | -83.06 | 1060.78 | 1144.20 | -83.43 |
| Nominal Net Current Expenditures on<br>prescribed 0-5 children's services (per children under 5) | 119.19 | 116.96 | 2.23 | 116.94 | 119.50 | -2.56 |
| Number of mothers who received a first face to face<br>antenatal contact with a Health Visitor at 28 weeks + | 1678.38 | 1762.64 | -84.26 | 1642.01 | 1819.62 | -177.6 |
| % of births that received a face to face New Birth Visit<br>within 14 days by a Health Visitor | 86.23 | 86.3 | -0.08 | 85.41 | 87.29 | -1.88 |
| % of infants who received a 6-8 week review by the time<br>they were 8 weeks | 81.95 | 74.8 | 7.14* | 80.81 | 75.17 | 5.63 |
| % of children who received a 12 month review by the time<br>they turned 12 months | 74.4 | 69.4 | 5 | 73.39 | 69.94 | 3.45 |
| % of children who received a 2-2.5 year review | 72.2 | 71.92 | 0.28 | 71.87 | 72.29 | -0.42 |
| % of children who received a 2-2.5 year review using ASQ-3 | 83.19 | 83.26 | -0.07 | 81.83 | 85.21 | -3.39 |

\*\*\* p<0.01, \*\* p<0.05, \*p<0.1

**Table B2:** Distributional comparison of LA characteristics by sample.

|  | Complete total FTE |  | Complete caseload FTE |  |
| --- | --- | --- | --- | --- |
|  | D statistic | p-value | D statistic | p-value |
| IMD 2015 - Crime | 0.195 | 0.125 | 0.196 | 0.127 |
| IMD 2015 - Education, Skills and Training | 0.078 | 0.979 | 0.117 | 0.711 |
| IMD 2015 - Employment | 0.115 | 0.721 | 0.112 | 0.752 |
| IMD 2015 - Health Deprivation and Disability | 0.115 | 0.718 | 0.089 | 0.937 |
| IMD 2015 - Income Deprivation Affecting Children Index | 0.140 | 0.468 | 0.122 | 0.655 |
| IMD 2015 - Income | 0.154 | 0.350 | 0.124 | 0.640 |
| IMD 2019 - Crime | 0.136 | 0.504 | 0.115 | 0.722 |
| IMD 2019 - Education, Skills and Training | 0.082 | 0.967 | 0.068 | 0.996 |
| IMD 2019 - Employment | 0.116 | 0.705 | 0.080 | 0.973 |
| IMD 2019 - Health Deprivation and Disability | 0.105 | 0.813 | 0.102 | 0.843 |
| IMD 2019 - Income Deprivation Affecting Children Index | 0.128 | 0.581 | 0.100 | 0.861 |
| IMD 2019 - Income | 0.142 | 0.452 | 0.112 | 0.749 |
| Total Population (in thousands) | 0.149 | 0.389 | 0.155 | 0.343 |
| Population under 5 (in thousands) | 0.160 | 0.304 | 0.182 | 0.180 |
| % of children achieving a good level of development | 0.157 | 0.333 | 0.105 | 0.818 |
| % of children achieving at least expected level<br>in all prime areas of learning | 0.145 | 0.428 | 0.118 | 0.699 |
| % of children obese (4-5 years old) | 0.172 | 0.229 | 0.158 | 0.329 |
| % of children overweight (4-5 years old) | 0.154 | 0.350 | 0.150 | 0.393 |
| % of children underweight (4-5 years old) | 0.082 | 0.978 | 0.084 | 0.973 |
| CLA Rate (per 10,000 children 1 year old) | 0.134 | 0.567 | 0.119 | 0.721 |
| CLA Rate (per 10,000 children 1-4 years old) | 0.101 | 0.856 | 0.071 | 0.993 |
| CIN Total Episodes rate (per 10,000 children under 18) | 0.110 | 0.766 | 0.124 | 0.638 |
| CPP Total rate (per 10,000 children under 18) | 0.104 | 0.825 | 0.101 | 0.855 |
| Nominal Public Health Ring Fenced Grant Allocation<br>(per children under 5) | 0.129 | 0.577 | 0.120 | 0.672 |
| Nominal Net Current Expenditures on<br>prescribed 0-5 children's services (per children under 5) | 0.072 | 0.992 | 0.098 | 0.885 |
| Number of mothers who received a first face to face<br>antenatal contact with a Health Visitor at 28 weeks + | 0.145 | 0.428 | 0.144 | 0.442 |
| % of births that received a face to face New Birth Visit<br>within 14 days by a Health Visitor | 0.098 | 0.892 | 0.109 | 0.814 |
| % of infants who received a 6-8 week review by the time<br>they were 8 weeks | 0.199 | 0.123 | 0.201 | 0.118 |
| % of children who received a 12 month review by the time<br>they turned 12 months | 0.172 | 0.243 | 0.179 | 0.206 |
| % of children who received a 2-2.5 year review | 0.110 | 0.789 | 0.123 | 0.674 |
| % of children who received a 2-2.5 year review using ASQ-3 | 0.152 | 0.668 | 0.103 | 0.971 |

\*\*\* p<0.01, \*\* p<0.05, \*p<0.1

**Table B3:** Health visiting workforce composition, 2016-2021.

| Variable | 2016 | 2017 | 2018 | 2019 | 2020 | 2021 |
| --- | --- | --- | --- | --- | --- | --- |
| **Panel A: Total FTE with or without caseload (N=75)** |  |  |  |  |  |  |
| Total no. of FTE HVs | 5631.2 | 5365.7 | 4991.2 | 4714.9 | 4535.7 | 4471.6 |
| Total no. of FTE CSMS | 2002.1 | 1987.5 | 2224.9 | 2337.1 | 2536.7 | 2664 |
| No. of FTE band 5 HVs | 27.2 | 23.4 | 27 | 23.9 | 1.2 | 2.3 |
| No. of FTE band 6 HVs | 4522.1 | 4336.3 | 4115.5 | 3886.9 | 3741.6 | 3663.7 |
| No. of FTE band 7 HVs | 999.1 | 915.6 | 774.8 | 723.6 | 717.9 | 726.2 |
| No. of FTE band 8 HVs | 82.8 | 90.4 | 73.9 | 80.5 | 75 | 79.4 |
| No. of FTE band 2 CSMS | 150.9 | 142.2 | 146.5 | 125.6 | 134.4 | 150.1 |
| No. of FTE band 3 CSMS | 180.9 | 185 | 213.7 | 189.3 | 178.6 | 245.5 |
| No. of FTE band 4 CSMS | 1092.7 | 1167 | 1230.1 | 1268.1 | 1369.7 | 1335.8 |
| No. of FTE band 5 CSMS | 364.7 | 289.2 | 365.2 | 438.7 | 568.7 | 635.4 |
| No. of FTE band 6 CSMS | 71.2 | 50.1 | 113.9 | 150.9 | 143.6 | 162.2 |
| No. of FTE band 7 CSMS | 122.5 | 126.9 | 123.3 | 130.7 | 118 | 109.8 |
| No. of FTE band 8 CSMS | 19.2 | 27.1 | 32.2 | 33.8 | 23.7 | 25.2 |
| **Panel B: Total FTE with caseload (N=66)** |  |  |  |  |  |  |
| Total no. of FTE HVs with caseload | 4840.5 | 4586 | 4314 | 4030.9 | 3764.1 | 3738.3 |
| Total no. of FTE CSMS with caseload | 875.9 | 824.1 | 937.9 | 1021.7 | 967.8 | 1141.2 |
| No. of FTE band 5 HVs with caseload | 19 | 15.2 | 18.8 | 22.7 | 0 | 0 |
| No. of FTE band 6 HVs with caseload | 4091.9 | 3932.1 | 3770.2 | 3537 | 3404.8 | 3336.9 |
| No. of FTE band 7 HVs with caseload | 714.5 | 627.6 | 515.8 | 459.4 | 348.7 | 386.3 |
| No. of FTE band 8 HVs with caseload | 15.1 | 11.1 | 9.2 | 11.8 | 10.6 | 15.1 |
| No. of FTE band 2 CSMS with caseload | 39.2 | 35.9 | 38.8 | 39 | 18.9 | 3.3 |
| No. of FTE band 3 CSMS with caseload | 36.2 | 27.2 | 46.4 | 49 | 45.4 | 57.2 |
| No. of FTE band 4 CSMS with caseload | 495.4 | 509.2 | 504.4 | 492.1 | 451.5 | 532.1 |
| No. of FTE band 5 CSMS with caseload | 168.5 | 140 | 174.1 | 227 | 298.2 | 351.4 |
| No. of FTE band 6 CSMS with caseload | 55 | 34.7 | 90.4 | 131.4 | 117.9 | 130.5 |
| No. of FTE band 7 CSMS with caseload | 72.8 | 69.2 | 76.2 | 74.8 | 34.9 | 58 |
| No. of FTE band 8 CSMS with caseload | 8.8 | 7.9 | 7.6 | 8.4 | 1 | 8.7 |

Note: HVs = Health Visitors. CSMS = Clinical Skill Mix Staff. Band 5 HVs are students. The sample in Panel A includes all 75 Local Authorities in the *complete total FTE* sample, i.e., those with complete data on the number of FTE HVs and CSMS, for all years from 2016 until 2021. The sample in Panel B includes all 66 Local Authorities in the *complete caseload FTE* sample, i.e., those with complete data on the number of total and caseload-holding FTE HVs and CSMS, for all years from 2016 until 2021.

### Additional information on the analysis of delivery metrics and spending

**Figure B3:**
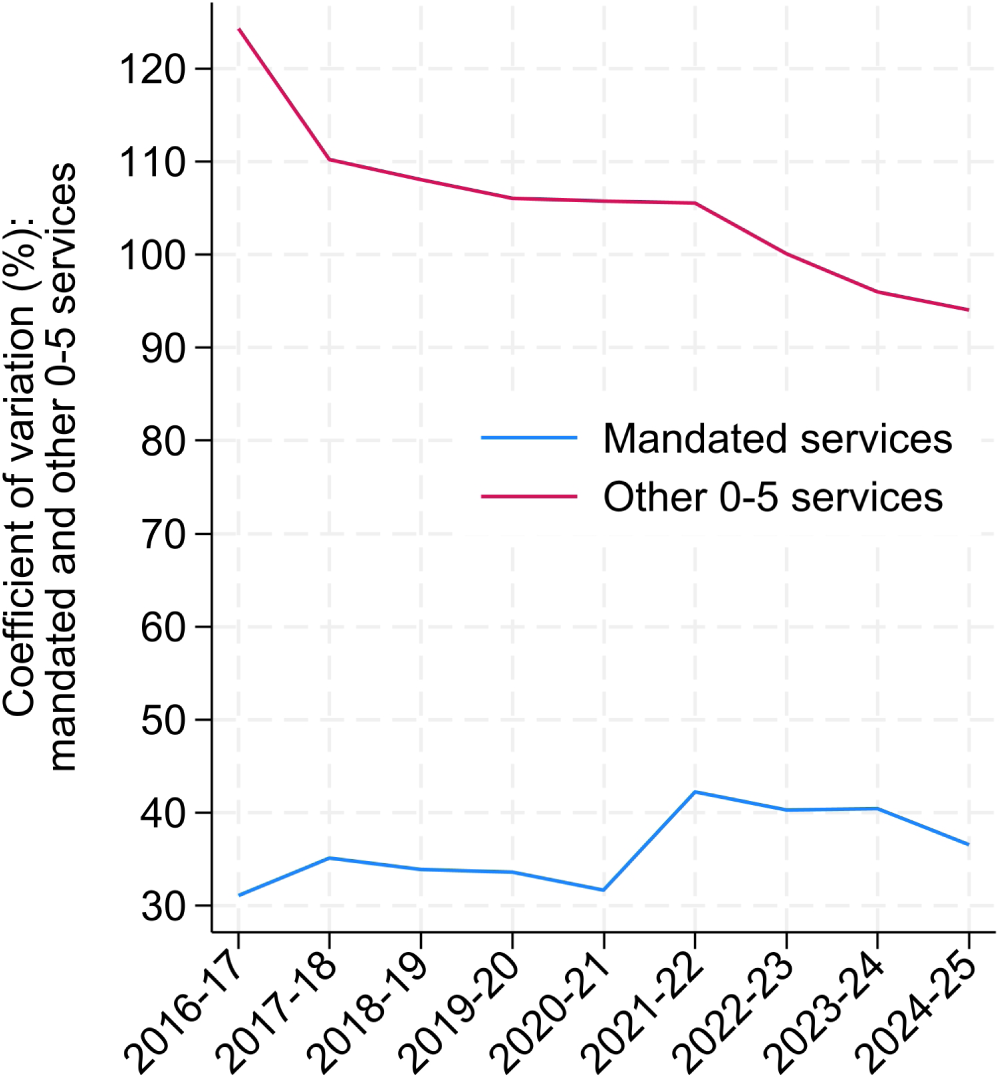
Variation in spending per child on mandated and other 0-5 services, 2016-17 to 2024-25. Note: Coefficient of variation (CV) in Local Authority spending per child under five (in 2023-24 GBP) on mandated 0-5 children’s services (prescribed functions; Line 84) and non-mandated 0-5 children’s services (non-prescribed functions; Line 85). The CV is calculated as the ratio of the standard deviation to the mean across Local Authorities in each financial year, expressed as a percentage. Figures are based on Local Authority revenue outturn (RO3) total expenditures and ONS mid-year population estimates, including all LAs with non-missing RO3 data (N = 146 in 2016-17, 147 in 2017-18 and 2018-19, 149 in 2019-20, 150 in 2020-21, 152 in 2021-22 and 2022-23, 148 in 2023-24, and 147 in 2024-25). Values adjusted using the GDP deflator.

**Table B4:** Local Authority characteristics and health visiting workforce change patterns, 2016– 2021.

|  | (1)<br>Expansion | (2)<br>Substitution | (3)<br>Contraction |
| --- | --- | --- | --- |
| IMD 2015 - Health | 0.322<br>(0.255) | 0.018<br>(0.277) | -0.340<br>(0.236) |
| IMD 2015 - IDACI | -3.374<br>(2.811) | -1.243<br>(3.171) | 4.617*<br>(2.759) |
| % population under 5 | 0.060<br>(0.065) | -0.076<br>(0.097) | 0.016<br>(0.091) |
| % children w/ good level of development | 0.007<br>(0.029) | 0.023<br>(0.037) | -0.031<br>(0.030) |
| % children w/ expected level of learning | -0.004<br>(0.026) | -0.024<br>(0.033) | 0.028<br>(0.027) |
| % of children obese (4-5 y.o.) | 0.036<br>(0.040) | 0.027<br>(0.046) | -0.062*<br>(0.037) |
| CLA rate (per 10k children) | -0.008<br>(0.005) | 0.005<br>(0.005) | 0.002<br>(0.005) |
| CIN rate (per 10k children) | -0.000<br>(0.000) | 0.000<br>(0.000) | -0.000<br>(0.000) |
| CPP rate (per 10k children) | 0.003<br>(0.003) | -0.005<br>(0.005) | 0.002<br>(0.005) |
| % New Birth Visit completeness | -0.001<br>(0.005) | -0.007<br>(0.007) | 0.008<br>(0.006) |
| % 6-8 week review completeness | 0.000<br>(0.002) | 0.002<br>(0.004) | -0.002<br>(0.004) |
| % 12 month review completeness | 0.002<br>(0.003) | 0.000<br>(0.004) | -0.002<br>(0.004) |
| % 2-2.5 year review completeness | -0.003<br>(0.003) | 0.001<br>(0.006) | 0.003<br>(0.005) |
| Public Health Grant allocations per child | 0.000<br>(0.000) | 0.000<br>(0.000) | -0.000<br>(0.000) |
| Spending per child on mandated HV | -0.001<br>(0.001) | -0.002<br>(0.001) | 0.002*<br>(0.001) |
| NHS Trust provider | 1.154***<br>(0.331) | -0.559<br>(0.355) | -0.596**<br>(0.261) |
| Obs. | 73 | 73 | 73 |

### Differences in spending, workforce and service delivery across deprivation tertiles

**Table B5:** Kruskal-Wallis tests for differences across baseline deprivation tertiles.

|  | 2016 | 2017 | 2018 | 2019 | 2020 | 2021 |
| --- | --- | --- | --- | --- | --- | --- |
| Mandated HV spending (£million) | 0.218 | 0.077 | 0.191 | 0.147 | 0.245 | 0.192 |
| Mandated HV spending per child 0-5 | 0.051 | 0.172 | 0.013 | 0.081 | 0.012 | 0.013 |
| Other 0-5 spending (£million) | 0.546 | 0.564 | 0.554 | 0.183 | 0.175 | 0.175 |
| Other 0-5 spending per child 0-5 | 0.112 | 0.095 | 0.048 | 0.007 | 0.006 | 0.003 |
| Public health grant allocation (£million) | 0.582 | 0.582 | 0.582 | 0.582 | 0.509 | 0.606 |
| Public health grant allocation per person | 0.000 | 0.000 | 0.000 | 0.000 | 0.000 | 0.000 |
| Total Health Visitors (FTE) | 0.090 | 0.060 | 0.058 | 0.096 | 0.085 | 0.082 |
| Health Visitors per 1,000 children 0-5 | 0.311 | 0.326 | 0.550 | 0.394 | 0.314 | 0.069 |
| Total Clinical Skill Mix Staff (FTE) | 0.847 | 0.655 | 0.629 | 0.511 | 0.443 | 0.408 |
| Clinical Skill Mix Staff per 1,000 children 0-5 | 0.003 | 0.001 | 0.155 | 0.171 | 0.290 | 0.004 |
| New Birth Visits completed (%) | 0.404 | 0.795 | 0.383 | 0.551 | 0.372 | 0.925 |
| 6-8 week reviews completed (%) | 0.076 | 0.064 | 0.065 | 0.005 | 0.007 | 0.370 |
| 12-month reviews completed (%) | 0.446 | 0.855 | 0.596 | 0.549 | 0.886 | 0.052 |
| 2-2.5 year reviews completed (%) | 0.283 | 0.813 | 0.999 | 0.446 | 0.846 | 0.245 |
| 2-2.5 year reviews with ASQ-3 (%) | 0.306 | 0.450 | 0.743 | 0.436 | 0.639 | 0.349 |

Note: The full sample includes 74 Local Authorities. Each cell reports the p-value from a Kruskal-Wallis test of equality of distributions across three baseline deprivation (IDACI) tertiles, conducted separately for each year. Deprivation tertiles are defined using the 2015 IDACI. The number of observations varies across outcomes and years due to missing data.

**Table B6:**
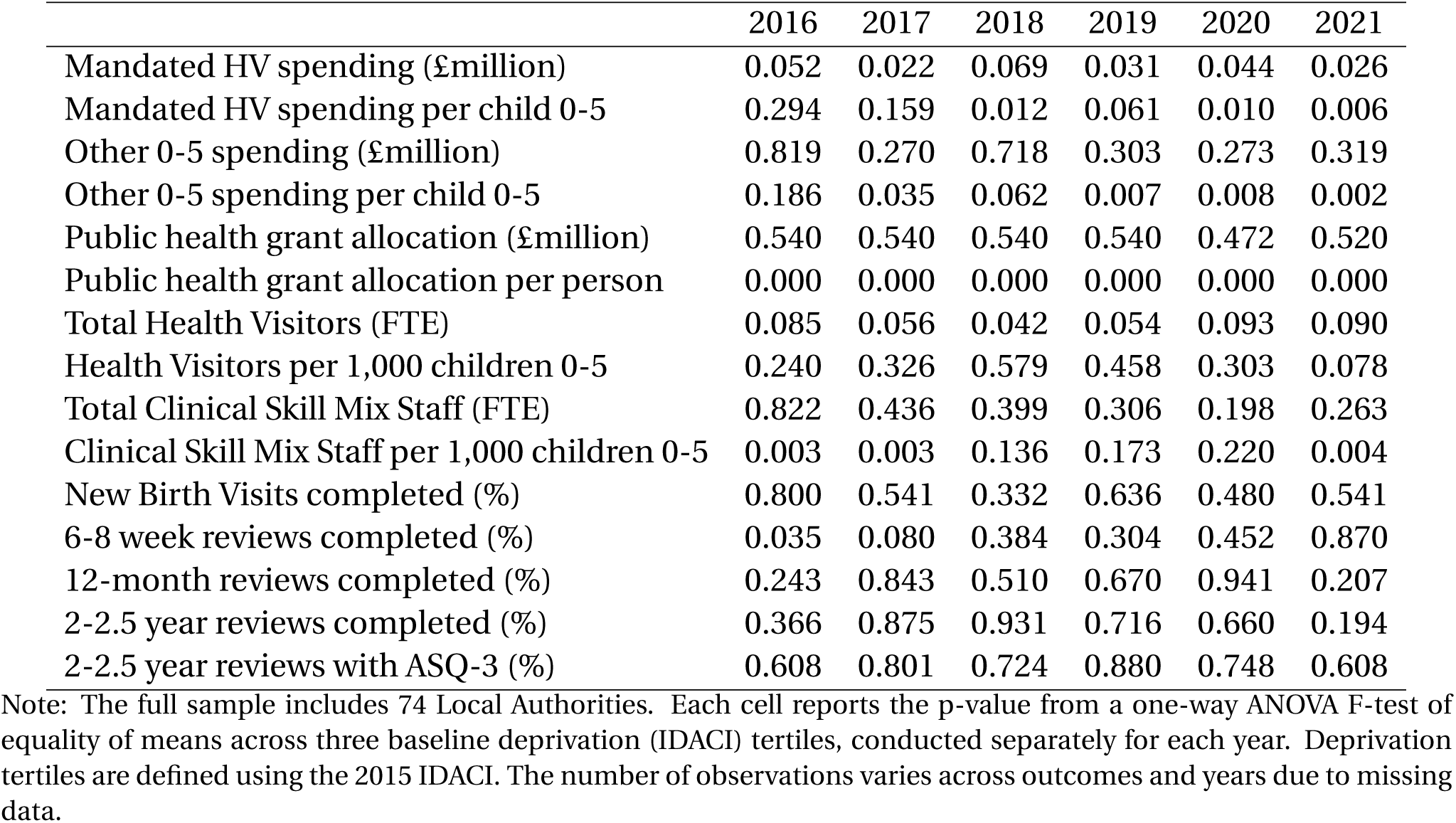
One-way ANOVA tests for differences across baseline deprivation tertiles.

|  | 2016 | 2017 | 2018 | 2019 | 2020 | 2021 |
| --- | --- | --- | --- | --- | --- | --- |
| Mandated HV spending (£million) | 0.052 | 0.022 | 0.069 | 0.031 | 0.044 | 0.026 |
| Mandated HV spending per child 0-5 | 0.294 | 0.159 | 0.012 | 0.061 | 0.010 | 0.006 |
| Other 0-5 spending (£million) | 0.819 | 0.270 | 0.718 | 0.303 | 0.273 | 0.319 |
| Other 0-5 spending per child 0-5 | 0.186 | 0.035 | 0.062 | 0.007 | 0.008 | 0.002 |
| Public health grant allocation (£million) | 0.540 | 0.540 | 0.540 | 0.540 | 0.472 | 0.520 |
| Public health grant allocation per person | 0.000 | 0.000 | 0.000 | 0.000 | 0.000 | 0.000 |
| Total Health Visitors (FTE) | 0.085 | 0.056 | 0.042 | 0.054 | 0.093 | 0.090 |
| Health Visitors per 1,000 children 0-5 | 0.240 | 0.326 | 0.579 | 0.458 | 0.303 | 0.078 |
| Total Clinical Skill Mix Staff (FTE) | 0.822 | 0.436 | 0.399 | 0.306 | 0.198 | 0.263 |
| Clinical Skill Mix Staff per 1,000 children 0-5 | 0.003 | 0.003 | 0.136 | 0.173 | 0.220 | 0.004 |
| New Birth Visits completed (%) | 0.800 | 0.541 | 0.332 | 0.636 | 0.480 | 0.541 |
| 6-8 week reviews completed (%) | 0.035 | 0.080 | 0.384 | 0.304 | 0.452 | 0.870 |
| 12-month reviews completed (%) | 0.243 | 0.843 | 0.510 | 0.670 | 0.941 | 0.207 |
| 2-2.5 year reviews completed (%) | 0.366 | 0.875 | 0.931 | 0.716 | 0.660 | 0.194 |
| 2-2.5 year reviews with ASQ-3 (%) | 0.608 | 0.801 | 0.724 | 0.880 | 0.748 | 0.608 |

Note: The full sample includes 74 Local Authorities. Each cell reports the p-value from a one-way ANOVA F-test of equality of means across three baseline deprivation (IDACI) tertiles, conducted separately for each year. Deprivation tertiles are defined using the 2015 IDACI. The number of observations varies across outcomes and years due to missing data.

**Table B7:** Bonferroni-adjusted pairwise comparisons of means across baseline deprivation tertiles.

| Outcome | 2016 |  |  | 2017 |  |  | 2018 |  |  | 2019 |  |  | 2020 |  |  | 2021 |  |  |
| --- | --- | --- | --- | --- | --- | --- | --- | --- | --- | --- | --- | --- | --- | --- | --- | --- | --- | --- |
|  | L-M | L-H | M-H | L-M | L-H | M-H | L-M | L-H | M-H | L-M | L-H | M-H | L-M | L-H | M-H | L-M | L-H | M-H |
| Mandated HV spending (£million) | 0.304 | 0.053 | 1.000 | 0.117 | 0.026 | 1.000 | 0.227 | 0.088 | 1.000 | 0.129 | 0.039 | 1.000 | 0.132 | 0.066 | 1.000 | 0.080 | 0.044 | 1.000 |
| Mandated HV spending per child 0-5 | 1.000 | 0.362 | 1.000 | 1.000 | 0.399 | 0.222 | 1.000 | 0.020 | 0.041 | 1.000 | 0.062 | 0.334 | 1.000 | 0.011 | 0.077 | 1.000 | 0.006 | 0.052 |
| Other 0-5 spending (£million) | 1.000 | 1.000 | 1.000 | 1.000 | 0.322 | 1.000 | 1.000 | 1.000 | 1.000 | 0.442 | 0.704 | 1.000 | 0.350 | 0.839 | 1.000 | 0.401 | 1.000 | 1.000 |
| Other 0-5 spending per child 0-5 | 1.000 | 0.206 | 0.864 | 1.000 | 0.033 | 0.266 | 0.463 | 0.059 | 1.000 | 0.136 | 0.005 | 0.645 | 0.121 | 0.007 | 0.829 | 0.152 | 0.002 | 0.297 |
| Public health grant allocation (£million) | 0.952 | 1.000 | 1.000 | 0.953 | 1.000 | 1.000 | 0.953 | 1.000 | 1.000 | 0.953 | 1.000 | 1.000 | 0.811 | 0.934 | 1.000 | 0.925 | 1.000 | 1.000 |
| Public health grant allocation per person | 0.026 | 0.000 | 0.000 | 0.028 | 0.000 | 0.000 | 0.027 | 0.000 | 0.000 | 0.026 | 0.000 | 0.000 | 0.032 | 0.000 | 0.000 | 0.067 | 0.000 | 0.000 |
| Total Health Visitors (FTE) | 0.949 | 0.081 | 0.636 | 0.638 | 0.051 | 0.700 | 0.340 | 0.040 | 1.000 | 0.350 | 0.053 | 1.000 | 0.729 | 0.090 | 0.890 | 0.674 | 0.087 | 0.940 |
| Health Visitors per 1,000 children 0-5 | 0.308 | 0.716 | 1.000 | 0.461 | 0.821 | 1.000 | 1.000 | 1.000 | 1.000 | 1.000 | 0.670 | 1.000 | 0.570 | 0.524 | 1.000 | 0.332 | 0.089 | 1.000 |
| Total Clinical Skill Mix Staff (FTE) | 1.000 | 1.000 | 1.000 | 0.820 | 1.000 | 0.779 | 1.000 | 0.671 | 0.778 | 1.000 | 0.453 | 0.691 | 1.000 | 0.280 | 0.475 | 1.000 | 1.000 | 0.312 |
| Clinical Skill Mix Staff per 1,000 children 0-5 | 0.057 | 0.003 | 0.919 | 0.018 | 0.006 | 1.000 | 0.275 | 0.233 | 1.000 | 0.309 | 0.325 | 1.000 | 0.283 | 0.655 | 1.000 | 0.008 | 0.016 | 1.000 |
| New Birth Visits completed (%) | 1.000 | 1.000 | 1.000 | 1.000 | 0.814 | 1.000 | 1.000 | 0.419 | 1.000 | 1.000 | 1.000 | 1.000 | 1.000 | 0.803 | 0.969 | 1.000 | 0.953 | 1.000 |
| 6-8 week reviews completed (%) | 1.000 | 0.049 | 0.110 | 1.000 | 0.200 | 0.113 | 1.000 | 0.506 | 1.000 | 0.444 | 0.726 | 1.000 | 0.628 | 1.000 | 1.000 | 1.000 | 1.000 | 1.000 |
| 12-month reviews completed (%) | 1.000 | 0.604 | 0.339 | 1.000 | 1.000 | 1.000 | 1.000 | 0.750 | 1.000 | 1.000 | 1.000 | 1.000 | 1.000 | 1.000 | 1.000 | 0.666 | 1.000 | 0.257 |
| 2-2.5 year reviews completed (%) | 1.000 | 0.535 | 0.855 | 1.000 | 1.000 | 1.000 | 1.000 | 1.000 | 1.000 | 1.000 | 1.000 | 1.000 | 1.000 | 1.000 | 1.000 | 0.395 | 0.324 | 1.000 |
| 2-2.5 year reviews with ASQ-3 (%) | 1.000 | 1.000 | 1.000 | 1.000 | 1.000 | 1.000 | 1.000 | 1.000 | 1.000 | 1.000 | 1.000 | 1.000 | 1.000 | 1.000 | 1.000 | 1.000 | 1.000 | 1.000 |

Note: The full sample includes 74 Local Authorities. Each cell reports the Bonferroni-adjusted p-value from pairwise comparisons of mean outcomes across baseline deprivation (IDACI) tertiles: Low deprivation vs Middle (L-M), Low vs High (L-H), Middle vs High (M-H), conducted separately for each year. Deprivation tertiles are defined using the 2015 IDACI. The number of observations varies across outcomes and years due to missing data.

**Table B8:**
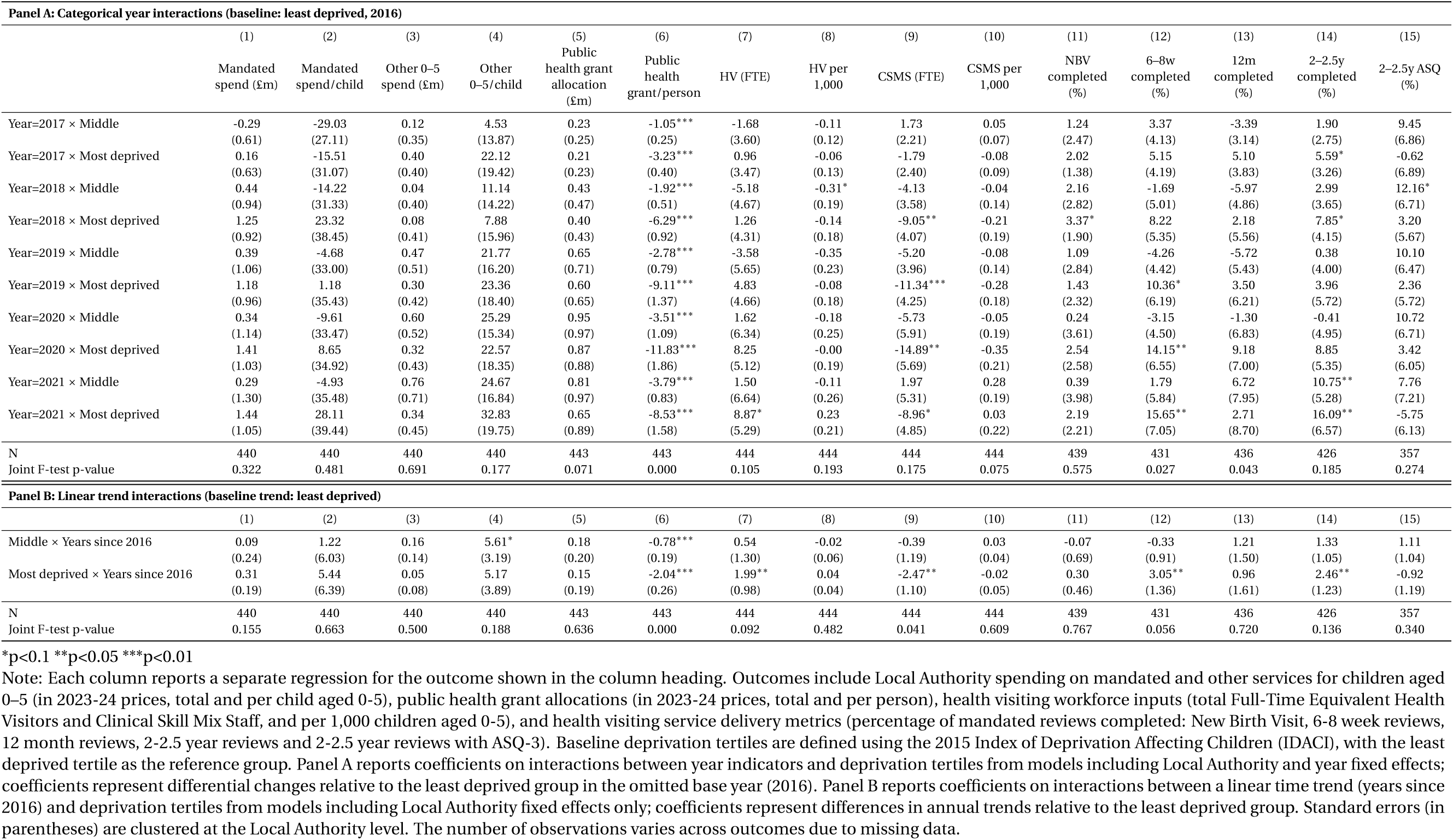
Differential trends over time by baseline deprivation: Local Authority fixed effects regressions.

**Figure B4:**
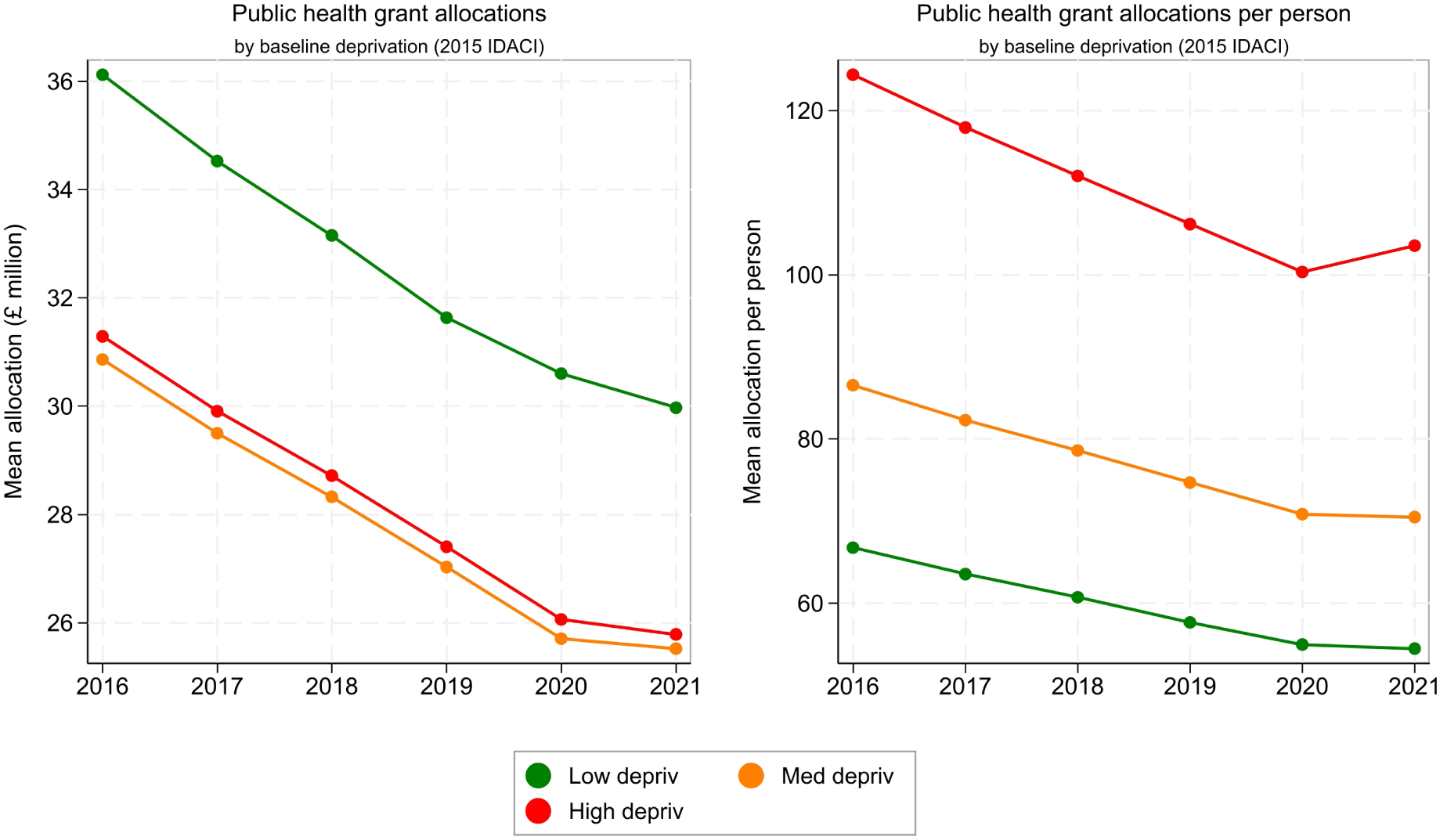
Public health grant allocations by baseline deprivation, 2016–2021. Note: Mean Local Authority public health grant allocations shown in total (£million; left panel) and per person (right panel), in 2023–24 prices. Lines are stratified by baseline deprivation tertile defined using the 2015 IDACI (Low, Middle, High deprivation). The full sample includes 74 Local Authorities. The number of observations varies across outcomes and years due to missing data.

**Figure B5:**
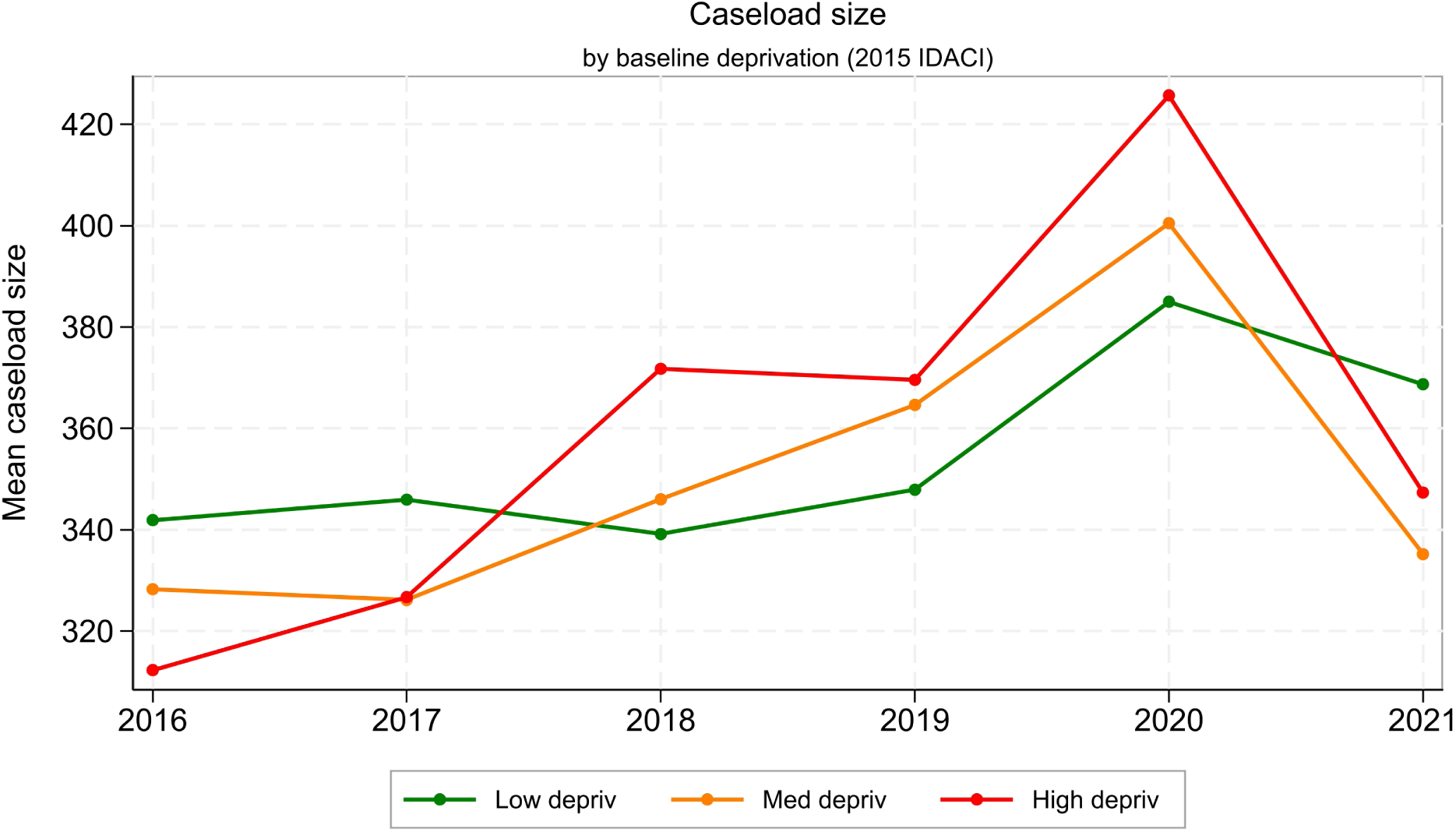
Caseload size by baseline deprivation, 2016–2021. Note: Mean caseload size, defined as the number of children aged 0-5 per Full-Time Equivalent caseload-holding health visiting staff, shown by baseline deprivation tertile defined using the 2015 Income Deprivation Affecting Children Index (IDACI): Low, Middle, and High deprivation. Lines plot annual means for the period 2016–2021 among 65 LAs with complete caseload data.

## Appendix C. The health visiting workforce in Greater London

Figure C1 shows the percentage of Health Visitors within the health visiting workforce across London boroughs from 2016 to 2021. On average, London boroughs started and remained below the national level: 62.8% HV share in 2016 compared with 73.6% in England, and 55.7% in 2021 compared with 63.8% nationally. Despite this lower baseline, London broadly mirrored the overall downward trajectory seen across England.

**Figure C1:**
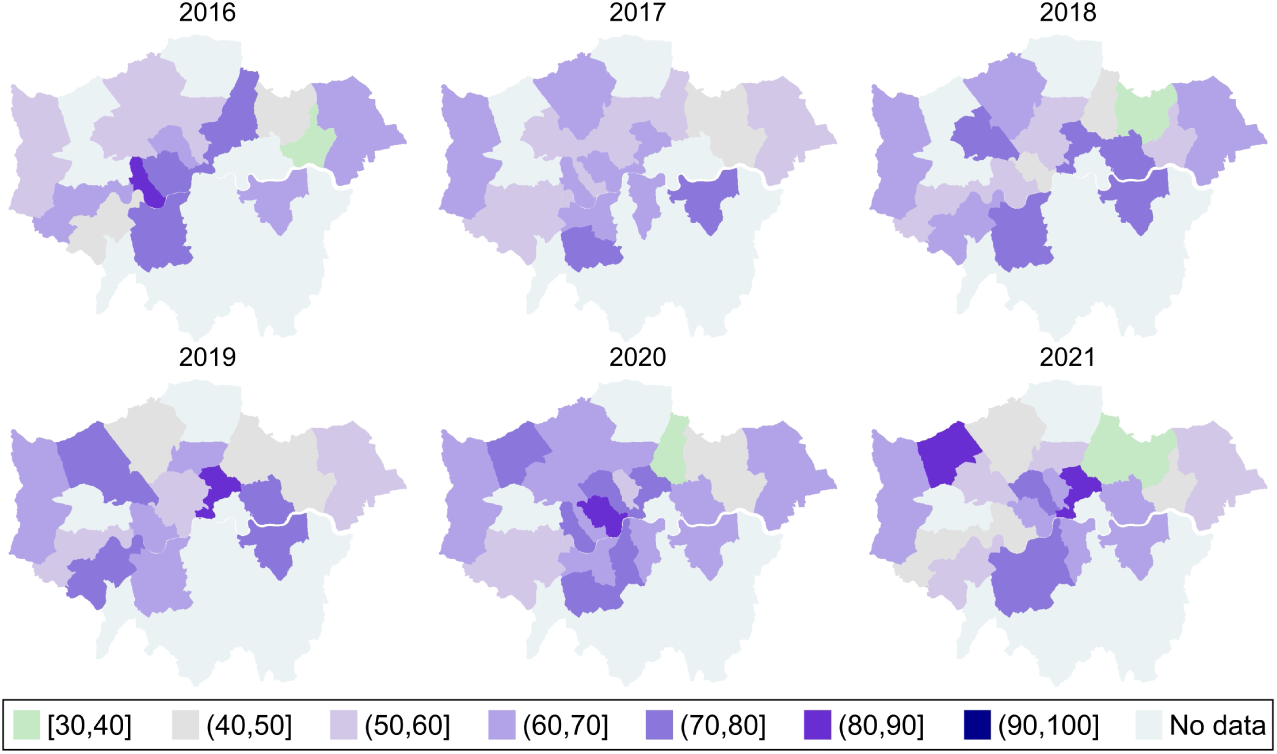
Percentage of Health Visitors in the total workforce, by London borough and year. Note: Percentage of Health Visitors in the total health visiting workforce, by London borough and year. Darker shading indicates a higher HV share. Based on the most complete sample available in each year.

Among the 19 boroughs with complete data, 57.9% recorded a decline in HV share, while 42.1% saw an increase. This also contrasts with the national picture, where nearly four-fifths of Local Authorities reported declines, indicating that although reductions were the dominant trend in London, increases were relatively more common than elsewhere.

To further examine how local workforce composition evolved, Figure C2 classifies London boroughs according to the direction of change in the number of Health Visitors and Clinical Skill Mix Staff between 2016 and 2021.

Of the 19 boroughs, 36.8% saw a reduction in HVs alongside an increase in CSMS, while another 42.1% experienced declines in both groups of staff. Only one borough (5.3%) reported increases in both HVs and CSMS, and one (5.3%) maintained HV levels while expanding CSMS. Two boroughs (10.5%) increased HVs while reducing CSMS. Compared to the England-wide figures (where over half of LAs combined HV reductions with CSMS increases), London shows a more even split between substitution (HV down, CSMS up) and broader contraction (both down).

Turning to caseload-holding staff, Figure C3 presents the percentage change in the number of caseload-holding HVs and CSMS in London boroughs between 2016 and 2021.

Between 2016 and 2021, 82.4% of London boroughs reported a decline in caseload-holding HVs, while only 11.8% saw an increase and 5.9% remained unchanged. By contrast, trends for caseload-holding CSMS were more mixed: 47.1% of boroughs reported increases, 29.4% no change, and 23.5% decreases. This suggests that, similar to the national pattern, many London teams attempted to mitigate HV shortfalls by relying more heavily on CSMS, though the shift was less uniform than in England overall.

**Figure C2:**
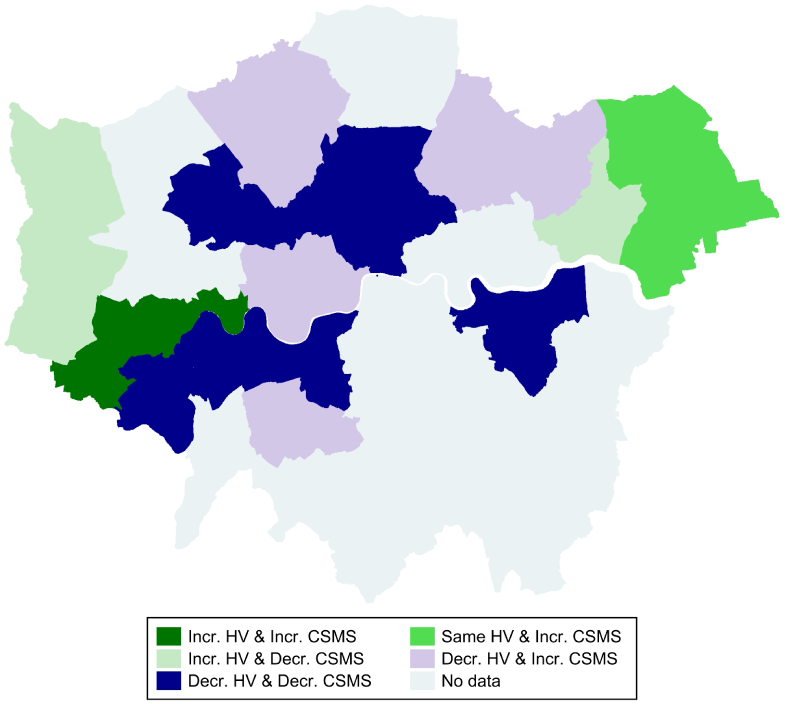
Change in workforce composition, 2016–2021, London boroughs. Note: Classification of London boroughs by the direction of change in Health Visitor and Clinical Skill Mix Staff numbers between 2016 and 2021. Based on 19 boroughs with complete data for both years.

**Figure C3:**
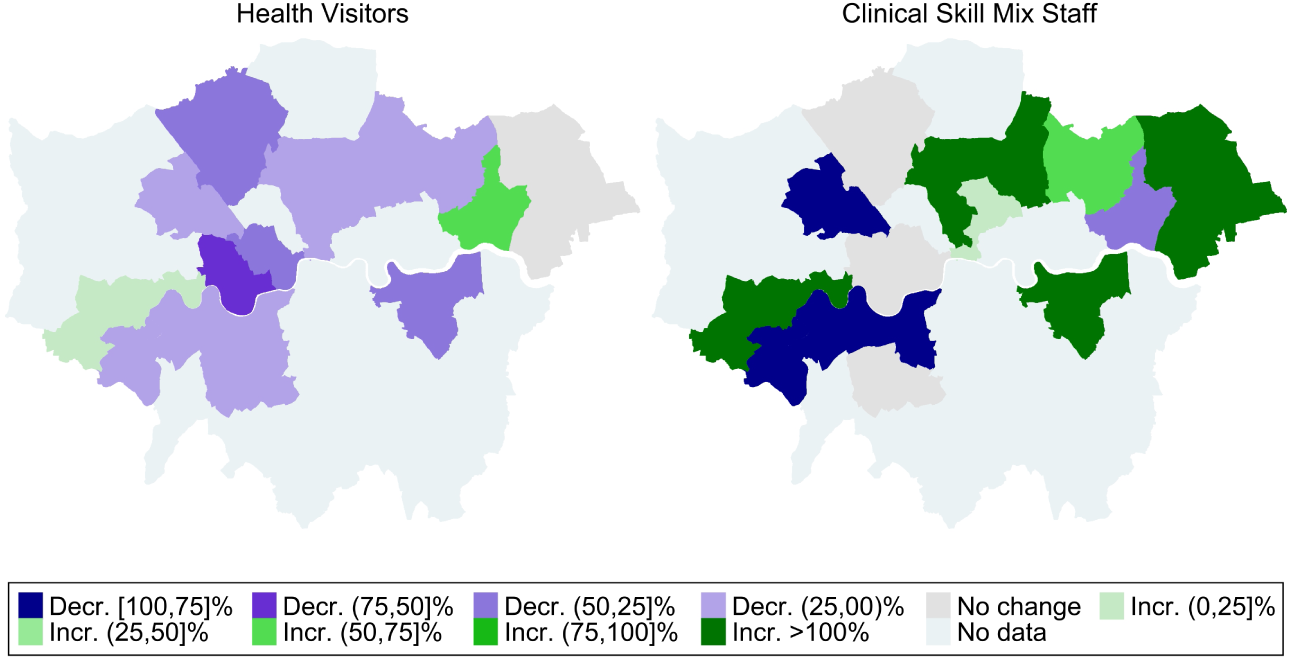
Change in number of caseload-holding HVs and CSMS, 2016–2021, London boroughs. Note: Left panel shows percentage change in the number of caseload-holding Health Visitors; right panel shows percentage change in caseload-holding Clinical Skill Mix Staff. Based on 17 London boroughs with complete caseload data in both 2016 and 2021.

## Appendix D. The health visiting workforce in Wales and Scotland

### Workforce composition before COVID-19

As a benchmark to the English picture described in the main report, we also collected FOI data from all seven Health Boards in Wales and fourteen Health Boards in Scotland, capturing workforce composition on 1 February 2020. Table D1 presents descriptive statistics of Health Visitors and Clinical Skill Mix Staff, broken down by pay band.

**Table D1:** Distribution of health visiting staff in Wales and Scotland on 1 February 2020.

| Variables | Wales |  |  |  | Scotland |  |  |  |
| --- | --- | --- | --- | --- | --- | --- | --- | --- |
|  | Mean | Std. Dev. | Min | Max | Mean | Std. Dev. | Min | Max |
| Total FTE Health Visitors | 128.3 | 60.42 | 28.90 | 217.2 | 120.4 | 130.5 | 6.800 | 471.9 |
| Total FTE Clinical Skill Mix Staff | 38.42 | 25.08 | 8.6 | 80.11 | 26.34 | 25.75 | 0.600 | 74.36 |
| FTE band 6 Health Visitors | 107.3 | 51.65 | 23.90 | 188.4 | 0 | 0 | 0 | 0 |
| FTE band 7 Health Visitors | 20.00 | 10.96 | 4 | 35.80 | 119.4 | 129.0 | 6.800 | 469.9 |
| FTE band 8 Health Visitors | 1 | 1.155 | 0 | 3 | 0.964 | 2.804 | 0 | 10.50 |
| FTE band 2 Clinical Skill Mix Staff | 0.143 | 0.378 | 0 | 1 | 1.368 | 4.344 | 0 | 16.33 |
| FTE band 3 Clinical Skill Mix Staff | 2.215 | 2.885 | 0 | 7.410 | 4.724 | 8.847 | 0 | 30.90 |
| FTE band 4 Clinical Skill Mix Staff | 26.92 | 16.16 | 8.150 | 48.60 | 13.17 | 13.64 | 0 | 38.50 |
| FTE band 5 Clinical Skill Mix Staff | 5.215 | 9.388 | 0 | 23.89 | 5.489 | 10.30 | 0 | 34.37 |
| FTE band 6 Clinical Skill Mix Staff | 1.046 | 1.393 | 0 | 3.050 | 0.297 | 0.866 | 0 | 3.160 |
| FTE band 7 Clinical Skill Mix Staff | 2.286 | 5.443 | 0 | 14.60 | 1.300 | 3.852 | 0 | 14.60 |
| FTE band 8 Clinical Skill Mix Staff | 0.571 | 1.134 | 0 | 3 | 0 | 0 | 0 | 0 |

Note: FTE staff numbers include both caseload holders and staff without caseload. Std. Dev. = standard deviation. Min. = minimum. Max = maximum.

On average, Health Boards in Wales employed 128.3 FTE HVs and 38.4 FTE CSMS, while in Scotland the averages were slightly lower at 120.4 and 26.3, respectively. However, the number of HVs in Scotland showed much greater variation across Health Boards, as reflected in the higher standard deviation. Looking at pay bands, the composition of the workforce also differed markedly. In Wales, HVs were concentrated in band 6, with a smaller share at band 7 and very few at band 8, while most CSMS were employed in bands 4 and 5. In Scotland, by contrast, virtually all HVs were recorded at band 7, with only marginal numbers in band 8. This reflects a policy change introduced in late 2018, which required HV posts to be graded at band 7, thereby phasing out band 6 roles. The distribution of CSMS also diverged, with Scotland employing more staff in bands 2 and 3 than Wales, but fewer in band 4.

Figure D1 displays the share of Health Visitors in the health visiting workforce across Health Boards in Wales and Scotland, as of February 2020.

**Figure D1:**
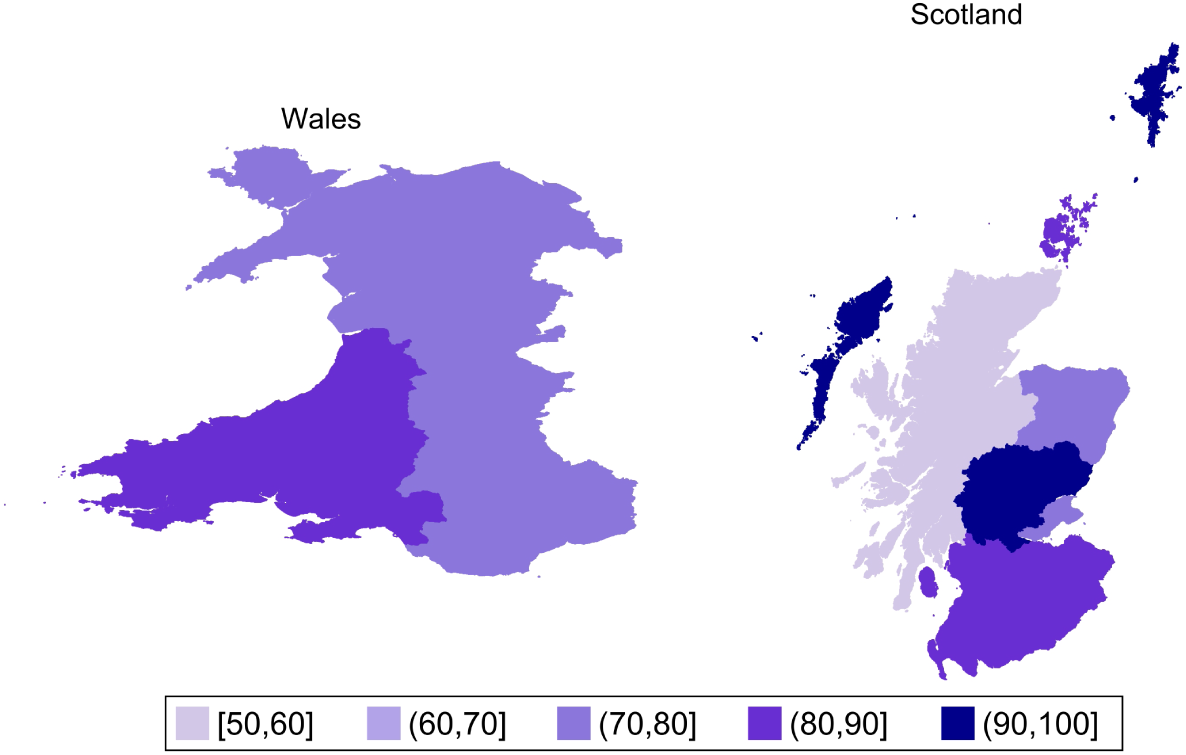
Percentage of Health Visitors in the total workforce across Health Boards in Wales and Scotland (1 February 2020) Note: Percentage of Health Visitors in the total health visiting workforce, by Health Board in Wales and Scotland, as of 1 February 2020. Darker shading indicates a higher HV share.

**Figure D2:**
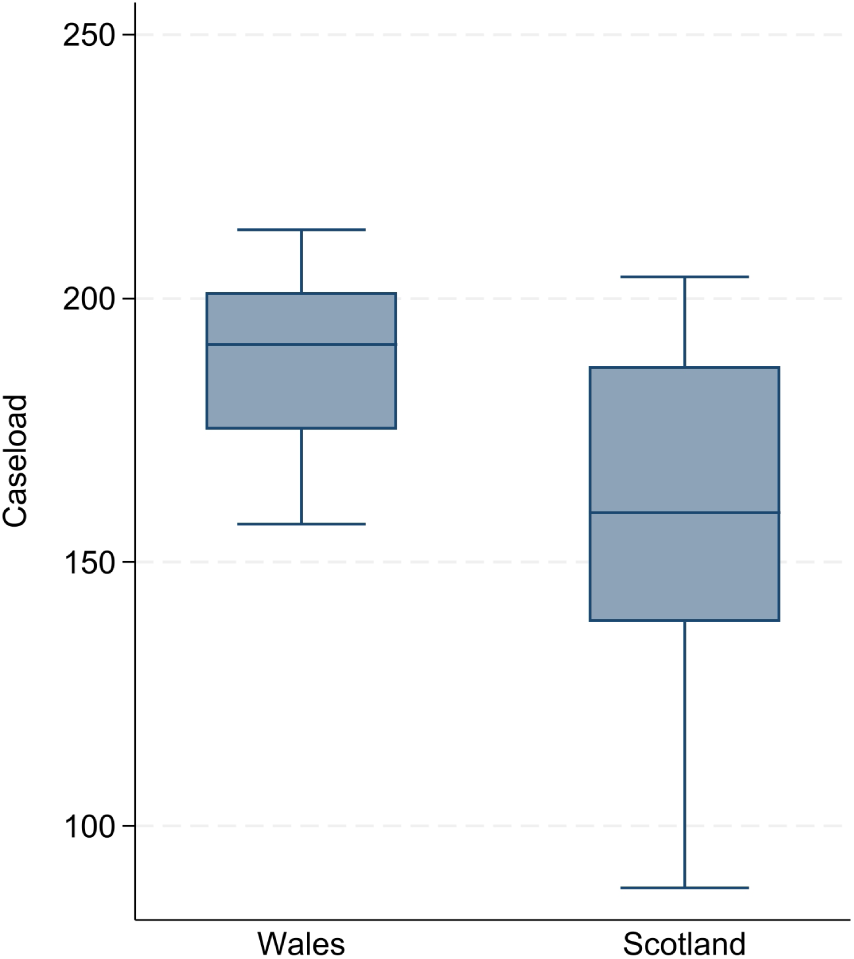
Caseload sizes of health visiting staff across Health Boards in Wales and Scotland (1 February 2020) Note: Caseload is calculated as the number of children under 5 per FTE caseload-holding staff, using population numbers provided in the FOI responses or the mid-year population estimates from the National Records of Scotland when an FOI response was not available. Whiskers represent the upper and lower adjacent values, defined as the 75th/25th percentile ± 1.5 times the interquartile range (the distance between the 25th and 75th percentiles). The sample covers all 7 Welsh Health Boards and 13 Scottish Health Boards (excluding Lothian, which did not report caseload-holding staff).

Two features stand out in stark contrast to England. First, there were no Health Boards in Scotland or Wales where HVs made up less than half of the workforce – i.e., nowhere were CSMS in the majority. Second, the average share of HVs was substantially higher: HVs accounted for 78% of the workforce in Wales and 83.2% in Scotland, compared with only 63.9% in England at the same point in time. Within this overall picture, Wales showed a fairly tight distribution, with all Health Boards between 70.5% and 85.8%. Scotland, by contrast, combined a slightly higher average with greater dispersion: while most Health Boards recorded HV shares above 80%, and some exceeded 90%, there was also one with a much lower share, down to 57.3%.

Figure D2 shows the distribution of caseload sizes across Health Boards in Wales and Scotland on 1 February 2020. In both nations, caseload-holding staff were almost exclusively Health Visitors. In Wales, only Cardiff and Vale reported any caseload-holding CSMS, and in Scotland, only Ayrshire and Arran did so; even in these cases, HVs remained the large majority, accounting for 82.3% and 80% of caseload-holding staff, respectively. Across all other Health Boards, HVs were the sole caseload holders.

Caseload sizes were markedly lower than in England, where the 2020 average reached around 400 children per FTE staff member. In Wales, the mean caseload was 189 children per FTE staff. In Scotland, the mean was 159 children per FTE staff.^33^ None of the Health Boards across the two nations exceeded the Institute of Health Visitings recommended threshold of 250 children per HV.

### Redeployment of health visiting staff in Wales and Scotland during COVID-19

Health Boards in Wales and Scotland were also under extreme pressure to provide urgent services during the early months of the pandemic, with health visiting staff redeployed to other roles. In Scotland, national guidance for health visiting was issued on 7 April 2020 and updated on 17 April 2020 (Scottish Government, 2020). The guidance halted non-essential and routine face-to-face visits, including antenatal contacts, but advised that Health Visitors should still be required to be available and responsive to parents to promote, support and safeguard the well-being of children and young people. Standard in-person contacts at 3-5 weeks, 3 months, 4 months, 8 months, and child health reviews were left to professional judgement, with phone or remote alternatives encouraged, and in-person visits reserved for essential needs. Only the 11-14 day and 6-8 week checks were explicitly recommended to continue face-to-face.

In Wales, the first guidance, published on 23 March 2020, advised Health Boards to prioritise contacts at 10-14 days, 6-8 weeks, and 6 months (Welsh Government, 2020). Updated guidance issued on 13 May 2020 advised reinstating additional contacts and working towards the full schedule. However, the second firebreak lockdown in October-November 2020, followed by the third lockdown beginning in December, led to further restrictions. On 22 December 2020, the Welsh Government advised Health Boards to risk assess cessation or the reduction of health visiting service (Welsh Government, 2020). As in Scotland, in-person contacts were largely replaced with phone, video, or instant messaging, which inevitably affected the quality of universal provision. The annual Healthy Child Wales Programme (HCWP) report, published in August 2021, noted that parental anxiety, shielding, and self-isolation reduced take-up of visits, while many contacts did not occur simply because parents did not answer the phone when they were called.

To document these changes, we collected data through FOI requests. Requests were submitted to all Scottish Health Boards on 16 November 2020, and to all Welsh Health Boards on 19 March 2021. Most Scottish responses were received in December 2020 and January 2021, with final data collected by August 2021. Responses from Welsh Health Boards were received between 13 April and 20 May 2021, and data collection was completed by 13 July 2021.

Building on this, we now examine the scale and timing of redeployment of Health Visitors and Clinical Skill Mix Staff. Figures D3-D6 present redeployment numbers across Health Boards in Wales and Scotland between March 2020 and March 2021, highlighting the pressures on both HVs and CSMS.^34^

Figure D3 displays the percentage of HVs redeployed in Wales. Between 19 March and 31 August 2020, during the first COVID-19 lockdown, almost all Health Boards reported redeploying HVs, with the highest levels in the southeast. Redeployment fell sharply between 1 September and 7 October 2020, when only one Board continued to redeploy staff. During the second “firebreak” lockdown (23 October-9 November 2020), only three Boards reported renewed redeployment, and by the first quarter of 2021, redeployment had largely subsided.

Figure D4 shows HV redeployment in Scotland. Between 19 March and 31 August 2020, half of all Boards redeployed HVs, though percentages were generally lower than in Wales. Redeployment decreased significantly between 1 September and 7 October 2020, when only Ayrshire and Arran reported redeployment. In the final quarter of 2020, two Boards (Ayrshire and Arran, and Lanarkshire) reported continuing redeployment.

**Figure D3:**
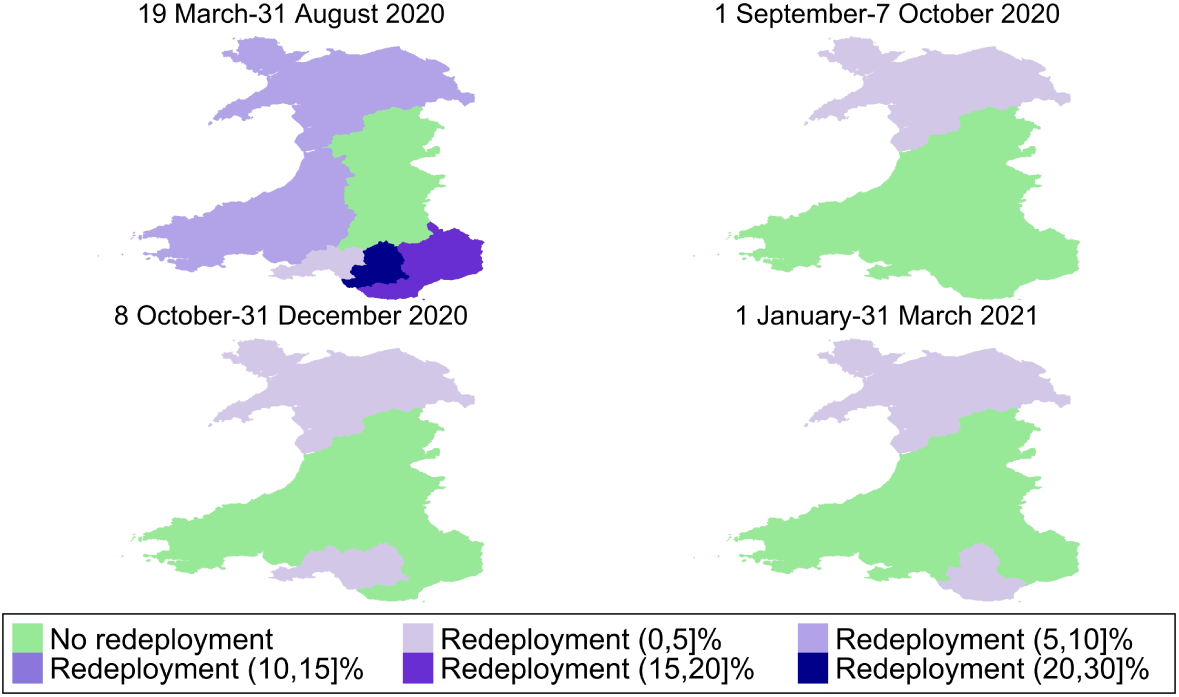
Redeployment of Health Visitors in Wales, March 2020-March 2021. Note: Redeployment is expressed as the percentage of total FTE HVs. Data provided by all seven Welsh Health Boards.

Turning to CSMS, Figure D5 shows redeployment patterns in Wales. Between 19 March and 31 August 2020, three Boards redeployed CSMS, often at higher percentages than their HV redeployment. No redeployment was reported between 8 October and 31 December 2020, but two Boards resumed redeployment between January and March 2021.

In Scotland, Figure D6 shows that half of all Boards redeployed CSMS between March and August 2020, and often at considerably higher percentages than HVs. Redeployment fell between September and early October 2020, when only three Boards reported redeployment, and was negligible by the end of the year.

Overall, redeployment patterns in Wales and Scotland mirrored those in England. CSMS were, on average, redeployed at higher shares than HVs, and redeployment peaked in the first months of the pandemic. Between March and August 2020, the average redeployment in Wales was 11.1% of all FTE HVs (ranging between 0% and 27.6%) and 15.7% of all FTE CSMS (0%-40.9%). In Scotland, it was 5.2% of HVs (0%-29.8%) and 19.5% of CSMS (0%-100%).

**Figure D4:**
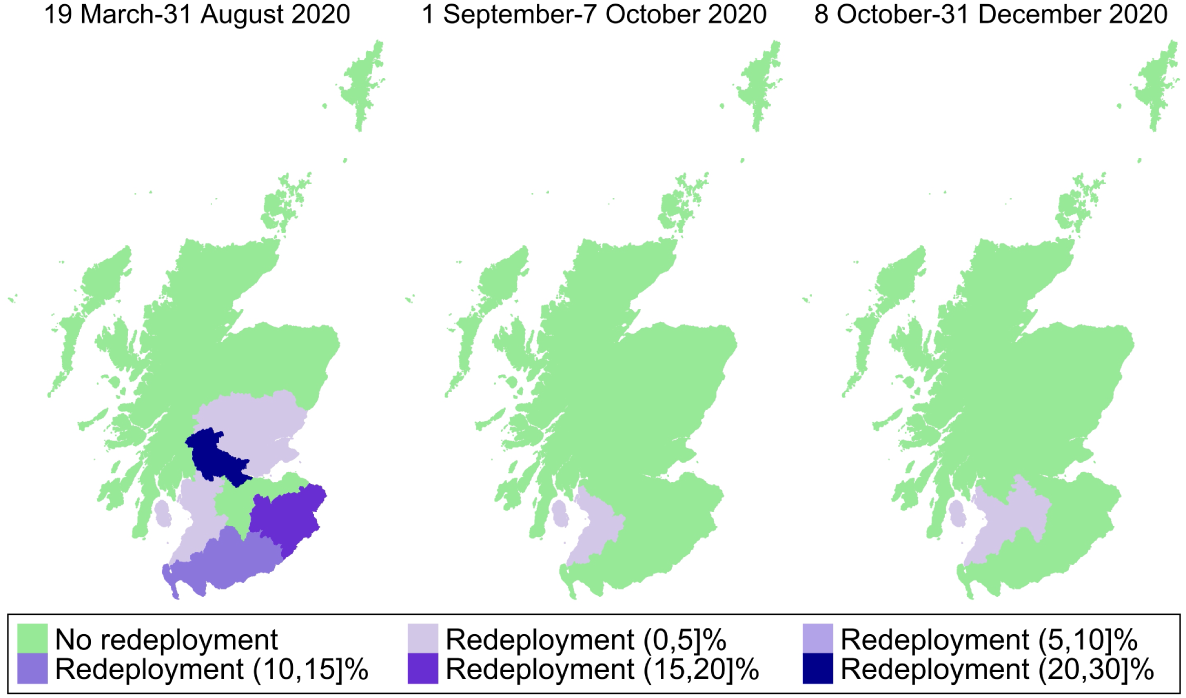
Redeployment of Health Visitors in Scotland, March-December 2020. Note: Redeployment is expressed as the percentage of total FTE HVs. Data provided by all fourteen Scottish Health Boards.

**Figure D5:**
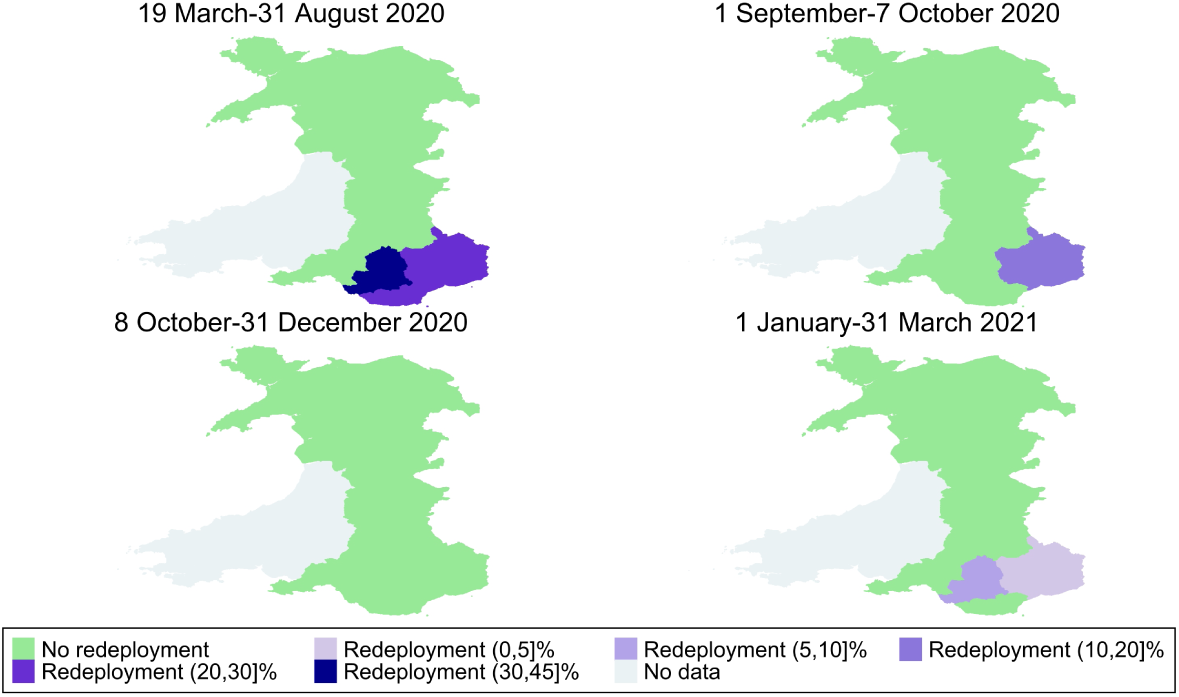
Redeployment of Clinical Skill Mix Staff in Wales, March 2020-March 2021. Note: Redeployment is expressed as the percentage of total FTE CSMS. Data provided by six Welsh Health Boards.

**Figure D6:**
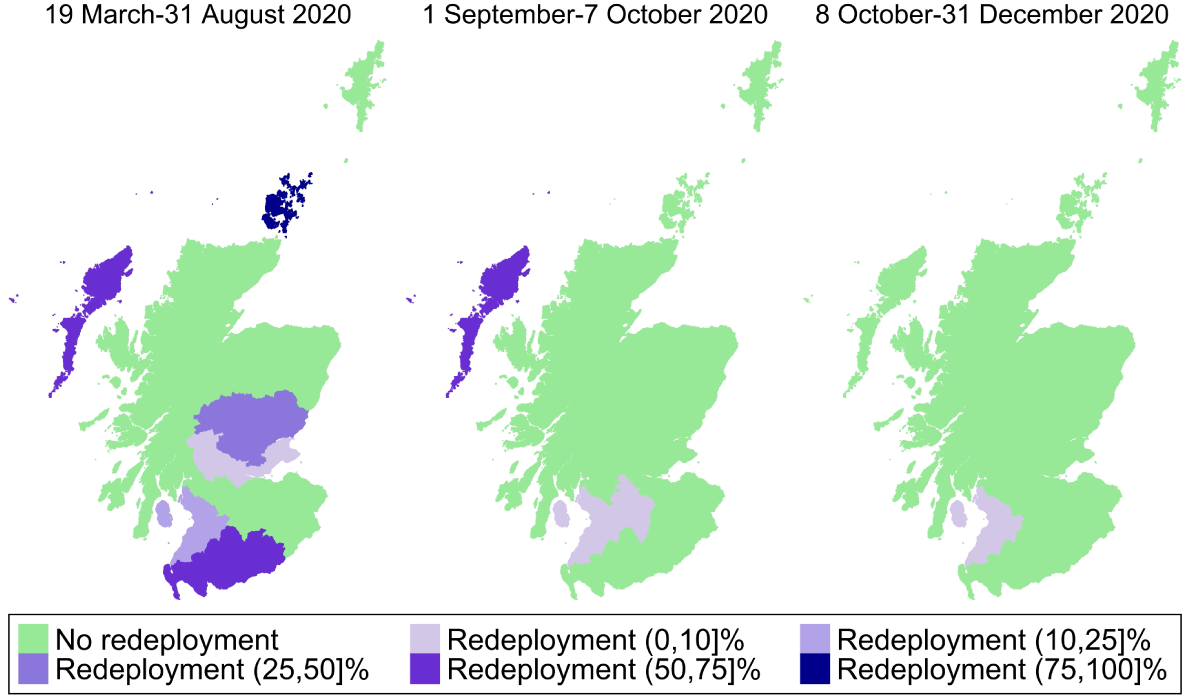
Redeployment of Clinical Skill Mix Staff in Scotland, March-December 2020. Note: Redeployment is expressed as the percentage of total FTE CSMS. Data provided by all fourteen Scottish Health Boards.

## Acknowledgements

Gabriella Conti, Dylan Celestino D’Mello and Yichen Yu gratefully acknowledge support from the European Research Council with the grant agreement 819752 DEVORHBIOSHIP-ERC-2018-COG under the European Unions Horizon 2020 research and innovation program; Gabriella Conti also gratefully acknowledges support from the Leverhulme Trust via the Philip Leverhulme Prize. This document updates and expands two previous reports by Conti and Dow: “Using FOI data to assess the state of Health Visiting Services in England before and during COVID-19” (2021), and “Rebuilding the health visiting workforce: costing policy proposals” (2021). We are very grateful to Abigail Dow for her effort and dedication in the initial phase of this research. We also thank Dr Michael Fanner and Professor Dame Sarah Cowley for their valuable input and many helpful discussions.

## Footnotes

1 The five health reviews are conducted at 28 weeks pregnancy (Antenatal Health Promoting Visit), at 10-14 days after birth (New Baby Review), at 6-8 weeks old (Six-Eight Week Review), at 9-12 months old (One Year Review), and at 2-2.5 years old (Two to Two-and-a-Half Year Review), during which health visiting team members build relationships with families to assess their needs, collect measurements of children’s health, growth, and development, provide the mother information on breastfeeding, nutrition, and healthy behaviours, and identify vulnerable or at-risk children. In 2021, two additional suggested (i.e., non-mandated) contacts were included in the guidance, at 3 and 6 months.

2 Available at: https://digital.nhs.uk/data-and-information/publications/statistical/nhs-workforce-statistics/.

3 Available at: https://digital.nhs.uk/data-and-information/publications/statistical/independent-healthcare-provider-workforce-statistics/.

4 Workforce data for NHS staff is collected through the NHS Electronic Staff Record and the workforce Minimum Data Set (wMDS) systems. Outside the National Health Service (NHS), some private providers might choose to make their workforce information public and/or submit it to some organisations (like the NHS Benchmarking Network); however, as such disclosure is made on a voluntary basis, the data is not available for all private providers.

5 The Community Services Dataset (CSDS) is a national collection managed by NHS England that records activity across community health services, including health visiting. While it has the potential to provide detailed information on service delivery, current completeness remains limited: only around 45% of mandated contacts recorded in the Health Visitor Service Delivery Metrics were also captured in CSDS between 2016 and 2020 (Clery et al., 2024). See https://digital.nhs.uk/data-and-information/data-collections-and-data-sets/data-sets/community-services-data-set for details about the data.

6 See Appendix A for further details.

7 For all LAs, including those served by non-NHS providers, we classified pay bands according to the NHS Agenda for Change (AfC) pay system for consistency. Specific job responsibilities and salaries may vary depending on the provider. As of the 2024/25 pay scales (https://www.nhsemployers.org/articles/pay-scales-202425), HVs earn between £37,338 and £44,962 depending on experience (Band 6). Team leaders and managers, often on Band 7, earn from £46,148 to £52,809. CSMS include roles such as clerical and healthcare assistants and support workers starting at Band 2 (£23,615) or Band 3 (£24,071 to £25,674), typically up to Band 4 health practitioners (£26,530 to £29,114), and Band 5 community or staff nurses (£29,970 to £36,483). See https://www.jobs.nhs.uk/candidate/search/results?keyword=Health%20Visitor&language=en for examples of current health visiting vacancies.

8 Further details about the data collection and methodology are available from the authors and in Appendix B, which includes the classification criteria and template.

9 We consider that an LA provided complete data when there is information on the number of total and caseload-holding FTE HVs and CSMS pay bands for all years, except when an inconsistent number was provided (e.g. FTE of staff with caseload > total FTE).

10 Although there were 151 LAs in total, four pairs provided combined data: Cornwall and Isles of Scilly, Hackney and City of London, Leicestershire and Rutland, and Essex and Thurrock. These are treated as single LAs throughout, and we combine their areas whenever we plot maps accordingly.

11 In some instances, pay bands with a small number of FTE were reported as “<5”, which we imputed as 2.5 FTE.

12 Additional follow-up requests were necessary to address incomplete responses in early waves: initial responses sometimes reported unexpectedly low pay bands for Health Visitors, prompting further follow-ups to amend any possible reporting errors and/or inconsistencies. For each year of data, we use the most recent FOI response received.

13 There was also one case of joint provision. In Suffolk, health visiting was delivered by Suffolk County Council Children and Young Peoples Services in East and West Suffolk, and by the social enterprise East Coast Community Health Care in North Suffolk until 31 March 2019. From 1 April 2019 the whole county moved to council provision. Our pre-2020 data for Suffolk refers only to the council-employed workforce in East and West Suffolk.

14 Table B3 in the Appendix displays the related figures for each year. See also Appendix C for workforce figures of London boroughs, and Appendix D for Wales and Scotland. Overall, London boroughs started and remained below the national average in the share of Health Visitors (62.8% vs. 73.6% in 2016; 55.7% vs. 63.8% in 2021), although the decline was slightly less uniform than elsewhere: around 42% of boroughs even recorded increases. In Wales and Scotland, HVs formed a much higher share of the workforce (78% and 83.2%, respectively) and caseloads were markedly lower, averaging 189 children per FTE in Wales and 159 in Scotland, compared with around 400 in England.

15 In the same period (February 2016 to February 2021), NHS workforce statistics show a decrease in the number of Health Visitors working in NHS trusts from 10,178 to 6,591 FTE, or 35.2%. This is not directly comparable to the 20.6% decline we see in our FOI data for two main reasons: (i) it refers to all of England, while we are restricted to 75 LAs; (ii) it only includes NHS trusts, while we also include other providers. It is not possible to restrict the NHS data only to the same providers in our FOI data, since any disaggregated numbers in the NHS workforce statistics are only available with a combined total of nurses and Health Visitors.

16 For general discussions of health workforce skill-mix and its potential implications, see Dubois and Singh (2009) and Spooner et al. (2022). Evidence from early home-visiting evaluations shows that delivery models and staffing configurations can affect maternal and child outcomes (Conti et al., 2024a, 2025; Schepan et al., 2025), but these studies do not directly evaluate substitution of registered Health Visitors with lower-banded staff in universal services.

17 In our FOI data collection, we asked LAs to report FTE HVs and CSMS ‘both with and without caseload’ and ‘only with caseload’. Throughout the report, we use the terms ‘with caseload’ and ‘caseload-holding’ interchangeably to refer to the latter. However, it is important to note that, while CSMS may hold a caseload *delegated* by a HV, the Health Visitor retains overarching accountability for all activities carried out within health visiting teams (Institute of Health Visiting, 2020). See also UK Government (2018) for an earlier mention of Health Visitors delegating work to other team members.

18 This number includes five LAs that had 0 FTE CSMS with caseload in 2016 and thus had an undefined percentage change in this period: Greenwich, Haringey, Islington, Suffolk, and West Sussex.

19 The skill mix of the team and the responsibilities of HVs are typically outlined in the service specification; however, the exact composition is determined by the provider to meet targets set in key performance indicators. Blackburn with Darwen Council (2015) is an example of service specification from the beginning of the period of our data collection.

20 It is worth noting that while we compute caseload size as the number of children under 5 (sourced from the Office for National Statistics’ mid-year population estimates) divided by the total number of FTE caseload-holding staff in health visiting teams (i.e., including both HVs and CSMS), the Institute of Health Visiting’s recommendation refers to a caseload of 250 per Health Visitor only (Institute of Health Visiting, 2017, 2020).

21 Note that our figure of an average caseload of 349.5 children per caseload-holding staff in 2021 is similar to that reported by NHS Benchmarking Network (2021), which indicates a mean caseload of 362 ‘per clinical whole-time equivalent in establishment’ in the universal provision of the HCP in the fiscal year 2020-21.

22 Caseloads can also follow a *combined model*, with individual allocation for a period followed by shared service provision (Whittaker et al., 2021).

23 This information was provided by the LAs or service providers themselves through various email exchanges. However, because we did not specifically ask LAs about caseload organisation in the FOI template, we cannot rule out that other LAs also followed a corporate model. Rochdale and Swindon did not provide complete data on FTE with and without caseloads, thus they are not included in the related sample.

24 These numbers were computed for the sample of 66 LAs in the *complete caseload FTE* sample. They correspond to averages across all years, and are statistically significantly different.

25 These examples of activities were retrieved from job descriptions of advertised community staff nurse positions in: https://www.jobs.nhs.uk/candidate/search/results?keyword=health%20visiting&language=en (accessed on 23/01/2025).

26 A previous version of this brief, focused on redeployment (Conti and Dow, 2021), contributed to the evidence base informing this guidance. See also the witness statement by Alison Morton (Director, Institute of Health Visiting) to the UK COVID-19 Inquiry: https://covid19.public-inquiry.uk/wp-content/uploads/2024/11/26175354/INQ000411557.pdf.

27 See also Appendix D for redeployment figures for Wales and Scotland. In both nations, redeployment peaked in the first months of the pandemic before declining sharply by late 2020. Average redeployment was higher among Clinical Skill Mix Staff (15.7% in Wales and 19.5% in Scotland) than among Health Visitors (11.1% and 5.2%, respectively), mirroring the pattern observed in England.

28 The Revenue Outturn: Social Care and Public Health Services (RO3) is part of the Local Authority Revenue Expenditure and Financing collection, published by the Department for Levelling Up, Housing and Communities and formerly by the Ministry of Housing, Communities and Local Government. We use figures reported under ‘Total Expenditures’ for ‘Mandated 0-5 children’s services (prescribed functions)’ and ‘Other 0-5 children’s services (non-prescribed functions)’. Figures have been adjusted to 2023-24 prices using the GDP deflator published in March 2025 (available at: https://www.gov.uk/government/collections/gdp-deflators-at-market-prices-and-money-gdp) and divided by the total population of children under five, based on ONS mid-year population estimates. The analysis begins in 2016-17, as funding for 2015-16 covered only part of the financial year.

29 Agenda for Change salaries attract High Cost Area Supplements (London weighting) for posts in Inner London, Outer London, and Fringe areas. Our cost estimates use national pay scales and therefore abstract from London weighting.

30 Band 4 includes only entry and top steps under the 2024/25 Agenda for Change scales, unlike Bands 57, which also have mid-points. As detailed in Table 1, we use the top point for Band 4 to reflect that most CSMS are employed at this level, with many on higher adjacent bands (as shown in Figure 4), while using mid-points for the other bands.

31 The real-terms per person public health grant allocation declined more in more deprived areas compared to the least deprived group between 2016 and 2021, as is shown in Figure B4 and confirmed by the cross-sectional tests and fixed effects regressions in Tables B5-B8. This is consistent with Thomas (2019) (2014-15–2019-20) and Finch et al. (2025) (2015-16–2025-26), both of whom find that cuts to local public health funding have typically been larger in more deprived areas.

32 Higher per-child spending in more deprived areas is also observed for non-mandated services, consistent with evidence from Fraser et al. (2022), who use CSDS data from 33 LAs and show that children in more deprived areas receive more health visiting contacts once non-mandated visits are taken into account.

33 To accurately reflect the caseload held by Health Visitors in Scotland due to the later school starting age, we also use data on the number of children under 6. Using this larger population as the numerator, the mean caseload in Scotland increases modestly to 184 children per FTE staff member.

34 The data for Scottish Health Boards covers the period until December 2020, whereas for Wales we have data until March 2021.

## References

Bhalotra, Sonia, Martin Karlsson, and Therese Nilsson (2017) “Infant Health and Longevity: Evidence from A Historical Intervention in Sweden,” Journal of the European Economic Association, 15 (5), 1101–1157, 10.1093/jeea/jvw028.

Blackburn with Darwen Council (2015) “Service Specification – Healthy Child Programme 0-5: Health Visiting Service,” https://democracy.blackburn.gov.uk/Data/Health%20&%20Wellbeing%20Board/201606211730/Agenda/Document%205.pdf, Accessed: 05/02/2025.

Clery, Amanda, Catherine Bunting, Mengyun Liu, Katie Harron, Jenny Woodman, and Louise Mc Grath-Lone (2024) “Can Administrative Data Be Used to Research Health Visiting in England? A Completeness Assessment of the Community Services Dataset,” International Journal of Population Data Science, 9 (1), 10.23889/ijpds.v9i1.2385.

Conti, Gabriella (2020) “The Economics of Prevention in the Early Years,” in Cowley, Sarah and Karen Whittaker eds. Community Public Health in Policy and Practice, 3rd edition: Elsevier Health Sciences, 10.1016/bs.hesedu.2022.11.005.

Conti, Gabriella and Abigail Dow (2021) “Using FOI Data to Assess the State of Health Visiting Services in England before and during COVID-19,” report, UCL Department of Economics, London, UK, https://discovery.ucl.ac.uk/id/eprint/10132710/.

Conti, Gabriella, Sören Kliem, and Malte Sandner (2024a) “Early home visiting delivery model and maternal and child mental health at primary school age,” in AEA Papers and Proceedings, 114, 401–406, American Economic Association 2014 Broadway, Suite 305, Nashville, TN 37203.

Conti, Gabriella, Malte Sandner, Tilman Brand, and Sören Kliem (2025) “Efficacy of delivery models in early intervention: Findings from Germany’s nurse-family partnership on family and child welfare services and pediatric medical incidents in high-risk families,” Child Abuse & Neglect, 166, 107513.

Conti, Gabriella, Joyce Smith, Elizabeth Anson, Susan Groth, Michael Knudtson, Andrea Salvati, and David Olds (2024b) “Early Home Visits and Health Outcomes in Low-Income Mothers and Offspring: 18-Year Follow-Up of a Randomized Clinical Trial,” JAMA network open, 7 (1), e2351752–e2351752.

Department for Education (2025) “Giving Every Child the Best Start in Life,” https://assets.publishing.service.gov.uk/media/686bd62a10d550c668de3be7/Giving_every_child_the_best_start_in_life.pdf.

Department of Health (2009) “Healthy Child Programme: Pregnancy and the first five years of life,” https://www.gov.uk/government/publications/healthy-child-programme-pregnancy-and-the-first-5-years-of-life, Accessed: 16/12/2024.

Department of Health (2011) “Health Visitor Implementation Plan 2011-15: A Call to Action,” https://assets.publishing.service.gov.uk/government/uploads/system/uploads/attachment_data/file/213110/Health-visitor-implementation-plan.pdf, Accessed: 16/12/2024.

Department of Health (2018) “Best start in life and beyond: Improving public health outcomes for children, young people and families. Guidance to support the commissioning of the Healthy Child Programme 0-19: Health visiting and school nursing services.”

Department of Health and Social Care (2025) “Fit for the Future: 10-Year Health Plan for England,” https://www.gov.uk/government/publications/10-year-health-plan-for-england-fit-for-the-future/fit-for-the-future-10-year-health-plan-for-Executive-Summary.

Dubois, Carl-Ardy and Debbie Singh (2009) “From staff-mix to skill-mix and beyond: towards a systemic approach to health workforce management,” Human resources for health, 7 (1), 87.

Duffee, James H, Alan L Mendelsohn, Alice A Kuo et al. (2017) “Early childhood home visiting,” Pediatrics, 140 (3), e20172150.

Finch, David, Anna Gazzillo, and Myriam Vriend (2025) “Investing in the public health grant: What it is and why greater investment is needed,” February, https://www.health.org.uk/reports-and-analysis/analysis/investing-in-the-public-health-grant.

Fraser, Caroline, Katie Harron, Jane Barlow, Samantha Bennett, Geoffrey Woods, Jenny Shand, Sally Kendall, and Jenny Woodman (2022) “Variation in health visiting contacts for children in England: cross-sectional analysis of the 2–2½ year review using administrative data (Community Services Dataset, CSDS),” BMJ Open, 12 (2), 10.1136/bmjopen-2021-053884.

Gulland, Anne (2017) “Spending on Public Health Cut as Councils Look to Save Money,” BMJ, 358, j3401, 10.1136/bmj.j3401.

Hjort, Jonas, Mikkel Sølvsten, and Miriam Wüst (2017) “Universal Investment in Infants and Long-Run Health: Evidence from Denmark’s 1937 Home Visiting Program,” American Economic Journal: Applied Economics, 9 (4), 78–104, 10.1257/app.20150087.

Institute of Health Visiting (2012) “A paper by Cheryll Adams on the History of Health Visiting,” https://ihv.org.uk/about-us/history-of-health-visiting/a-paper-by-cheryll-adams/, Accessed: 16/12/2024.

Institute of Health Visiting (2016) “PHE: New integrated 4-5-6 model and updated high impact areas,” https://ihv.org.uk/news-and-views/news/phe-new-integrated-4-5-6-model-updated-high-impact-areas/, Accessed: 18/02/2025.

Institute of Health Visiting (2017) “Health Visitors in England fear for some children’s futures as their numbers are reduced,” https://ihv.org.uk/wp-content/uploads/2017/12/171204-Institute-of-Health-Visiting-survey-results-safeguarding-children-2-12-17.pdf, Accessed: 26/02/2025.

Institute of Health Visiting (2018) “Position statement: Health visiting and the NHS in the next 10 years,” https://ihv.org.uk/news_tag/10-year-plan/, Accessed: 26/02/2025.

Institute of Health Visiting (2019) “Position statement: Worrying cuts to health visiting services across England: Ticking the box but missing the point,” https://ihv.org.uk/news-and-views/news/worrying-cuts-to-health-visiting-services-across-england-ticking-the-box-but-missing-the-poin Accessed: 26/02/2025.

Institute of Health Visiting (2020) “State of Health Visiting in England: Results from a survey of 1040 practising health visitors,” https://bit.ly/3rG0H8a, Accessed: 26/02/2025.

Institute of Health Visiting (2022) “Health visitor workforce numbers in England reach an all-time low,” https://ihv.org.uk/news-and-views/news/health-visitor-workforce-numbers-in-england-reach-an-all-time-low/, Accessed: 16/12/2024.

Institute of Health Visiting (2025) “State of Health Visiting 2024, UK Survey Report,” https://bit.ly/4hmR3Me, Accessed: 05/02/2025.

Institute of Health Visiting (n.d.) “History of Health Visiting,” https://ihv.org.uk/about-us/history-of-health-visiting/, Accessed: 16/12/2024.

Liu, Mengyun, Jenny Woodman, Louise Mc Grath-Lone et al. (2024) “Local area variation in health visiting contacts across England for children under age 5: a cross-sectional analysis of administrative data in England 2018–2020,” International Journal of Population Data Science, 9 (2), 2382, https://ijpds.org/article/view/2382.

Local Government Association (2020) “Joint letter on Winter Planning: Support to Children and Families, 7 October 2020,” Local Government Association, NHS and Public Health England, https://www.local.gov.uk/joint-letter-winter-planning-support-children-and-families-7-october-2020, Accessed: 03/03/2025.

NHS Benchmarking Network (2021) “Generic Community Services Report 2020/2021,” https://www.nhsbenchmarking.nhs.uk, Data covers the 2020/21 period from April 2020 to March 2021.

NHS Digital (n.d.a) “NHS Occupation Codes Manual,” https://digital.nhs.uk/data-and-information/areas-of-interest/workforce/nhs-occupation-codes, Accessed: 16/12/2024.

NHS Digital (n.d.b) “NHS Workforce Statistics,” https://digital.nhs.uk/data-and-information/publications/statistical/nhs-workforce-statistics, Accessed: 16/12/2024.

NHS England (2020a) “COVID-19 prioritisation within community health services,” https://web.archive.org/web/20200330104733/https://www.england.nhs.uk/coronavirus/publication/covid-19-prioritisation-within-community-health-services-with-annex_19-march-2020/, Archived publication. Accessed: 03/03/2025.

NHS England (2020b) “COVID-19 restoration of community health services for children and young people: second phase of NHS response,” https://www.england.nhs.uk/coronavirus/wp-content/uploads/sites/52/2020/03/C0552-Restoration-of-Community-Health-Services-Guidance-CYP-version-3-June-2020-1.pdf, Archived publication. Accessed: 03/03/2025.

NHS England (2020c) “Next steps on NHS response to COVID-19,” NHS England and NHS Improvement, https://www.england.nhs.uk/coronavirus/publication/next-steps-on-nhs-response-to-covid-19-letter-from-simon-stevens-and-amanda-pritchard/, Accessed: 03/03/2025.

Reid, Bernie and Julie Tracey (2023) “Health visitor workload: an integrative review of the literature,” Primary Health Care, 33 (4), 10.7748/phc.2023.e1804.

Schepan, Marie Lisanne, Malte Sandner, Gabriella Conti, Sören Kliem, and Tilman Brand (2025) “Maternal and Child Health Following 2 Home Visiting Interventions vs Control: Five-Year Follow-Up of a Randomized Clinical Trial,” JAMA pediatrics, 179 (4), 367–374.

Scottish Government (2020) “Coronavirus (COVID-19): Nursing and Community Health Staff Guidance,” https://webarchive.nrscotland.gov.uk/20200901190114/https://www.gov.scot/publications/coronavirus-covid-19-nursing-and-community-health-staff-guidance/, Archived publication. Accessed: 16/12/2024.

Spooner, Sharon, Imelda McDermott, Mhorag Goff, Damian Hodgson, Anne McBride, and Katherine Checkland (2022) “Processes supporting effective skill-mix implementation in general practice: A qualitative study,” Journal of health services research & policy, 27 (4), 269–277.

Thomas, Chris (2019) “Hitting the poorest worst? How public health cuts have been experienced in Englands most deprived communities,” November, https://www.ippr.org/articles/public-health-cuts.

UK Government (2018) “Best start in life and beyond: Improving public health outcomes for children, young people and families,” https://web.archive.org/web/20190725071109/https://assets.publishing.service.gov.uk/government/uploads/system/uploads/attachment_data/file/686928/best_start_in_life_and_beyond_commissioning_guidance_1.pdf, Archived publication. Accessed: 05/02/2025.

UK Government (2021) “Health Visiting and School Nursing Service Delivery Model,” https://www.gov.uk/government/publications/commissioning-of-public-health-services-for-children, Accessed: 16/12/2024.

Welsh Government (2020) “Healthy Child Wales Programme: 2020; Quality and Methodology Information: Changes to the Programme Due to the COVID-19 Pandemic,” https://gov.wales/healthy-child-wales-programme-2020-html/, Accessed: 16/12/2024.

Whittaker, Karen, Jane V Appleton, Sue Peckover, and Cheryll Adams (2021) “Organising health visiting services in the UK: Frontline perspectives,” Journal of Health Visiting, 9 (2), 68–75, 10.12968/johv.2021.9.2.68.

